# A Systematic Review and Meta-Analysis of Automated Methods for Quantifying Enlarged Perivascular Spaces in the Brain

**DOI:** 10.1101/2024.03.04.24303705

**Authors:** Jennifer M.J. Waymont, Maria C. Valdés Hernández, Jose Bernal, Roberto Duarte Coello, Rosalind Brown, Francesca M. Chappell, Lucia Ballerini, Joanna M. Wardlaw

## Abstract

Research into magnetic resonance imaging (MRI)- visible perivascular spaces (PVS) has recently increased, as results from studies in different diseases and populations are cementing their association with sleep, disease phenotypes, and overall health indicators. With the establishment of worldwide consortia and the availability of large databases, computational methods that allow to automatically process all this wealth of information are becoming increasingly relevant. Several computational approaches have been proposed to assess PVS from MRI, and efforts have been made to summarise and appraise the most widely applied ones. We systematically reviewed and meta-analysed all publications available up to September 2023 describing the development, improvement, or application of computational PVS quantification methods from MRI. We analysed 67 approaches and 60 applications of their implementation, from 112 publications. The two most widely applied were the use of a morphological filter to enhance PVS-like structures, with Frangi being the choice preferred by most, and the use of a U-Net configuration with or without residual connections. Older adults or population studies comprising adults from 18 years old onwards were, overall, more frequent than studies using clinical samples. PVS were mainly assessed from T2-weighted MRI acquired in 1.5T and/or 3T scanners, although combinations using it with T1-weighted and FLAIR images were also abundant. Common associations researched included age, sex, hypertension, diabetes, white matter hyperintensities, sleep and cognition, with occupation-related, ethnicity, and genetic/hereditable traits being also explored. Despite promising improvements to overcome barriers such as noise and differentiation from other confounds, a need for joined efforts for a wider testing and increasing availability of the most promising methods is now paramount.

## 1. Introduction

Perivascular spaces (PVS), also referred to as ‘Virchow-Robin spaces’ (VRS), are fluid-filled cavities surrounding the blood vessels of the brain. PVS are thought to facilitate the uptake of cerebrospinal fluid (CSF) and the removal of metabolic waste products from the brain, but the precise involvement in these processes in humans remains elusive (Wardlaw et al., 2020). PVS are not always visible in brain imaging or at autopsy, but are increasingly visible as they become enlarged. Enlarged PVS are associated with increased age (Francis et al., 2019), other markers of small vessel disease (SVD; Wardlaw et al., 2013, Duering et al., 2023), and may be more prevalent in individuals with vascular risk factors (e.g., hypertension; Zhu et al., 2010). A high burden of enlarged PVS may be associated with worse brain health outcomes, including increased risk of vascular dementia (more so than Alzheimer’s disease (AD); Smeijer et al., 2019) and stroke (Debette et al., 2019).

PVS appear as ovoid or tubular structures, with bright (hyperintense) signal in T2-weighted (T2w) magnetic resonance imaging (MRI) and dark (hypointense) signal in T1-weighted (T1w) MRI, and are occasionally visible on fluid-attenuated inversion recovery (FLAIR) imaging (Wardlaw et al., 2013; Duering et al., 2023). Larger ovoid PVS may appear similar to lacunes—also round CSF-filled cavities and thus with similar signal characteristics to PVS (Valdés Hernández et al., 2013; Duering et al., 2023). PVS may also overlap with one another, exhibit tortuosity, have irregular diameters, and may traverse adjacent imaging slices, complicating their quantification when using multiple 2D slices from images with anisotropic voxels, commonly acquired in clinical practice.

Several automated and semi-automated computational pipelines and methods have been developed to quantify PVS. These methods are heterogeneous in terms of their implementation, validation, and application (Barisano et al., 2022; Moses et al., 2023; Pham et al., 2022). The principles and drawbacks of some of these methods have been summarised previously (Barisano et al., 2022; Moses et al., 2023; Pham et al., 2022), and suggestions about further requirements to increase the understanding of PVS by means of their computational assessment using MRI have been drawn (Pham et al., 2022). But there has been also a wealth of computational developments to either enhance performance of previously developed methods, establish their limits of validity, or reduce noise effects, or increase the accuracy of their output, which have not been summarised or reviewed. Neither has this wealth of information been systematically identified or meta-analysed.

The relevance of PVS has become increasingly evident in areas not studied before such as in relation to the occlusion of sinonasal cavities (Sáenz de Villaverde Cortabarría et al., 2023; Valdés Hernández et al., 2023), in patients with the metabolic syndrome (Hayden, 2023), or in patients with amyotrophic lateral sclerosis (Schreiber et al., 2023), and, with it, the efforts in improving the accuracy of the currently accepted as state-of-the-art computational methods to assess them in the presence of confounding pathology such as white matter hyperintensities (WMH) or lacunes.

Consequently, we systematically reviewed and meta-analysed the literature in an attempt to fill this void. In this systematic literature review of (semi-/fully-) automated quantification of PVS, we not only identify and meta-analyse the data from the PVS quantification methods that have been developed, and summarise how they have been applied in studies of PVS associations with health and lifestyle indicators, but also analyse the improvements to these methods, and the computational efforts around this topic.

## 2. Methods

### 2.1. Protocol registration

We registered this systematic review protocol with the International Prospective Register of Systematic Reviews (PROSPERO), registration number: CRD42022359951 (September 2022). We planned to meta-analyse estimates where methodological and clinical characteristics of contributing studies were similar.

### 2.2. Search strategy

We searched PubMed, Web of Science, and Google Scholar for literature published between January 1990 and September 2023. We also included any hand-selected papers identified from references in relevant literature reviews. Three reviewers (JMJW, MCVH, and JB) conducted the literature search. Each paper was assessed by two reviewers and discrepancies were resolved by discussion. After running pilot searches, we decided on the following search terms (Table 1).

**Table 1.** Search strategy.

| Table 1. Search strategy |  |
| --- | --- |
| Grouping | Terms |
| #1 | “Perivascular spaces” OR “PVS” OR “EPVS” OR “Virchow-Robin spaces” OR “VRS” |
| #2 | “Enlarged” OR “Dilated” OR “Visible” OR “Dysfunction” OR “Enlargement” OR “Visibility” OR “Dilation” |
| #3 | #1 AND #2 |
| #4 | Comput* OR “Segmentation” OR Quantif* OR Algorithm* OR “Estimation” OR Automat* OR Semi-automat* OR Semiauto* |
| **Search: #3 AND #4** |  |

### 2.3. Inclusion and exclusion criteria

We included studies that described the development, improvement, or application of computational (i.e., semi-/automated) PVS quantification methods in human brain magnetic resonance imaging (MRI).

Exclusion criteria included records without accompanying full-text; studies not reported in English; animal model/pre-clinical studies; studies of PVS using only visual or manual (i.e., non-computational) PVS quantification; studies reporting computational lesion quantification that has not been applied to PVS; studies reporting results from the application of a computational PVS quantification methods but which did not cite or describe the PVS quantification method in-text or in a prior publication or provide any detail. Studies on non-parenchymal fluid dynamics and glymphatic clearance function metrics were also excluded.

### 2.4. Data extraction and risk of bias assessment

Six reviewers (JMJW, JB, RDC, RB, LB, and MCVH) extracted the data. To cross-check data entry, a reviewer (MCVH) performed double extraction independently and blind to extraction results. We grouped studies into ‘method development’ (i.e., the paper focuses primarily on the development and validation of a new proposal for identifying, quantifying, or segmenting PVS), ‘improvement/validation’ (i.e., the paper either provides evidence of a certain PVS quantification issue or proposes an approach to deal with it), and ‘application’ (i.e., the paper uses a previously presented approach to assess PVS to analyse results in relation to clinical data) categories, and extracted relevant data accordingly. For all studies, we extracted data relating to year of publication, authors, MR sequences/field strengths, population type and sample size. For ‘method development’ and ‘improvement/validation’ studies, we extracted data relating to pre-processing steps, reference standards, computational PVS method used, and accuracy and/or validation results. For ‘application’ studies, we additionally extracted more detailed information on population type, variables assessed in relation to PVS burden, and results.

We conducted a risk of bias assessment on included studies, based on the QUADAS-2 quality assessment tool (Whiting et al., 2011). Risk of bias plots were produced using the Risk of Bias Visualization tool (McGuinness & Higgins, 2020).

## 3. Results

### 3.1. Search results

We identified 2978 potentially relevant records. Before screening, we removed 354 results because they were duplicates or were not in English. During title and abstract screening, we screened titles and abstracts of 2624 records, and excluded 2427 of them. We sought retrieval of the remaining 197.

Among these, five were not accessible either publicly or via our institutional library, five were conference abstracts, and two considered PVS quantification for histology samples. Therefore, 185 studies were ultimately assessed for eligibility. From them, 73 studies were ineligible and 112 studies that met our criteria were included in this review. A flow chart of the identification and screening process is provided in Figure 1.

**Figure 1.**
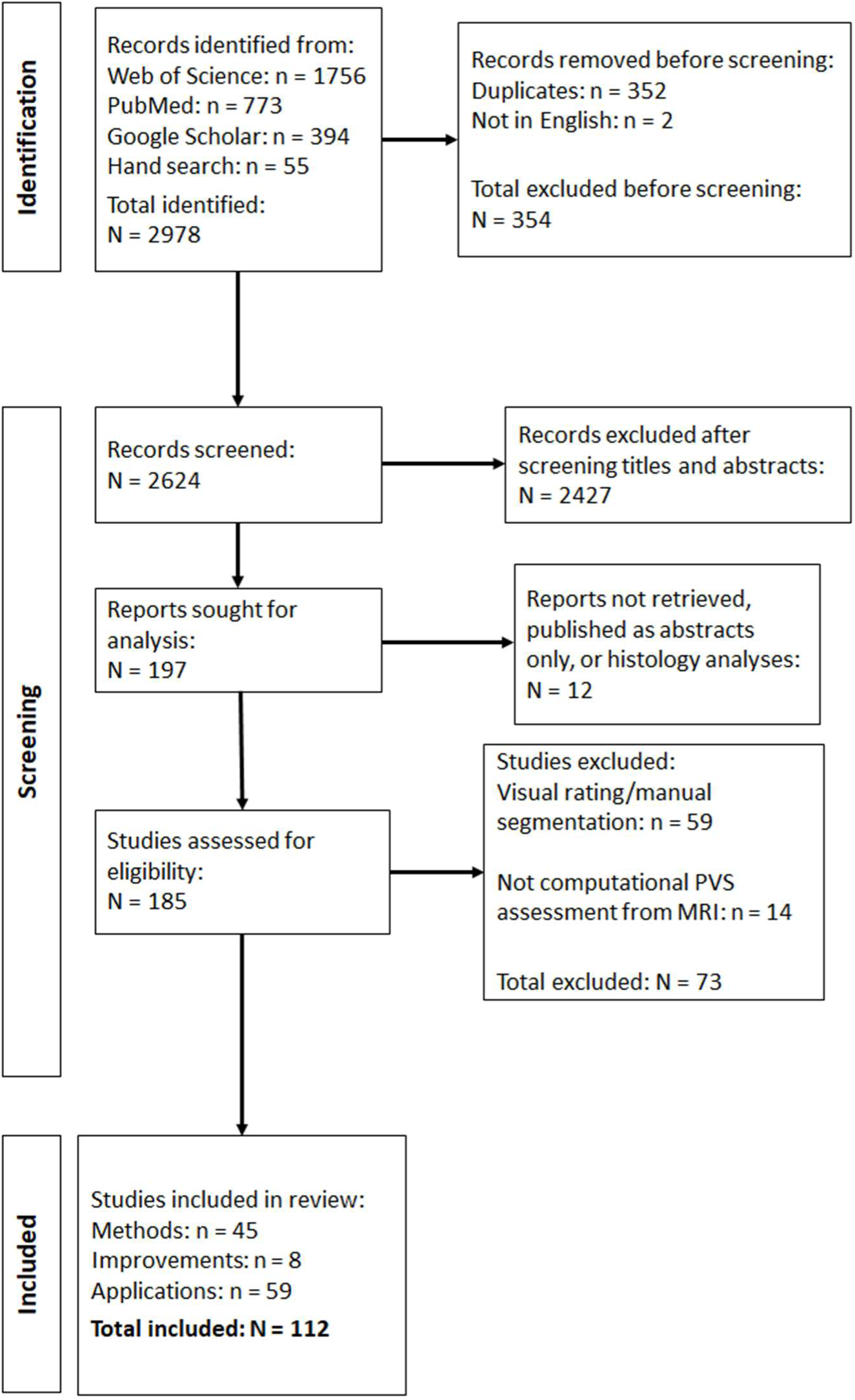
Flow chart of identification, screening, and selection process.

### 3.2. Overview of included studies

Of the 112 included studies, 45 fitted under ‘method development’, eight under ‘improvement’, and 59 under ‘application’. Four separate PVS quantification methods participated in the Vascular Lesions Detection Challenge 2021, as documented in (Sudre et al., 2022). We treat them as four distinct records, thus increasing the count of ’method development’ studies to 48, and the total number of studies to 115. Overviews of all included studies within these three categories are presented in their respective subsections (method development: 3.3, Table 6; improvement: 3.4, Table 8; application: 3.5, Table 9).

#### 3.3.1. Publication timeline

The earliest record identified was published in 2002 (Kruggel et al., 2002). Method development publications became more prevalent from around 2016 onwards reaching a peak in 2019 (16.67% from all publications in this category), and publications applying computational PVS quantification increased from around 2020 (Figure 2). Supplementary Table 1 provides a breakdown of publications by type and year.

**Figure 2.**
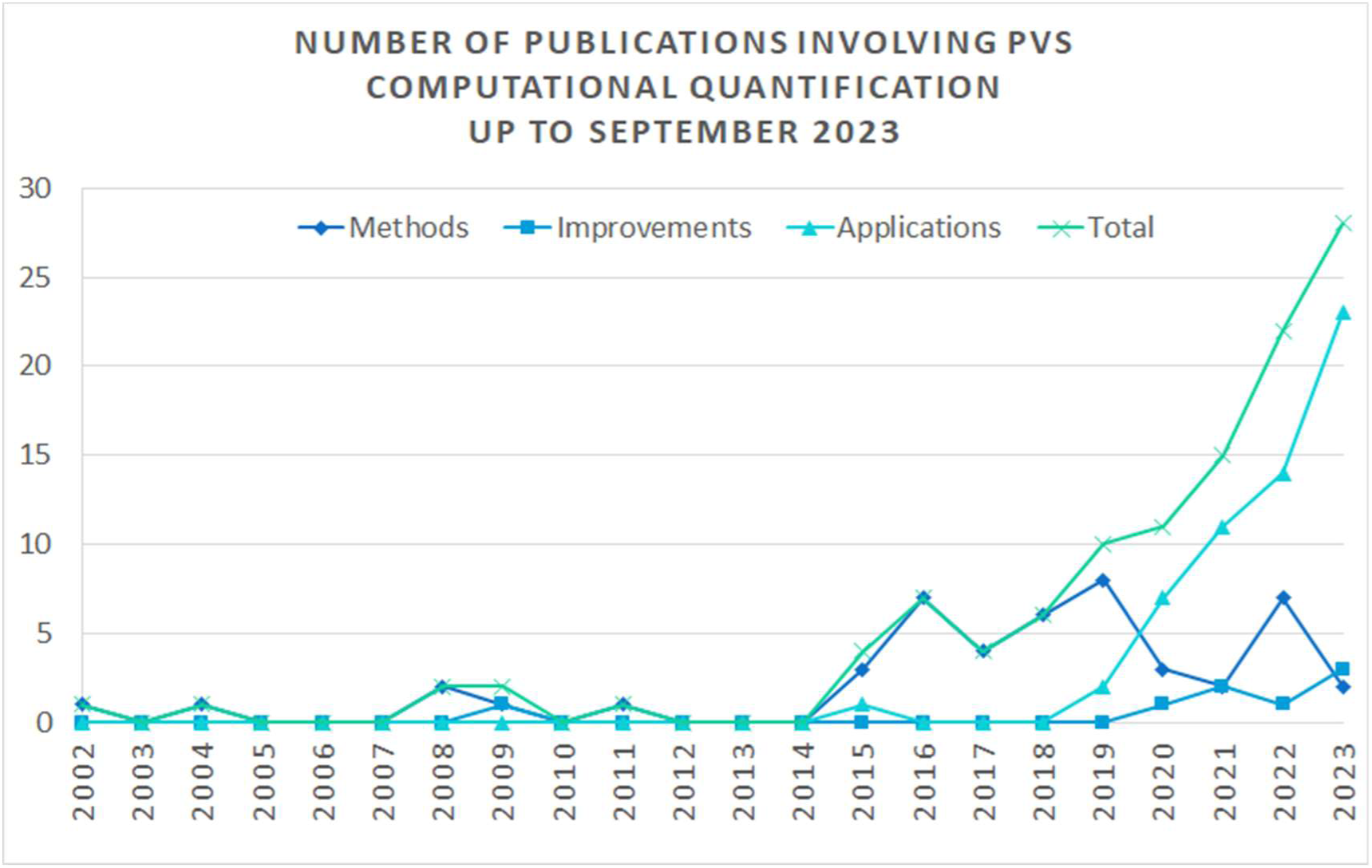
Line graph showing number of computational PVS quantification papers published between 2002 and September 2023.

#### 3.2.2. Populations and sample sizes

Clinical (i.e., patient) and non-clinical (e.g., healthy volunteers, community cohorts) samples have been used in the development, improvement, and application of computational PVS quantification methods, with non-clinical samples (e.g., community-dwelling cohorts) forming the largest proportion by a small margin in ’method developments’ and ‘improvement’ groups. Four sources describing methods (Uchiyama et al., 2008,2009; Zhang et al., 2016, 2017) did not specify the type of population. Table 1 shows population type overall and by method/improvement/application categories. From the 51 studies that included a clinical sample, 23 included a control population.

From the clinical samples, patients with cerebrovascular or neurodegenerative diseases were more represented than patients with psychiatric conditions, communicable diseases, or genetic diseases.

**Table 1.**
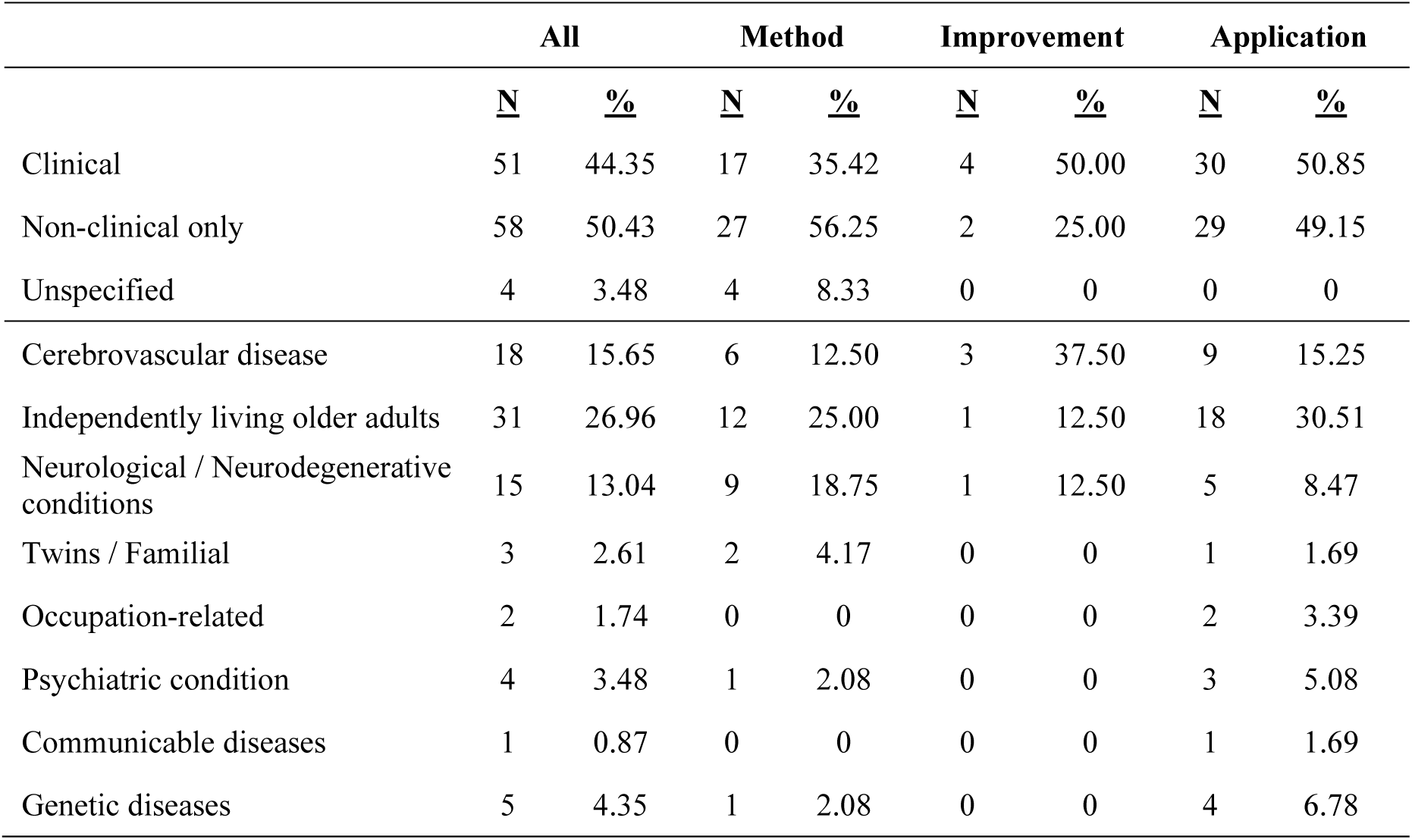
Population types and clinically-relevant demographic groups.

|  | All |  | Method |  | Improvement |  | Application |  |
| --- | --- | --- | --- | --- | --- | --- | --- | --- |
|  | <u>N</u> | <u>%</u> | <u>N</u> | <u>%</u> | <u>N</u> | <u>%</u> | <u>N</u> | <u>%</u> |
| Clinical | 51 | 44.35 | 17 | 35.42 | 4 | 50.00 | 30 | 50.85 |
| Non-clinical only | 58 | 50.43 | 27 | 56.25 | 2 | 25.00 | 29 | 49.15 |
| Unspecified | 4 | 3.48 | 4 | 8.33 | 0 | 0 | 0 | 0 |
| Cerebrovascular disease | 18 | 15.65 | 6 | 12.50 | 3 | 37.50 | 9 | 15.25 |
| Independently living older adults | 31 | 26.96 | 12 | 25.00 | 1 | 12.50 | 18 | 30.51 |
| Neurological / Neurodegenerative conditions | 15 | 13.04 | 9 | 18.75 | 1 | 12.50 | 5 | 8.47 |
| Twins / Familial | 3 | 2.61 | 2 | 4.17 | 0 | 0 | 1 | 1.69 |
| Occupation-related | 2 | 1.74 | 0 | 0 | 0 | 0 | 2 | 3.39 |
| Psychiatric condition | 4 | 3.48 | 1 | 2.08 | 0 | 0 | 3 | 5.08 |
| Communicable diseases | 1 | 0.87 | 0 | 0 | 0 | 0 | 1 | 1.69 |
| Genetic diseases | 5 | 4.35 | 1 | 2.08 | 0 | 0 | 4 | 6.78 |

Age groups of populations ranged from children and adolescents through to older adults. Studies comprising only older adults (60+ years) and population studies including adults from 18 years old onwards prevailed. Ten studies (predominantly method development studies) did not provide information about participants’ age. Table 2 shows population age groupings used in all and each of the study categories.

**Table 2.** Population age groups.

|  | All |  | Method |  | Improvement |  | Application |  |
| --- | --- | --- | --- | --- | --- | --- | --- | --- |
|  | <u>N</u> | <u>%</u> | <u>N</u> | <u>%</u> | <u>N</u> | <u>%</u> | <u>N</u> | <u>%</u> |
| Older adult (>60) | 34 | 29.56 | 10 | 20.83 | 1 | 12.50 | 22 | 37.29 |
| Middle adult (40-59) | 3 | 2.61 | 0 | 0 | 0 | 0 | 3 | 5.08 |
| Young adult (18-39) | 10 | 8.70 | 7 | 14.58 | 0 | 0 | 3 | 5.08 |
| Adult (18+) | 23 | 20.00 | 10 | 20.83 | 3 | 37.50 | 10 | 16.95 |
| Mid/Older adult (>40) | 18 | 15.65 | 11 | 22.92 | 0 | 0 | 7 | 11.86 |
| Young/Mid adult (18-59) | 4 | 3.48 | 1 | 2.08 | 1 | 12.50 | 2 | 3.39 |
| Child and adolescent (<18) | 4 | 3.48 | 0 | 0 | 0 | 0 | 5 | 8.47 |
| Child, adolescent, and young adult (<40) | 1 | 0.87 | 0 | 0 | 0 | 0 | 1 | 1.69 |
| Adult unspecified | 3 | 2.61 | 0 | 0 | 1 | 12.50 | 2 | 3.39 |
| Older adult unspecified | 3 | 2.61 | 2 | 4.17 | 0 | 0 | 1 | 1.69 |
| Young adult unspecified | 1 | 0.87 | 1 | 2.08 | 0 | 0 | 0 | 0 |
| Unspecified | 10 | 8.70 | 6 | 12.50 | 2* | 25.00 | 3 | 5.08 |
| Total | 115 | 100 | 48 | 100 | 8 | 100 | 59 | 100 |

Samples sizes ranged from one to 39,976 participants. Two studies that used *in-silico* and/or physical phantoms (Duarte Coello et al., 2023, 2024) did not involve the use of human data. The median sample size across all study types was 106 participants (IQR = 456.5). Application studies had the largest median sample size (161 participants) but also the greatest variance (IQR = 482.5). Sample sizes distribution by groups are shown in Table 3.

**Table 3.** Range and average sample sizes (N)

| Study categories | Min | Max | Median | IQR [Q1 Q3] |
| --- | --- | --- | --- | --- |
| All | 0 | 39,976 | 106 | [38 494.5] |
| Method development | 1 | 2202 | 86 | [19 272] |
| Improvement | 0 | 747 | 50 | [7.5 102] |
| Application | 12 | 39,976 | 161 | [57.5 540] |

#### 3.2.3. MRI sequences and field strengths

The included studies used a variety of MRI sequences, including T1w, T2w, proton density (PD), fluid-attenuated inversion recovery (FLAIR), and T2*w. Across all study types, the combined use of T1w and T2w MRI was the most frequently used (27.83% of studies), followed by the use of T2w imaging only (20.87% of all studies). One study used synthesised T2-type images for the development of a Digital Reference Object to evaluate PVS enhancement methods (Bernal et al., 2022). Although many studies of all types reported a combination of two or more sequence types, the majority used a single sequence, either T1w or T2w, to identify the PVS. Frequencies of individual and combined sequences are presented in Table 4.

**Table 4.**
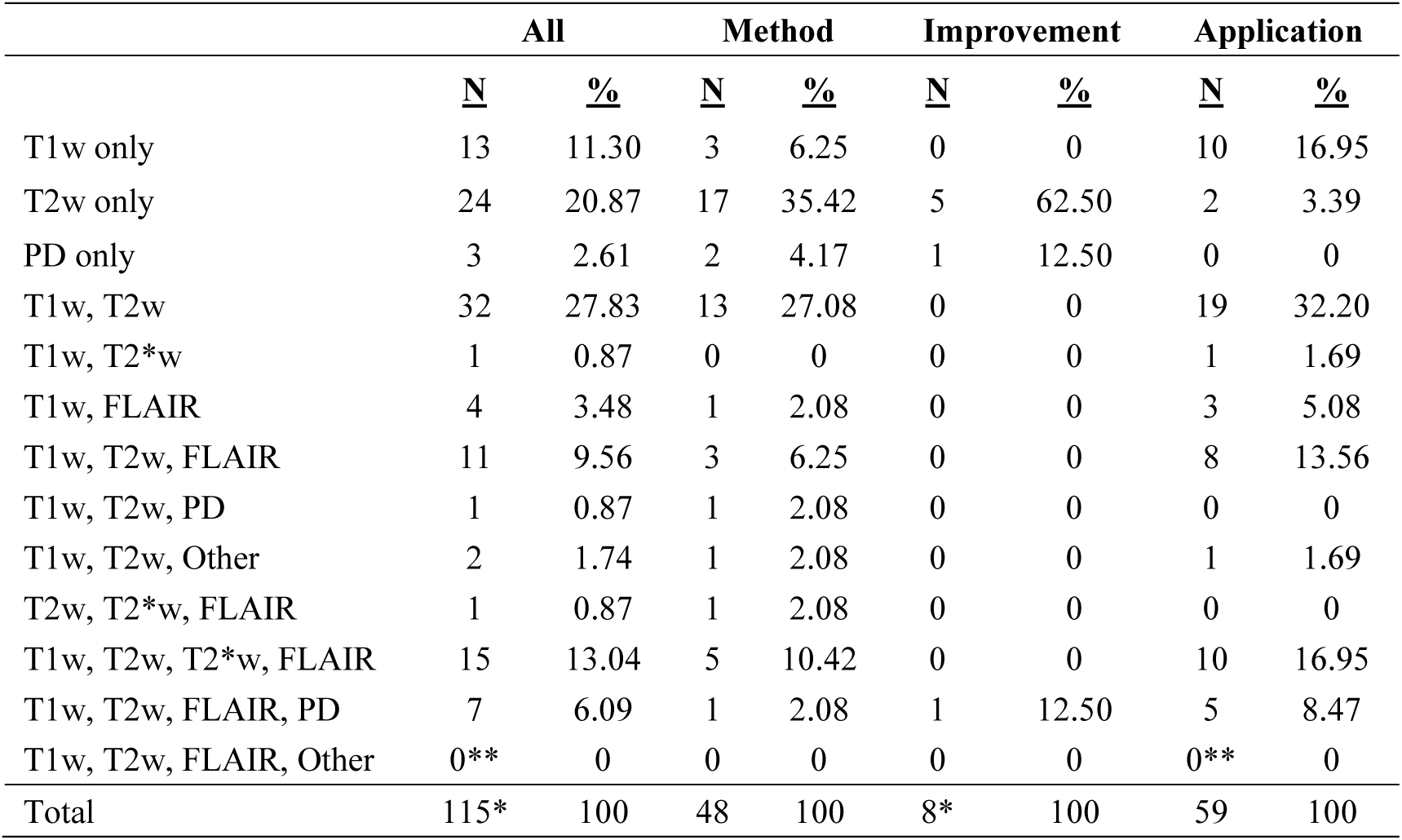
MR sequence combinations Legend: N: number of sources, %: Percentage with respect to the number of sources in each group (*: MRI sequences are not relevant for one of the Improvement papers. **: An application study has T1w, T2w, FLAIR, and 3DFlow MRI, but only uses T1w and T2w for PVS segmentation.)

|  | All |  | Method |  | Improvement |  | Application |  |
| --- | --- | --- | --- | --- | --- | --- | --- | --- |
|  | <u>N</u> | <u>%</u> | <u>N</u> | <u>%</u> | <u>N</u> | <u>%</u> | <u>N</u> | <u>%</u> |
| T1w only | 13 | 11.30 | 3 | 6.25 | 0 | 0 | 10 | 16.95 |
| T2w only | 24 | 20.87 | 17 | 35.42 | 5 | 62.50 | 2 | 3.39 |
| PD only | 3 | 2.61 | 2 | 4.17 | 1 | 12.50 | 0 | 0 |
| T1w, T2w | 32 | 27.83 | 13 | 27.08 | 0 | 0 | 19 | 32.20 |
| T1w, T2*w | 1 | 0.87 | 0 | 0 | 0 | 0 | 1 | 1.69 |
| T1w, FLAIR | 4 | 3.48 | 1 | 2.08 | 0 | 0 | 3 | 5.08 |
| T1w, T2w, FLAIR | 11 | 9.56 | 3 | 6.25 | 0 | 0 | 8 | 13.56 |
| T1w, T2w, PD | 1 | 0.87 | 1 | 2.08 | 0 | 0 | 0 | 0 |
| T1w, T2w, Other | 2 | 1.74 | 1 | 2.08 | 0 | 0 | 1 | 1.69 |
| T2w, T2*w, FLAIR | 1 | 0.87 | 1 | 2.08 | 0 | 0 | 0 | 0 |
| T1w, T2w, T2*w, FLAIR | 15 | 13.04 | 5 | 10.42 | 0 | 0 | 10 | 16.95 |
| T1w, T2w, FLAIR, PD | 7 | 6.09 | 1 | 2.08 | 1 | 12.50 | 5 | 8.47 |
| T1w, T2w, FLAIR, Other | 0** | 0 | 0 | 0 | 0 | 0 | 0** | 0 |
| Total | 115* | 100 | 48 | 100 | 8* | 100 | 59 | 100 |

The most commonly used MR field strengths were, in order, 3T, 1.5T, and 7T. Method development studies used 1.5T imaging more often than 3T or 7T (50.00%, 33.33%, 22.92%, respectively), with seven studies using MRI acquired at two different magnetic fields and one study (Dubost, Dünnwald, et al., 2019c) using MRI acquired at three different magnetic fields: 1T, 1.5T and 3T. In general, 17 studies used imaging from more than one field strength, typically a combination of 1.5T and 3T images or of 3T and 7T. Five ‘method development’ studies and one ‘improvement’ study (Paz et al., 2009) did not report the magnetic field of the MRI used. Application studies more commonly used 3T imaging (71.19%). One application study (Jokinen et al., 2020) used a small number of images from a 0.5T scanner. Frequencies of field strengths used in the included studies are presented in Table 5.

**Table 5.** MRI field strengths used Legend: N: number of sources, %: Percentage with respect to the number of sources in each group

|  | All |  | Method |  | Improvement |  | Application |  |
| --- | --- | --- | --- | --- | --- | --- | --- | --- |
|  | <u>N</u> | <u>%</u> | <u>N</u> | <u>%</u> | <u>N</u> | <u>%</u> | <u>N</u> | <u>%</u> |
| 1.5T | 45 | 39.13 | 24 | 50.00 | 1 | 14.28 | 20 | 33.90 |
| 3T | 61 | 53.04 | 16 | 33.33 | 3 | 42.86 | 42 | 71.19 |
| 7T | 18 | 15.65 | 11 | 22.92 | 2 | 28.57 | 5 | 8.47 |
| 1T | 1 | 0.87 | 1 | 2.08 | 0 | 0 | 0 | 0 |
| 0.5T | 1 | 0.87 | 0 | 0 | 0 | 0 | 1 | 1.69 |
| Unclear/Unreported | 6 | 5.22 | 5 | 10.42 | 1 | 14.28 | 0 | 0 |
| More than one magnetic field | 17 | 14.78 | 8 | 29.17 | 0 | 0 | 9 | 15.25 |

#### 3.2.4. Voxel sizes and slice thickness

Voxel sizes varied amongst the samples. Fourteen out of the 48 methods proposed were 2D, owed to the anisotropy of the voxels in the images of the samples used. Consequently, application studies using those methods comprised images with largely anisotropic voxels (e.g., 0.4 x 0.4 x 5 or 6 mm^3^) (Tables 6 and 9). But not all 3D methods studies used images with isotropic voxels, as in many cases pipelines involved resampling and interpolation to convert the images to 1 mm isotropic (Table 6).

### 3.3 Method development studies

Summaries of all method development studies (N = 48) are presented in Table 6, ordered by year of publication. Twenty-seven were journal articles, 16 published in conference proceedings, and five in preprint repositories. It is worth noting that the four methods showcased by Sudre et al. (2022) submitted as part of the challenge organised by the publication authors were, at the time this review was conducted, in a preprint repository. Despite the semi-automatic method proposed by Smith et al. (2020) being also at a preprint repository, this method per-se has already been applied by a clinical study (Langan et al., 2022).

#### 3.3.1. Computational methods

Seven of the 48 ‘method development’ studies used a semi-automated approach, while the remaining 41 used a fully automated one. Broadly speaking, computational methods included filtering, machine learning, deep learning, combined approaches and intensity thresholding (i.e., as main technique) (Table 6). It is worth noting that most, if not all papers, involve thresholding. CNNs for example, often output a response map that is thresholded to get the binary masks. Also, filtering techniques generate a response that is thresholded to obtain the PVS proxies. The most frequent approach was the use of deep learning, proposed in 16 studies (33.33%). Frequencies of all types of approaches proposed are presented in Table 7.

**Table 6.**
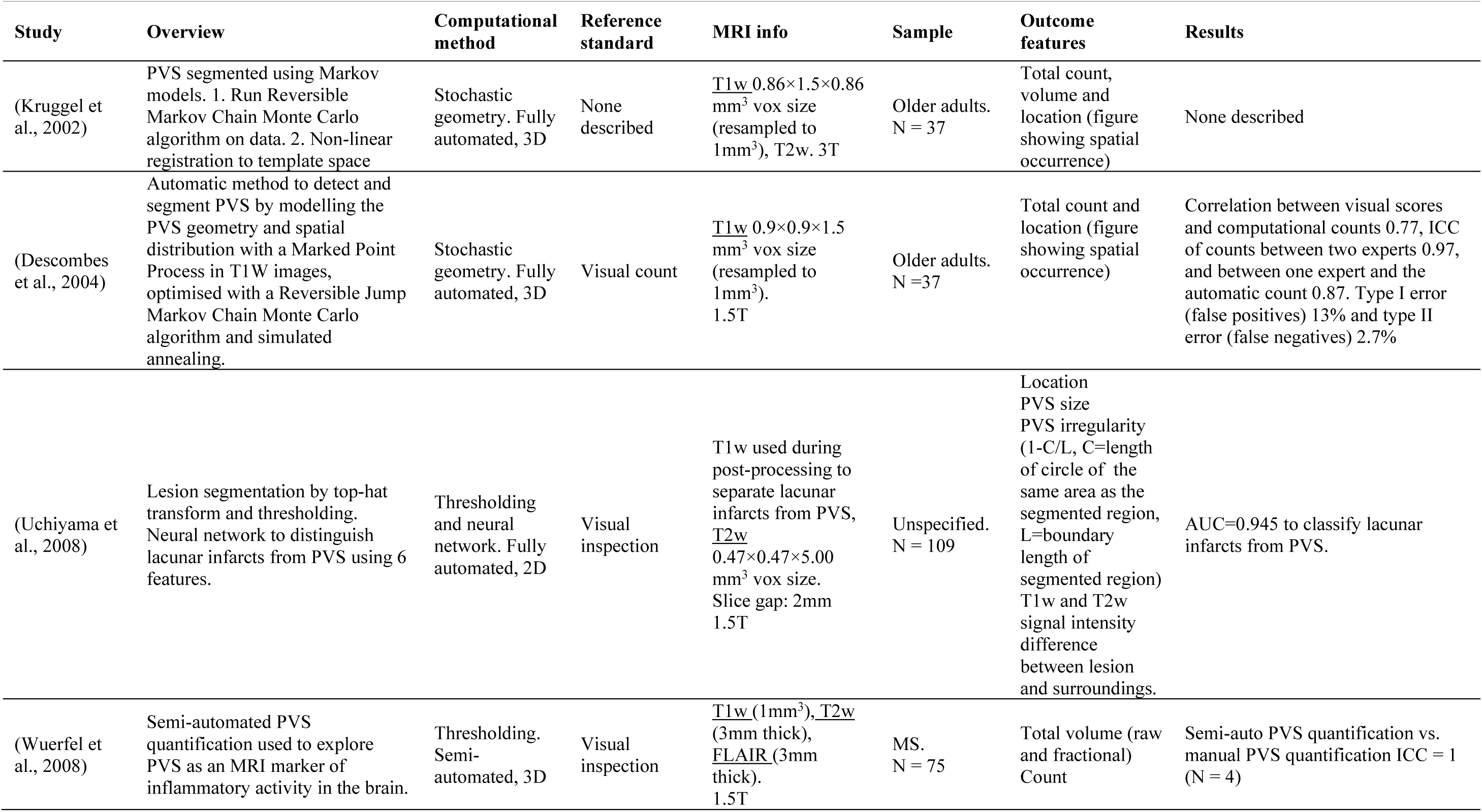
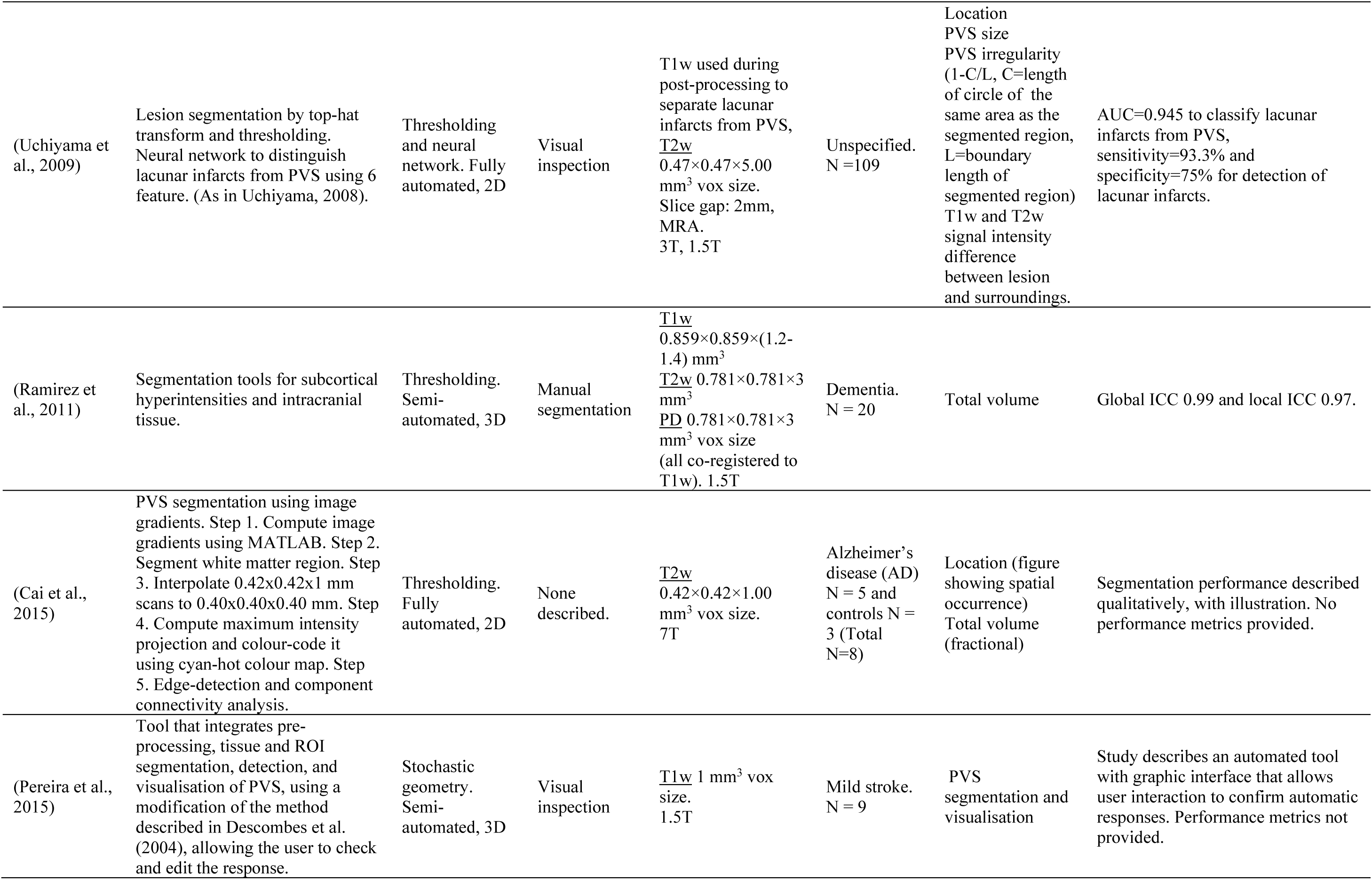
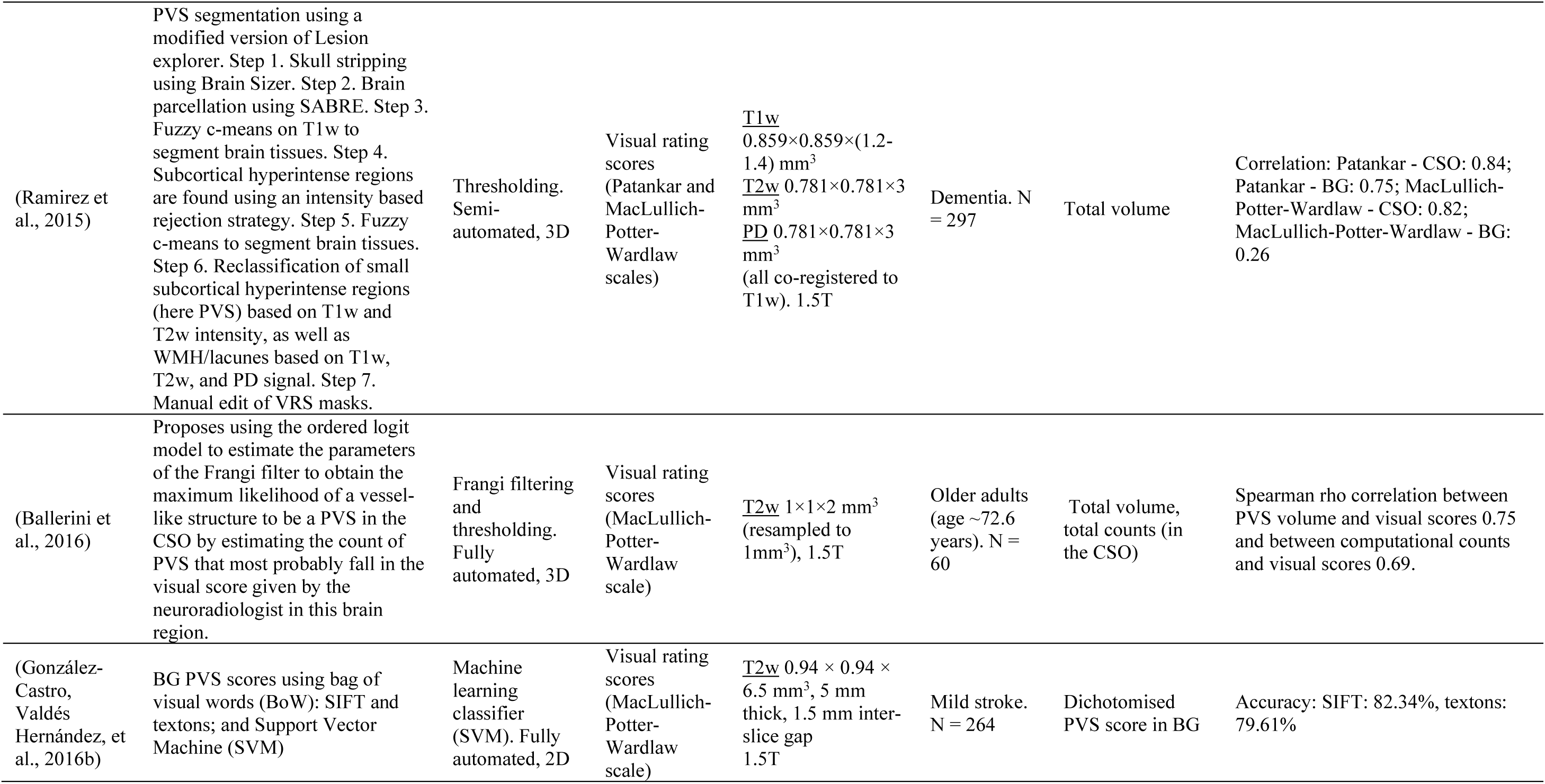
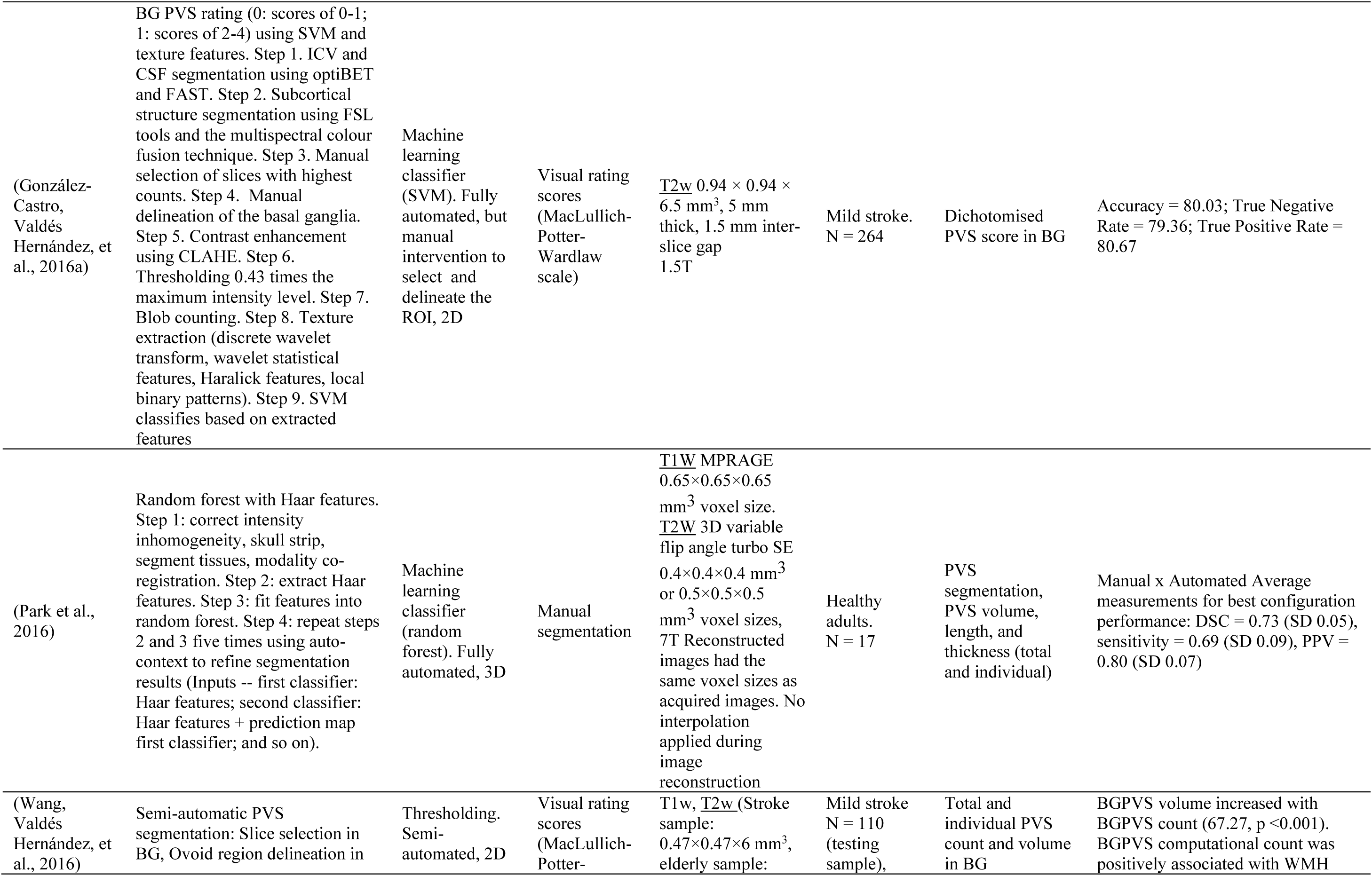
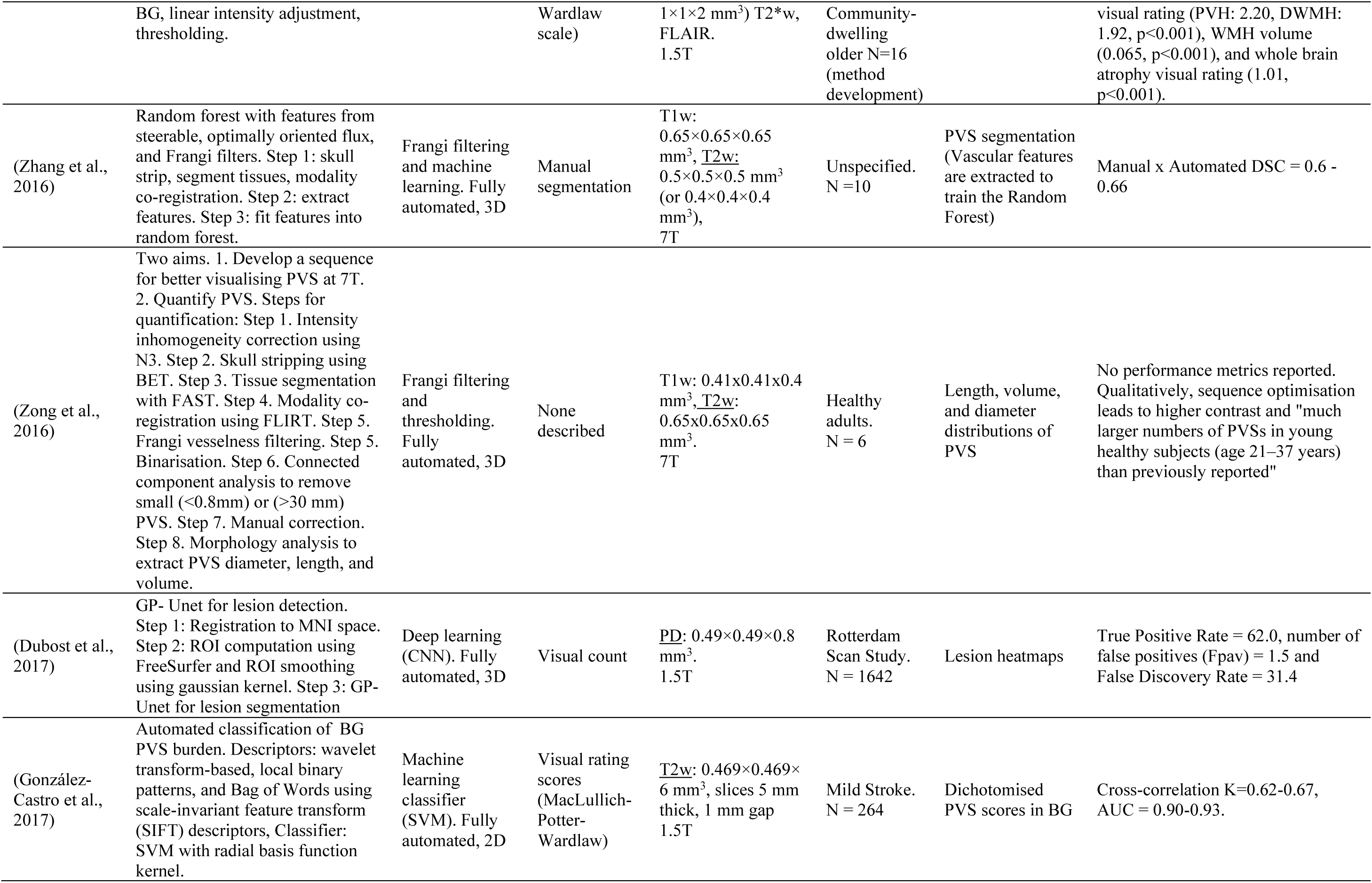
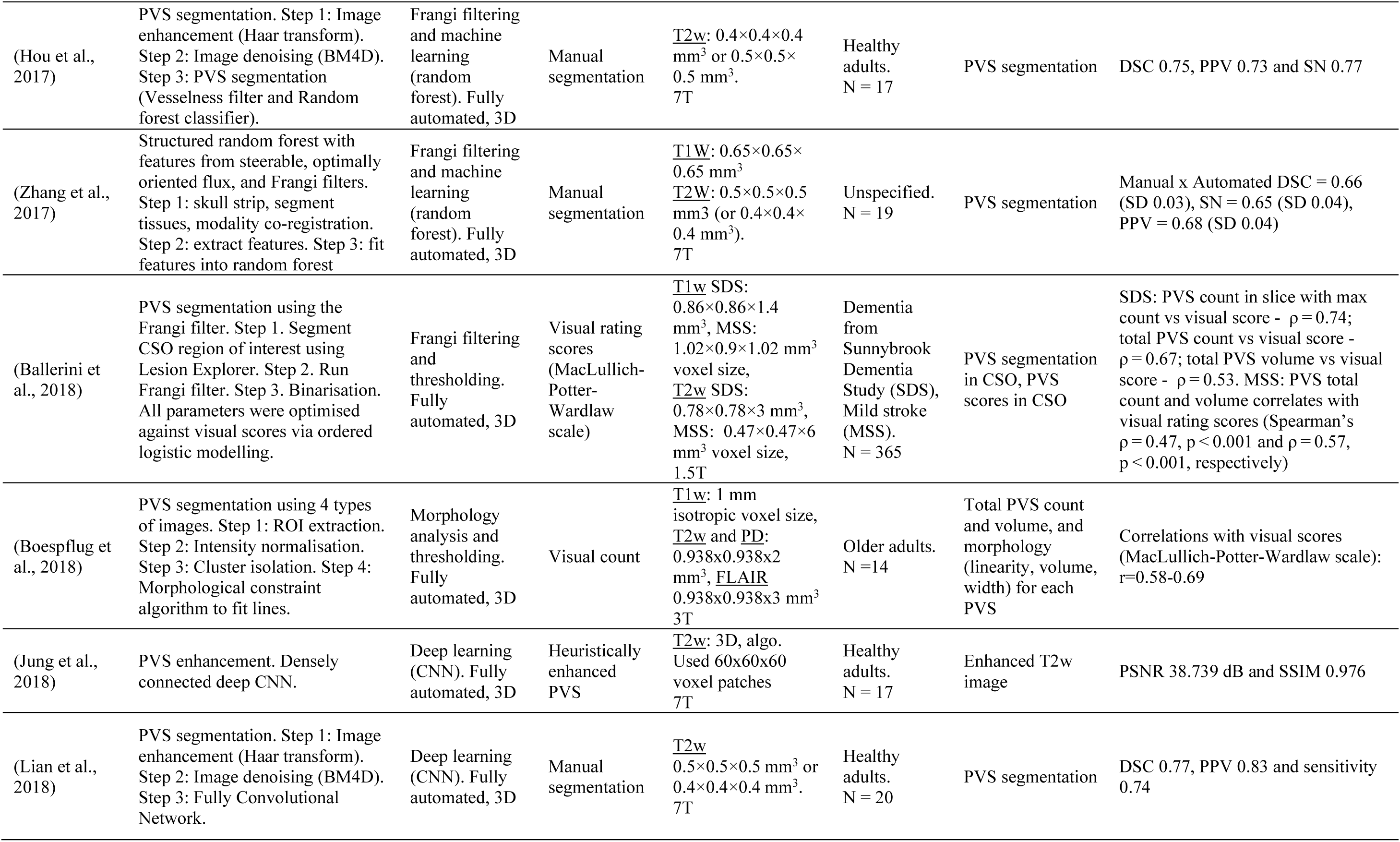
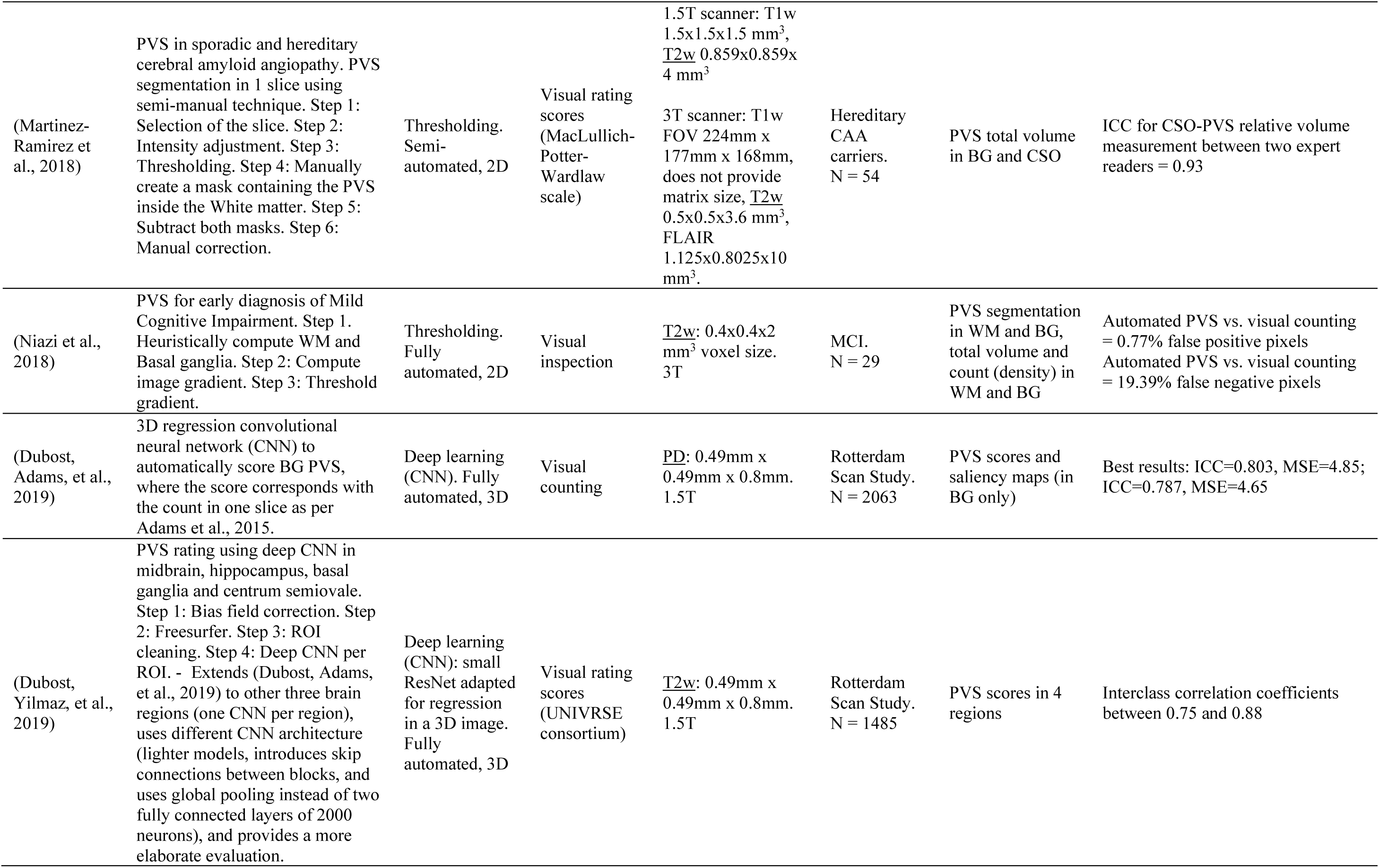
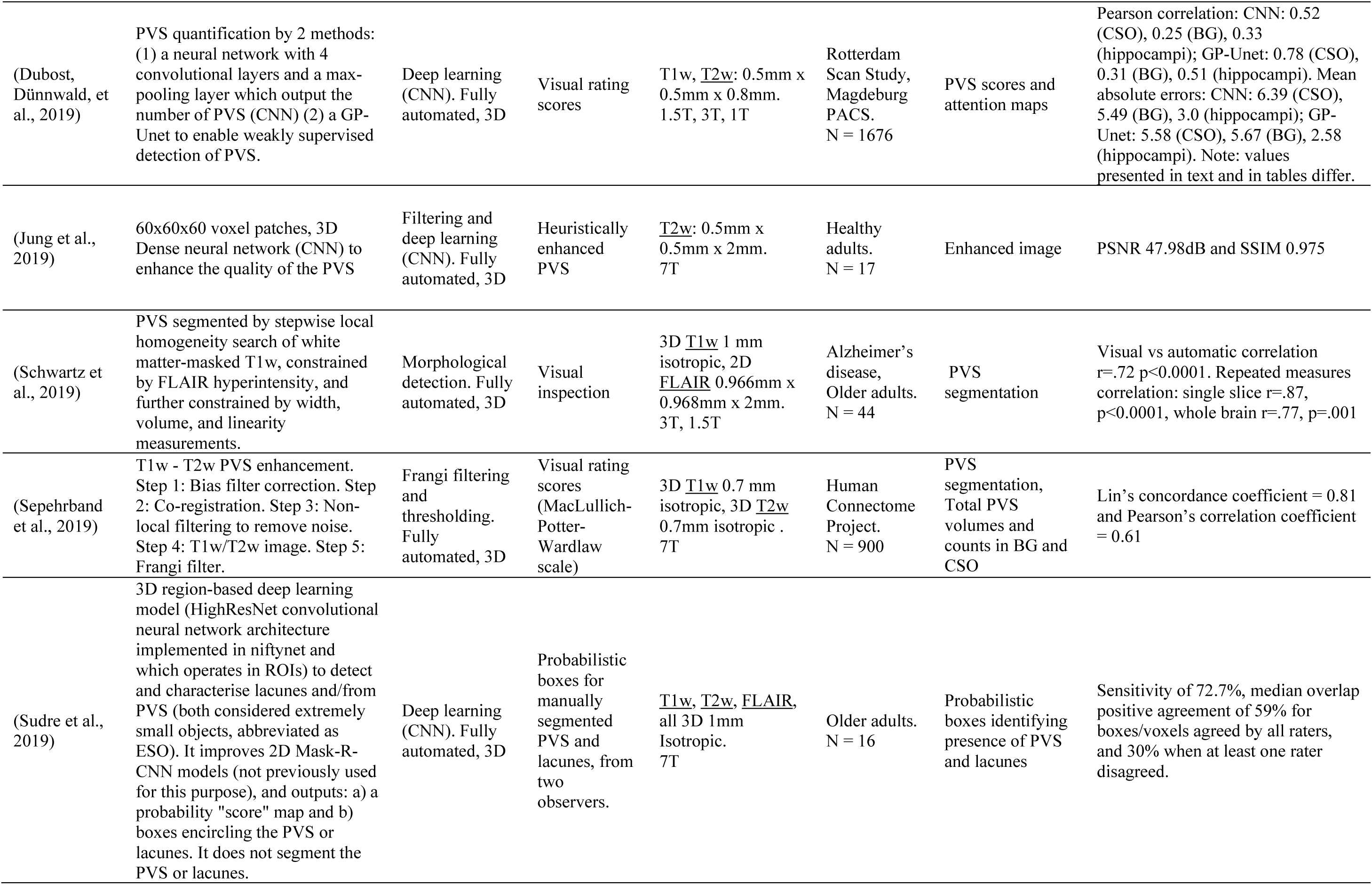
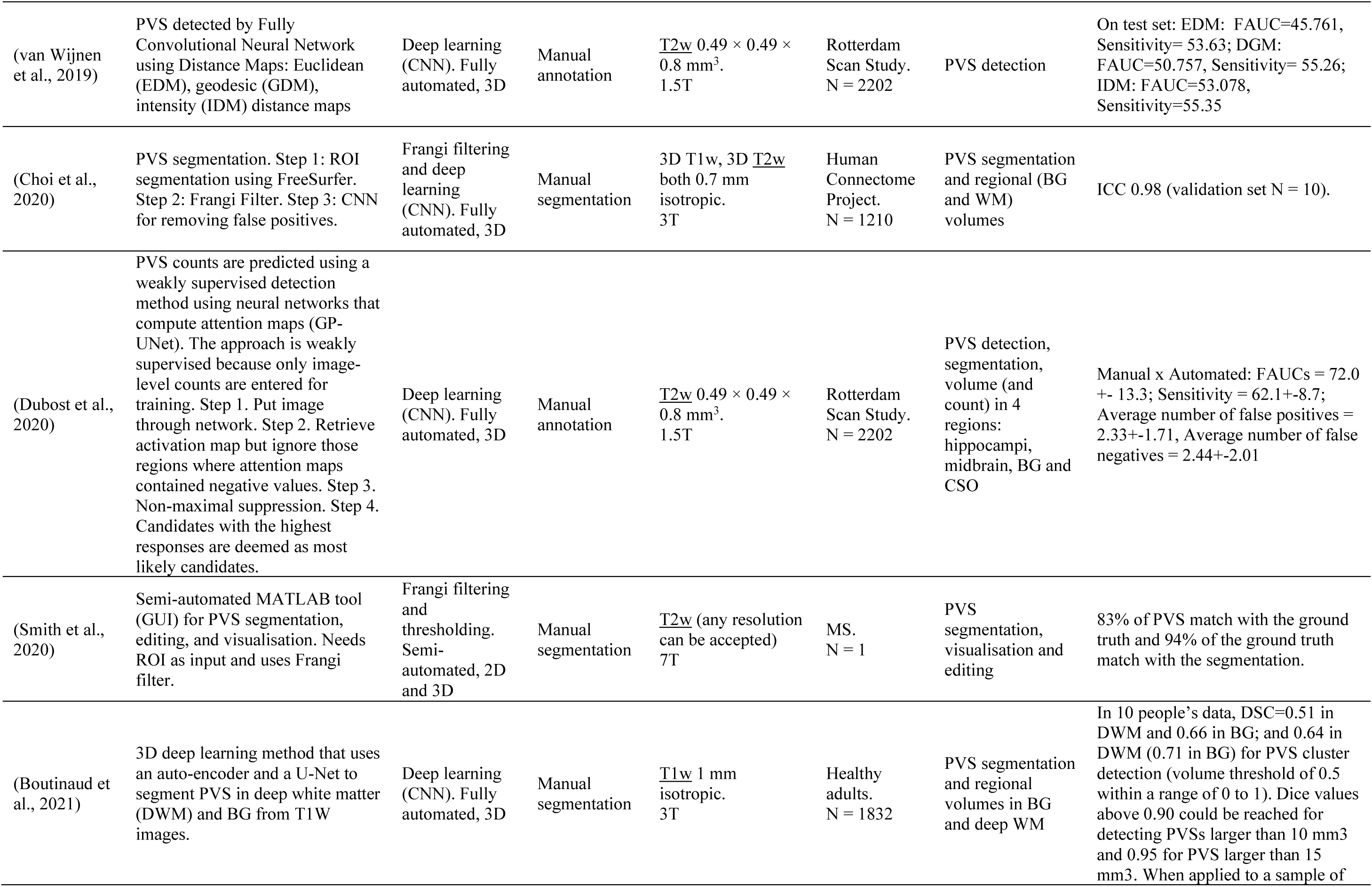
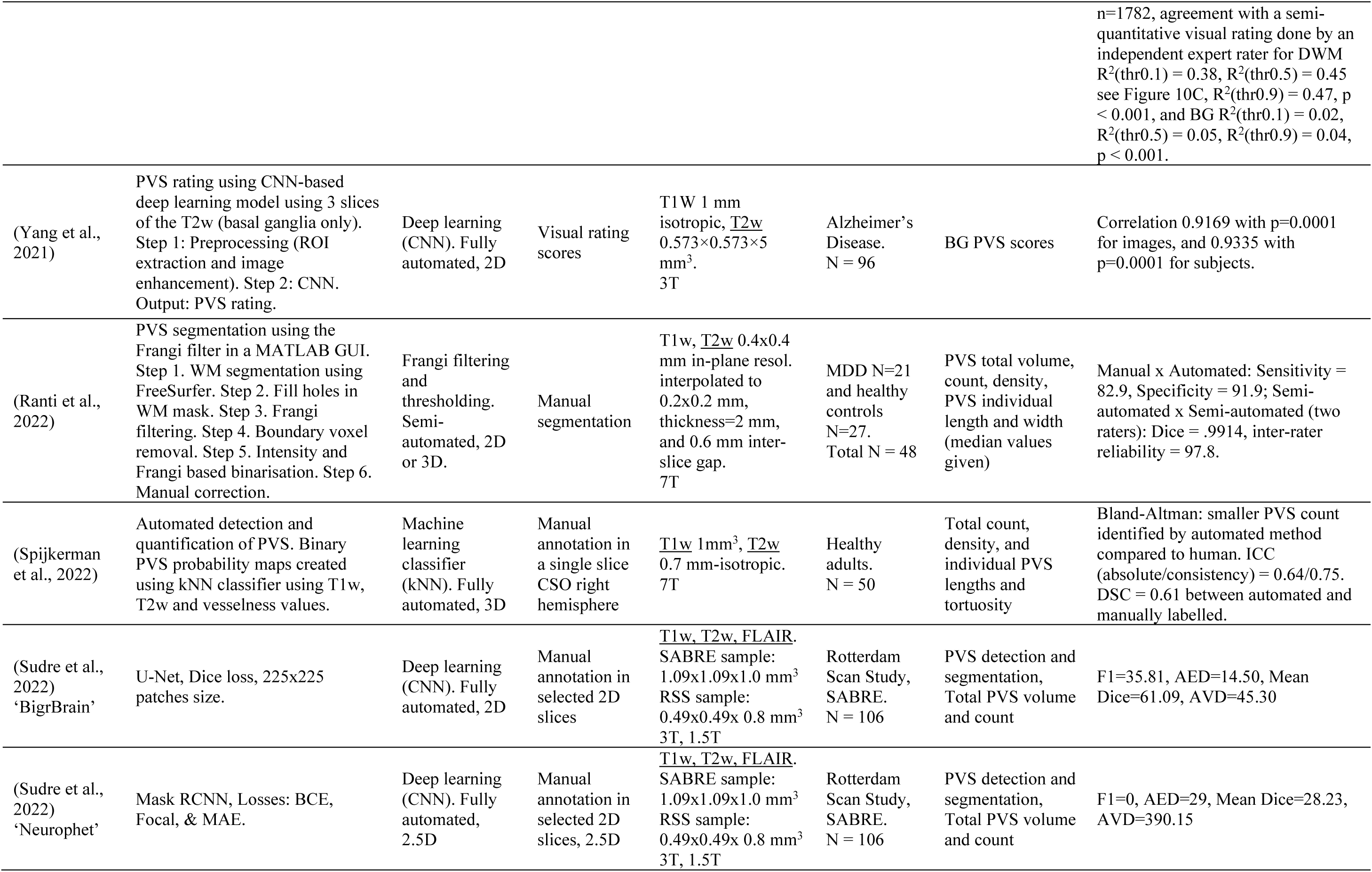
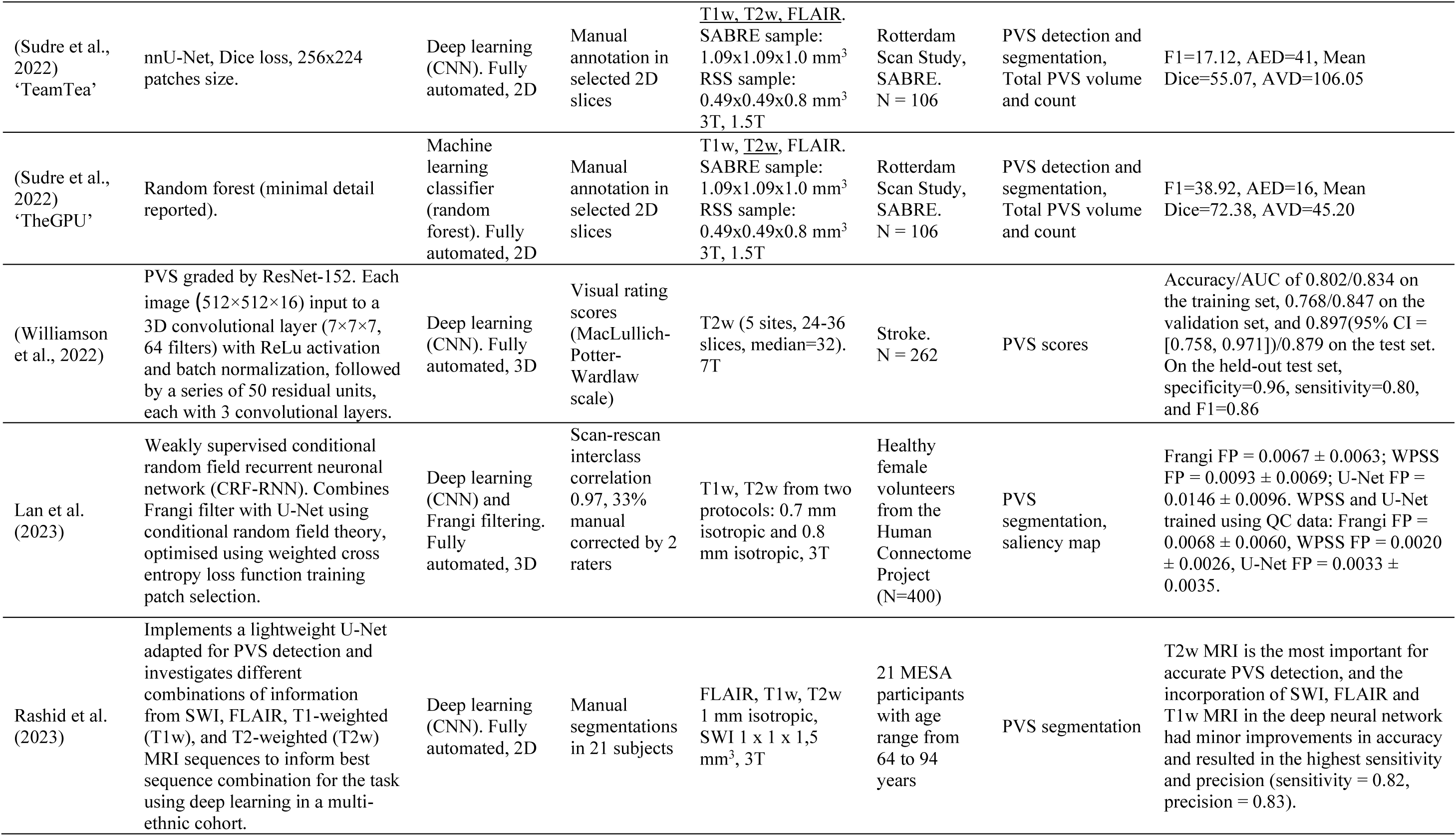
Method development study summaries Note: The MRI sequence(s) underlined is(are) the one(s) used specifically to generate the PVS assessments. The non-underlined MRI sequences in each case are used in pre- /post-processing. Legend: PVS: Perivascular Spaces, AUC: Area Under the (ROC) Curve, FAUC: F stat (ratio of two variances) of the AUC from two measurements (e.g. manual vs. automatic) CI: Confidence Interval, CNN: Convolutional Neural Networks, GUI: Graphic Unit Interface, AED: Absolute Error Difference, ICC: Intra-class Correlation Coefficient, AVD: Absolute Volume Difference, DSC: Dice Similarity Coefficient, BG: basal ganglia, CSO: centrum semiovale, FOV: Field Of View, CLAHE: Contrast Limited Adaptive Histogram Equalization, FP: False Positives, SWI: Susceptibility Weighted Images, MDD: Major Depressive Disorder, WM: White Matter, GM: Grey Matter, DWM: Deep White Matter, PSNR: Peak Signal-to-Noise Ratio, SSIM: Structural Similarity Index Metric, PPV: Positive Predictive Value, SABRE: Southall And BRent Revisited study, MESA: Multi-Ethnic Study of Atherosclerosis, WPSS: Weakly Supervised Perivascular Spaces Segmentation, PVH: periventricular hyperintensities, DWMH: deep white matter hyperintensities, MSE: Mean Squared Error, MAE: Mean Absolute Error, F1: accuracy metric that combines the precision and recall scores of a model by computing how many times a model made a correct prediction across the entire dataset, QC: Quality Control, TPR: True Positive Rate, FDR: False Discovery Rate, CNN: Convolutional Neural Network, SIFT: Scale Invariant Feature Transform, SD: Standard Deviation.

**Table 7.** Types of computational approaches *Legend: N: number of sources, %: Percentage with respect to the 48 sources in the Method development group*

| Table 7. Types of computational approaches |  |  |
| --- | --- | --- |
|  | <u>N</u> | <u>%</u> |
| <u>Classical image processing</u> |  |  |
| Intensity thresholding | 7 | 14.58 |
| Frangi filtering + Thresholding | 6 | 12.50 |
| <u>Machine learning</u> |  |  |
| Stochastic geometry | 3 | 6.25 |
| Support Vector Machine (SVM) | 3 | 6.25 |
| Random Forest (RF) | 2 | 4.17 |
| Convolutional Neural Networks (CNN) | 16 | 33.33 |
| K-Nearest Neighbours (KNN) | 1 | 2.08 |
| <u>Classical image processing + Machine learning</u> |  |  |
| Intensity thresholding + Neural network | 2 | 4.17 |
| Frangi filtering + RF | 3 | 6.25 |
| Filtering + CNN | 1 | 2.08 |
| PVS Morphological detection (width, length, size) | 2 | 4.17 |
| Frangi filtering + CNN | 2 | 4.17 |

**Table 8.** Computational PVS quantification improvement study summaries Legend: PVS: perivascular spaces, T1w: T1-weighted MRI sequence, T2w: T2-1weighted MRI sequence, SWI: Susceptibility Weighted Imaging, FLAIR: Fluid-Attenuated Inversion Recovery MRI sequence, SD: Standard Deviation, RORPO: Ranking Orientation Responses of Path Operators, IQR: Inter-Quartile Range, BG: basal ganglia region, CSO: centrum semiovale region, PVS-DRO: PVS Digital Reference Object, MSE: Mean Squared Error

| Study | Overview | Approach | Reference standard | MRI sequences | Sample N | Result |
| --- | --- | --- | --- | --- | --- | --- |
| (Paz et al., 2009) | Determines level of compression that allows for the detection of small lesions (including PVS) | Mathematical observer models | Visual assessment | T2w. Unspecified | MS. N = 10 | The blurring effect introduced by the JPEG 2000 compression technology does not jeopardize the lesion detection task when using bit rates higher than 0.125 bpp. |
| (Bernal et al., 2020) | Framework for improving PVS segmentation by analysing image texture (i.e., energy, contrast, correlation, homogeneity, entropy and variance) and filtering when required (i.e., if the texture analysis suggest the image is noisy), using a total variation filter. | Frangi filtering and thresholding | Visual rating scores | T2w. 3T | Mild stroke. N = 60 | Combination of all 6 textural features improved classification in noisy vs. less noisy/clean (AUC=88.8%), R2 values were approximately 13% and 40% for count and volume of PVS in the basal ganglia in original scans, and increased to 30% and 47% for the same measurements when filtered. In the CSO, the improvement in R2 values was from 0.73% to 11% for counts and from 7.55% to 35% for volume. Overall, the scheme proposed improves the PVS quantification, but it is unclear if can be extended to other protocols/ images from other scans. |
| (Bernal et al., 2021) | Explores motion artefact reduction. PVS segmented using Frangi filter. Step 1: quality assessment. Step 2: if images are of bad quality (contain motion artefacts), filter them by manually removing segments of k-space displaying errors. Step 3: filter images with Frangi filter. Step 4: threshold. | Frangi filtering and thresholding | Visual rating scores | T2w. 3T | Mild stroke. N = 60 | Visual x Automated Counts: pBG = 0.50, pCSO=0.34; Visual x Automated Volume: pBG = 0.72, pCSO = 0.86. This proof-of-concept paper shows that processing images depending on their quality leads to improved results. |
| (Zong et al., 2021) | Determines effect of motion artefacts and motion correction of PVS quantification. | Deep learning (CNN) | Data without motion artefacts | T2w. 7T | Healthy adults. N = 39 | Without motion correction (MC), VF, and count decreased significantly with increasing head rotation. MC improved PVSV visualization in all cases with severe motion artefacts. While both raters agreed on the improvements of PVSV visibility by MC in all cases with severe motion artefacts, the agreement was only 43% in other cases. MC decreased diameter in white matter (WM) and increased VF, count, and contrast in basal ganglia and WM. The changes of VF, count, and contrast after MC strongly correlated with motion severity. MC eliminated the significant dependences of VF and count on rotation and reduced the inter-subject variations of VF and count. |
| (Bernal et al., 2022) | Compares different vesselness filtering methods with synthetic data and shows quantification problems. | Frangi, Jerman, RORPO filtering and thresholding | T2w-like images with synthetically generated PVS-like tubular structures (i.e., cylinders) | T2w-like synthesised data | Synthetic data | RORPO best when voxels isotropic. Performance of all filters affected by image quality. Filters unable to distinguish PVS from hyperintense structures. Area under precision-recall curve dropped substantially (Frangi: from 94.21 [IQR 91.60, 96.16] to 43.76 [IQR 25.19, 63.38]; Jerman: from 94.51 [IQR 91.90, 95.37] to 58.00 [IQR 35.68, 64.87]; RORPO: from 98.72 [IQR 95.37, 98.96] to 71.87 [IQR 57.21, 76.63] without and with other hyperintense structures, respectively. |
| (Duarte Coello et al., 2024) | Uses a purpose-built digital reference object to construct an in-silico phantom for establishing the limits of validity of PVS quantification, and validates it using a physical phantom. Uses cylinders of different sizes as models for PVS. Also evaluates the influence of 'PVS' orientation, and different sets of parameters of the two vesselness filters that have been used for enhancing tubular structures, namely Frangi and RORPO filters, in the measurements' accuracy. | Frangi or RORPO filtering and thresholding. | Ideal values, physically measured using a hole gage and a calliper in the physical phantom, or derived from the cylinder equation in the in-silico phantom | T2w contrast using a turbo spin-echo sequence (Physical phantom imaged at 7T) | Synthetic data | PVS measurements in MRI are only a proxy of their true dimensions, as the boundaries of their representation are consistently overestimated. The success in the use of the Frangi filter relies on a careful tuning of several parameters. Alpha= 0.5, beta= 0.5 and c= 500 yielded the best results. RORPO does not have these requirements and allows detecting smaller cylinders in their entirety more consistently in the absence of noise and confounding artefacts. The Frangi filter seems to be best suited for voxel sizes equal or larger than 0.4 mm-isotropic and cylinders larger than 1 mm diameter and 2 mm length. 'PVS' orientation did not affect measurements in data with isotropic voxels. |
| (Duarte Coello et al., 2023) | Curve approximation for accurate estimation of PVS morphometrics: length and diameter, out from PVS segmentations. | Approximation of the connected component to a Bezier curve | Ideal values, derived from counting the voxels in a binarised curve, derived from a PVS-DRO with voxel size of $0.35 \times 0.35 \times 0.35$ mm<sup>3</sup> | Not applicable | Not applicable | The Bézier curve approximation performs better than the ellipsoid, being closer to the ideal measurements. For diameter: MSE Bezier=0.26mm, MSE ellipsoid=7.74mm, For length: MSE Bezier=0.051mm, MSE ellipsoid=10.34mm |
| (Valdés Hernández et al., 2024) | Evaluates modifications and alternatives to a state-of-the-art PVS segmentation method that uses a vesselness filter to enhance PVS discrimination, followed by thresholding of its response, applied to brain magnetic resonance images (MRI) from patients with sporadic small vessel disease acquired at 3T. | Frangi or Jerman filtering and thresholding. | Visual rating scores (McLulich-Potter-Wardlaw), and visual inspection | T1w (1 mm isotropic), T2w (0.90 mm × 0.937 mm × 0.937 mm), 3T (SWI and FLAIR are used for visual assessment of the results) | Mild stroke N=228.<br>(mean age (SD): 65.77 (11.17) years, 34% female) | The method is robust against inter-observer differences in threshold selection, but separate thresholds for each region of interest (i.e., BG, CSO, and midbrain) are required. Noise needs to be assessed prior to selecting these thresholds, as effect of noise and imaging artefacts can be mitigated with a careful optimisation of these thresholds. PVS segmentation from T1w images alone, misses small PVS, therefore, underestimates PVS count, may overestimate individual PVS volume especially in the basal ganglia, and is susceptible to the inclusion of calcified vessels and mineral deposits. Visual analyses indicated the incomplete and fragmented detection of long and thin PVS as the primary cause of errors, with the Frangi filter coping better than the Jerman filter. |

From all the sources in this category, only nine provide their code in a public repository, 28 do not provide the code, and one provides via to request the code (Supplementary Table 2). The rest use publicly available resources like the Frangi vesselness filter implementation by Dirk-Jan Kroon (2009) hosted by Matlab Central File Exchange (https://www.mathworks.com/matlabcentral/fileexchange/24409-hessian-based-frangi-vesselness-filter), or the 3D multi-radii optimally oriented flux responses for curvilinear structure analysis implementation by Max W.K. Law (2013) also hosted by Matlab Central File Exchange (https://www.mathworks.com/matlabcentral/fileexchange/41612-optimally-oriented-flux-oof-for-3d-curvilinear-structure). The FMRIB Software Library (FSL, https://fsl.fmrib.ox.ac.uk/fsl/fslwiki) is the preferred software used for sequence co-registration, occasionally for brain extraction, and in two sources (Gonzalez-Castro et al., 2016b, 2017) it is also used for generating priors of the basal ganglia region. AFNI (https://afni.nimh.nih.gov/) is used by two sources (Boespflug et al., 2018; Williamson et al., 2022) for co-registration and skull-stripping, and elastix (https://elastix.lumc.nl/) is used for co-registration by one (Spijkerman et al., 2022). Not all methods correct for b1 inhomogeneities in the magnetic field, compensate for the presence of noise, or refer to a software/algorithm for normalising the intensities of the images prior to segmentation. The software most commonly used for generating the regions of interest is Freesurfer (https://surfer.nmr.mgh.harvard.edu/) (See Supplementary Table 2).

#### 3.3.2. Model training

Thirty-two (66.68%) of the 48 quantification methods developed required training. Training and validation approaches included ‘leave one out’ (5 studies, 10.42%), ‘k-fold cross-validation’ (7 studies, 14.58%) and variations involving training data in one subset of data and testing in another (14 studies, 29.17%), training in one subset, validating in another, and testing in a further subset (4 studies, 8.33%), and a progressive/iterative training including altering thresholds and increasing sample sizes (2 studies, 4.17%).

#### 3.3.3. Reference standard

The reference standard, or ‘ground truth’, most frequently used in PVS quantification method development was visual rating scores (13 studies, 27.08%). Other visual reference standards included visual count of PVS (5 studies, 10.42%) and ‘visual inspection’ (often lacking further detail of what this entailed, but where mentioned included differentiating PVS from lacunes; 5 studies, 10.42%).

Manual segmentation was used in 11 (22.92%) studies, and manual annotation in eight (16.67%). Three studies (6.25%) did not describe a ground truth or reference standard, and a further 3 studies (6.25%) used more idiosyncratic reference standards (e.g., heuristically enhanced PVS, and probabilistic boxes for previously manually segmented PVS).

#### 3.3.4. Populations and sample sizes

A brief overview of populations and sample sizes across all study types is provided in section 3.2.2, Tables 1, 2 and 3. The most frequent age group in which computational PVS quantification methods have been developed was in mid- and older adults (participants aged 40 years and over; 11 studies, 22.92%), followed by general adult (18 years and over; 10 studies, 20.83%), and older adult (60 years and over; 10 studies, 20.83%). Non-clinical populations were more widely represented than clinical (27 studies, 56.25% non-clinical, 17 studies, 35.42% clinical). Of the method development studies that used clinical populations, medical conditions within these populations included hereditary conditions (one study involving hereditary cerebral amyloid angiopathy carriers; 2.08%), six studies involving stroke patients (i.e., cerebrovascular; 12.5%), nine studies with samples with neurological or neurodegenerative diseases (e.g., Alzheimer’s disease, multiple sclerosis; 18.75%), and one study involving patients with major depressive disorder; 2.08%). However, there is considerable overlap between the samples used by the different studies. Figure 3 comparatively illustrates the distribution of sample sizes of the studies in this category. Dubost et al. (2017, 2019a, 2019b, 2019c, 2020) and van Wijnen et al. (2019) use more than 1000 scans from the Rotterdam Scan Study. A manually annotated subset of it is used in the four studies described by Sudre et al. (2022). Ballerini et al. (2018) and Ramirez et al. (2011, 2015) use data from the Sunnybrook Dementia Study. Gonzalez et al. (2016a, 2016b, 2017), Wang et al. (2016), and Ballerini et al. (2018) all use scans from the Edinburgh Mild Stroke Studies. Other common data sources used are the Human Connectome Project (Shepehrband et al., 2019; Choi et al., 2020; Lan et al., 2023), the Lothian Birth Cohort 1936 (Ballerini et al., 2016, Wang et al., 2016), the Multi-Ethnic Study of Atherosclerosis (MESA) cohort (Rashid et al., 2023) and the Southall And BRent REvisited (SABRE) (Sudre et al., 2022, 4 participating studies). All these are well-known clinical and population studies with data available either by request to the data holders or through public databases, which facilitates comparability in the results. Studies using small or not publicly available samples (22 studies), or which did not specify the source of data used (4 studies) made up 54.17% of the total number of studies.

**Figure 3.**
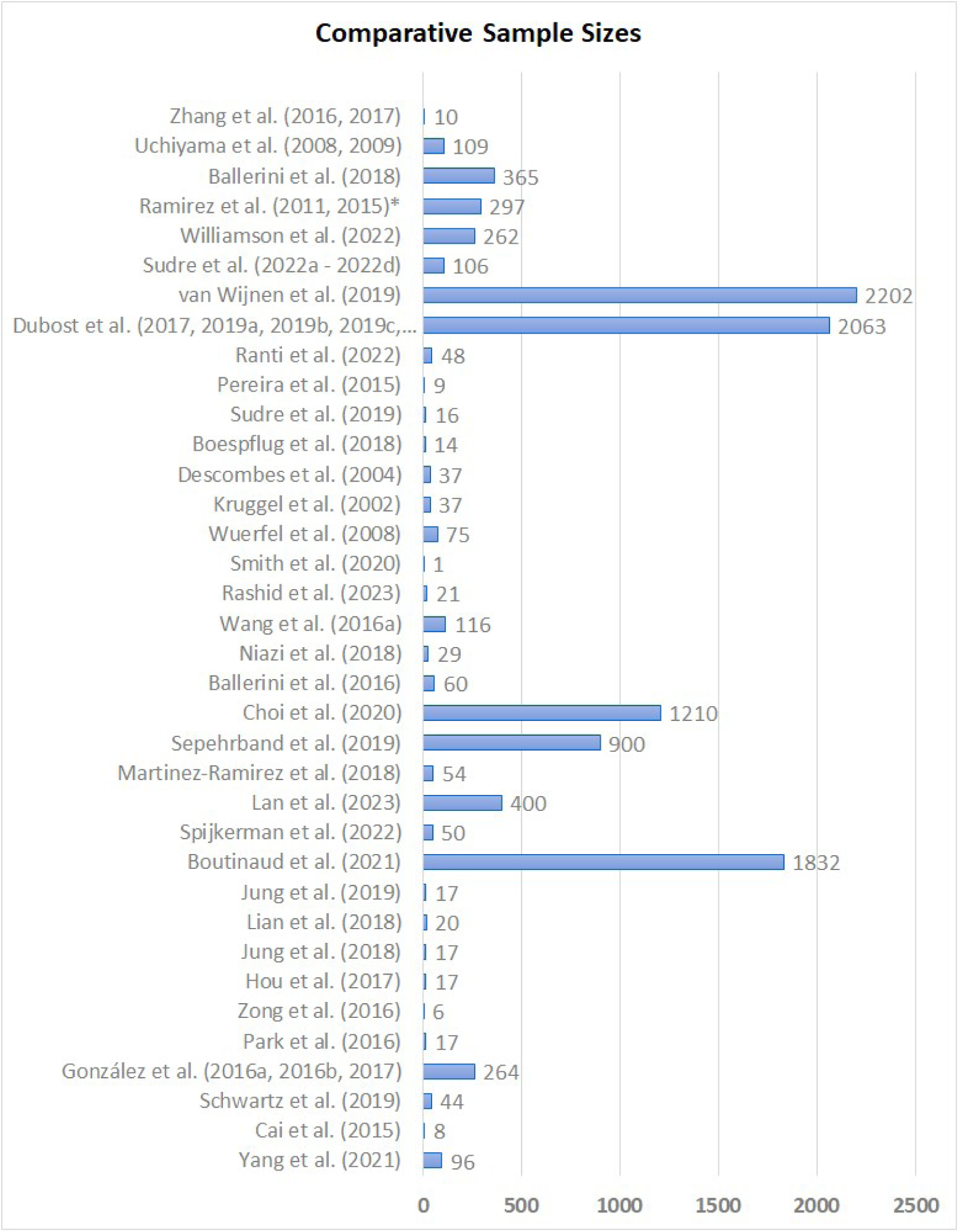
Comparative sample sizes of the studies in the method development category. Dubost et al. and Ramirez et al. used overlapped subsamples from the same data sources. For these cases, the largest sample is represented in the graph.

#### 3.3.6. Assessing computational PVS quantification

A number of different approaches to assessing the quality of PVS quantification were used, making direct comparisons difficult. More commonly used assessments include measures of associations between computational output and reference standard (e.g., correlations), measures of accuracy (e.g., false positive/false negatives, ROC analyses), and measures of spatial agreement. Eighteen studies calculated association (correlation, regression) with reference standard measurements or scores (Table 6), with best performance (manual vs. computational approach) plotted in Figure 4. Best results were achieved with semi-automated thresholding methods (Wuerfel et al., 2008; Ramirez et al., 2011), and by the approach presented by Choi et al. (2020) that uses a CNN to remove false positives from segmentation priors derived from thresholding the output from the Frangi filter, although this was only validated in 10 scans. The CNN schemes proposed by Dubost et al. (2019a) and Yang et al. (2021) followed in order of good performance. The highest variability in the method’s performance was achieved with the filtering approach proposed by Boespflug et al. (2018).

**Figure 4.**
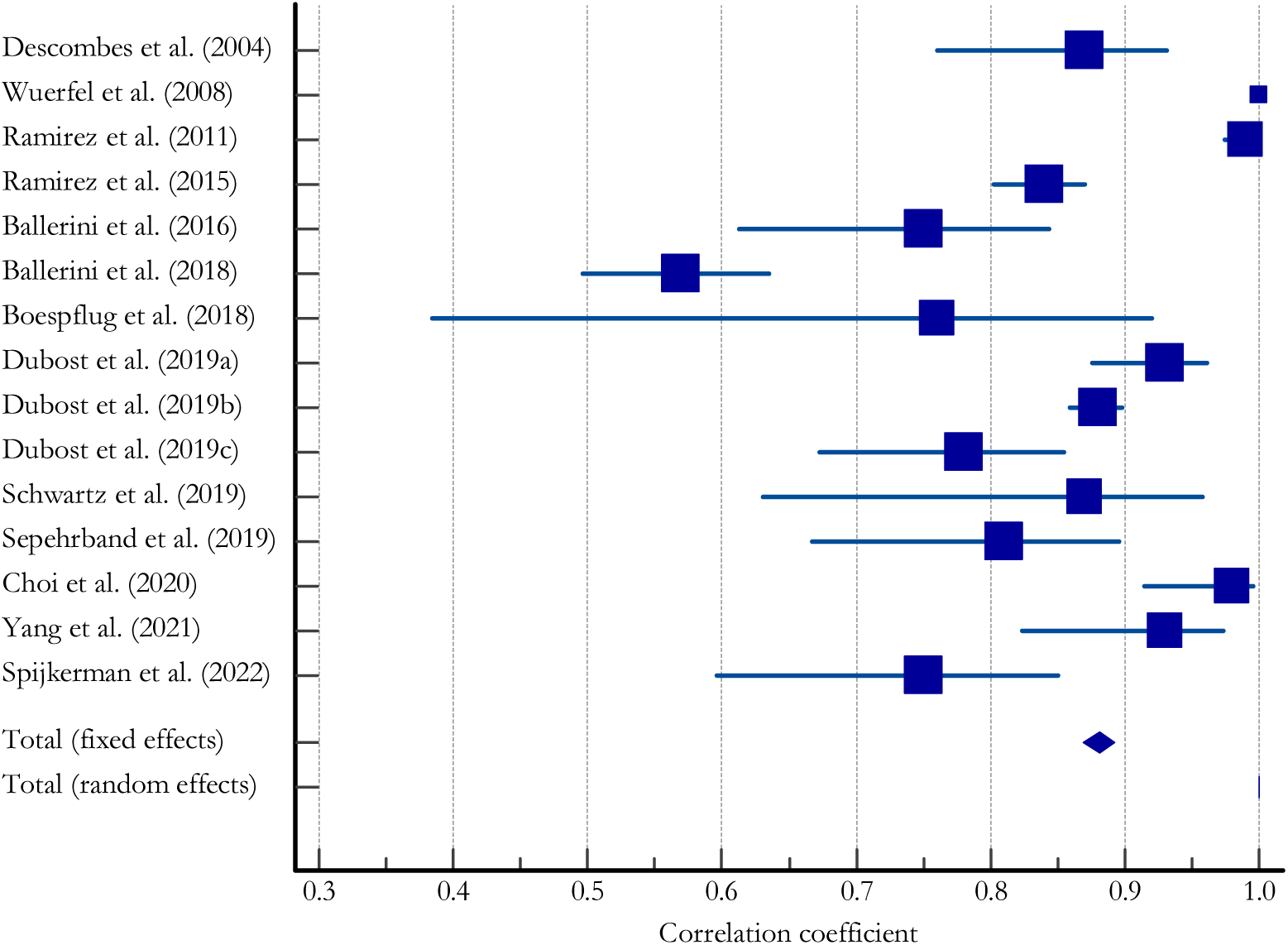
Forest plot of correlation coefficients of best performance (non-computational vs. computational) PVS quantification (MedCalc (https://www.medcalc.org/)). Square size indicates relative sample size, diamond indicates pooled fixed and random effects correlation coefficient.

Accuracy metrics varied, largely depending on the reference standard, but all methods reporting accuracy results used various metrics (Table 6 and Supplementary Table 3). Average Dice similarity coefficient (DSC) values from sources that report them are illustrated in Figure 5. The majority are between 0.6 and 0.8. Only Ranti et al. (2022) stands out with a value of 0.9914 (Table 6, Figure 5).

**Figure 5.**
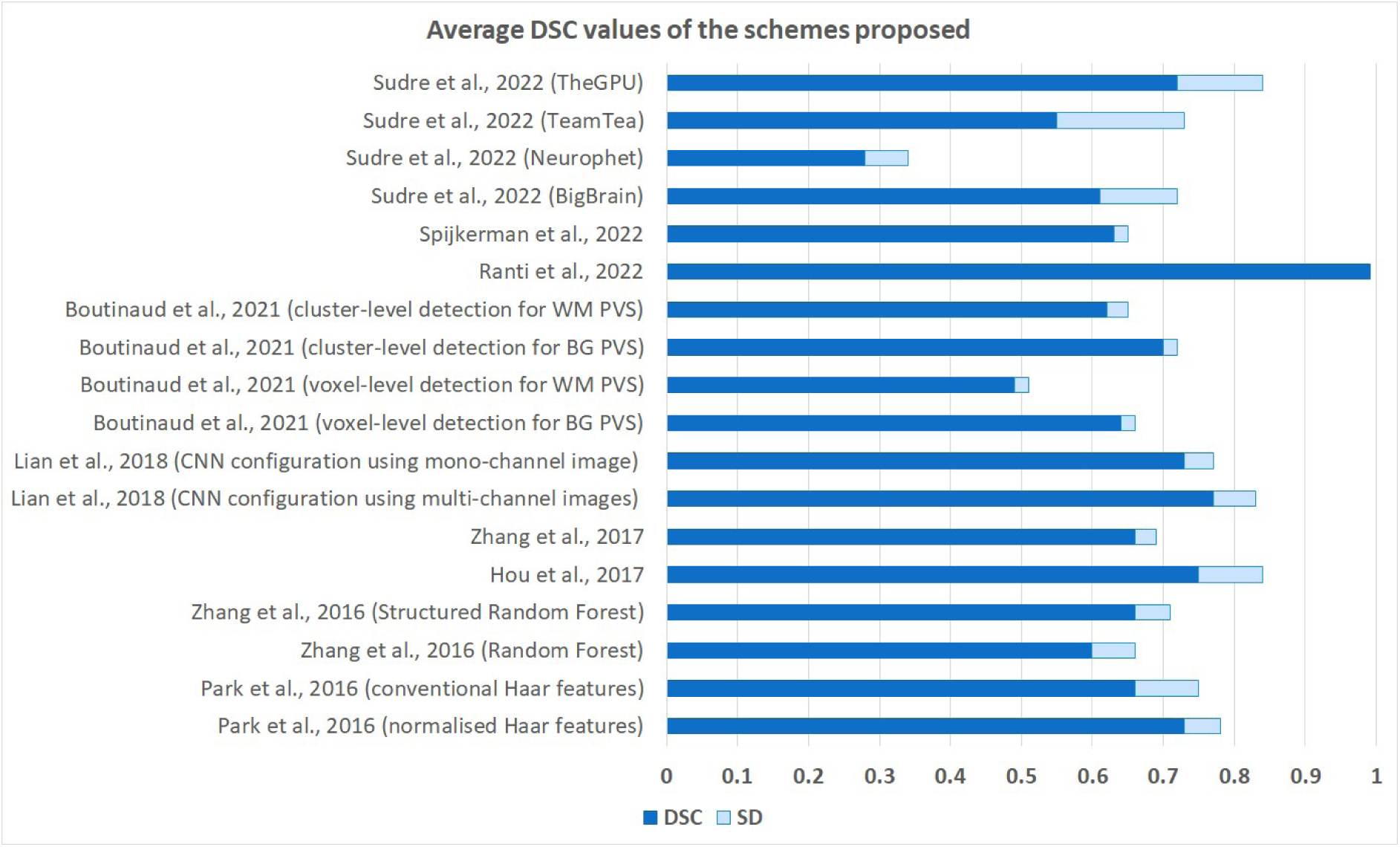
Average DSC and standard deviations (interquartile range for the schemes proposed in Sudre et al., 2022) results for each of the schemes proposed.

### 3.4. Improvement studies

Summaries of all improvement studies identified, which fitted our inclusion criteria (N = 8), are presented in Table 8. Targets for quantification improvement included image compression, motion artefacts and noise in general, image texture, comparison of vessel likelihood filters, and any source from which to derive useful recommendations for improving any of the methods developed up to date. Three sources are chapters in conference proceeding books (Bernal et al., 2020, 2021; Duarte Coello et al., 2023). The rest are journal publications.

#### 3.4.1. Computational approach

Five out of the eight studies in this category utilised Frangi filtering and thresholding approaches (with three also assessing RORPO and/or Jerman filters). One study used a Deep Learning (CNN) approach (Zong et al., 2021), and two used mathematical observer models (Paz et al., 2009; Duarte Coello et al., 2023).

#### 3.4.2. Reference standards

Three of the eight studies used visual rating scores (Bernal et al., 2020, 2021, Valdés Hernández et al., 2024), two used visual assessment of the results (Paz et al., 2009; Valdés Hernández et al., 2024), one used data without motion artefacts (Zong et al., 2021), and three used measurements of synthetically-generated cylindrical structures (Bernal et al., 2022, Duarte Coello et al., 2023, 2024) as reference standards.

#### 3.4.3. MR sequences and field strengths

All studies aimed at improving the detectability and segmentation accuracy of PVS used T2w imaging, or synthesised T2w-like images in one case, as this is the preferred sequence to identify PVS as per STRIVE recommendations (Wardlaw et al., 2013). Three studies used 3T images, two used 7T images, and one study did not specify field strength. The study that used T1w images (Valdés Hernández et al., 2024) found inconsistencies in the results compared with those from T2w, and suggested these might reflect presence of vessel mineralisation, hypointense, as PVS, in T1w MRI sequence.

#### 3.4.4. Populations and sample sizes

Populations used in improvement studies included clinical groups (multiple sclerosis, mild stroke), and healthy adults. Three studies used synthetic data. Sample sizes ranged from N = 10 to N = 228 participants, although one study (Bernal et al., 2022) used intensity values from T2w images from 700 community-dwelling older individuals, in addition to WMH probability distribution maps derived from 47 patients with systemic lupus erythematosus to generate the Digital Reference Object used for the evaluations.

#### 3.4.5. Improvement study results

Each of the eight improvement studies had different targets (Table 8). One study examined image compression (Paz et al., 2009), finding that JPEG 2000 compression does not jeopardise PVS detectability when bit rates are greater than 0.125bpp. One study proposes a framework for PVS quantification by analysing image texture features and applying a total variation filter if necessary, to improve PVS quantification in noisy versus less noisy or clean imaging (AUC = 88.8%) (Bernal et al., 2020). Three studies examined the impact of motion artefacts on PVS quantification, finding that correcting motion artefacts improved PVS quantification when using Frangi filtering plus thresholding (Bernal et al., 2021) and when using a convolutional neural network approach (Zong et al., 2021), or that simply using higher thresholds after filtering for noisy images compared with the thresholds used for clean images will be enough (Valdés Hernández et al., 2024). Comparison of the performance of Frangi, RORPO and Jerman filters for highlighting PVS candidates was the subject of three studies. One study compared the three filtering approaches and found that RORPO performed best in isotropic imaging, but that all filter types were unable to distinguish between PVS and other hyperintense features (Bernal et al., 2022). Another compared Frangi with Jerman and found more incomplete and fragmented detection of long and thin PVS with Jerman than with Frangi (Valdés Hernández et al., 2024). Another compared the performance of RORPO and Frangi in phantoms to conclude that RORPO, in addition of having less adjustment requirements, detected smaller cylinders in their entirety more consistently while Frangi seemed to be best suited for voxel sizes equal or larger than 0.4 mm-isotropic and cylinders larger than 1 mm diameter and 2 mm length (Duarte Coello et al., 2024). One study diverges from the rest in terms of aims (Duarte Coello 2023), as its focus is the improvement in the accuracy of the quantification of the PVS morphometrics, namely length and width, and not in the detection or segmentation of PVS *per se*. PVS morphometrics are traditionally assessed by measuring the maximum and minimum diameters of the ellipsoid that encircles each PVS (Ballerini et al., 2020). The traditional approach, implemented in Matlab by the function regionprops3, is accurate if the PVS are linear and straight, but not if they are curved or elongated with irregular shapes. Three sources have linked data/code repositories: Bernal et al. (2022), Duarte Coello et al. (2024), and Valdés Hernández et al. (2024) to enable compatibility in follow-up improvements, reproducibility, and fair comparability of these results with those from further analyses.

3.5. Application studies

Summaries of all included application studies (N = 59) are presented in Table 9. Methodological and clinical differences between the studies ruled out meta-analysis of the estimates obtained.

#### 3.5.2. Factors explored in relation to PVS burden

Application studies explored a range of factors in relation with PVS. These are: PVS morphology and topology (6 studies, 10.17%), broad epidemiology (typically clinical risk factors and cognitive/functional outcomes, 19 studies, 32.20%) in the presence or not of a pathology or disabling event (e.g., in patients with stroke, traumatic brain injury, or Parkinson’s disease, 4 studies), pathophysiology and manifestation of a particular disease or disorder (e.g., Parkinson’s disease, Huntington’s disease, autism, obstructive sleep apnoea, COVID, diabetes mellitus, among others, 22 studies, 37.29%), vascular function or vascular disease (including concurrent SVD lesions and white matter hyperintensity burden, 13 studies, 22.03%), cognition (10 studies, 16.95%), sleep (5 studies, 8.47%%), retinal vasculature parameters (2 studies, 3.39%), space flight duration (2 studies, 3.39%), and one study exploring PVS in relation to preterm birth. Often, multiple factors or themes were explored (e.g., PVS associated with cognition and established disease or disorder). Two studies (3.39%) investigated, in the same sample, various aspects of PVS heritability in twins (Choi et al., 2020; Lee et al., 2021)

#### 3.5.2. Computational PVS quantification methods used

Many of the computational methods used in the application studies have been described in section 3.3, and have been applied directly or used as the basis of PVS quantification approaches in application studies. The most frequently applied method was thresholding the response of the Frangi filter, as described by Ballerini et al.’s (2016, 2018) (12 studies) or in the frameworks proposed by Sepehrband et al., (2019) (10 studies), Smith et al., 2020 (1 study), or Ranti et al., (2022) (1 study), which in total make up for 54.24% of the application studies. Ten studies use CNN configurations, mainly U-Net and U-ResNet (16.95%), to assess PVS burden, and four studies use this approach to post-process the output from the Frangi filter. Morphological detection as proposed by Schwartz et al. (2019), followed by thresholding is used in six studies, while a study uses the morphological detection approach described by Sato et al., (1998).

Computational PVS quantification methods applied but not described in section 3.3. include approaches based on Guerrero et al.’s (2017) method (a deep learning/CNN approach), Yushkevich et al’s (2006) method (a semi-automated, 6-connected voxel active contours approach), and seven in-house methods (including thresholding and filtering methods, and approaches based on Dubost et al. (2019) and Sepehrband et al.’s (2019) methods). One study refers using the 2D U-Net approach for multisite learning as developed by Remedios et al., (2020) for segmenting haemorrhages in CT scans.These were not among ‘method development’ papers included in this review as they did not independently meet inclusion criteria (e.g., the quantification approach may not have been designed explicitly for PVS detection (Guerrero et al., 2017; Remedios et al., 2020), or may have relied heavily on a manual component (Yushkevich et al., 2006)). A list of the computational PVS quantification approaches and the frequency of their use in the application studies identified in this review is presented in Table 10.

**Table 9.**
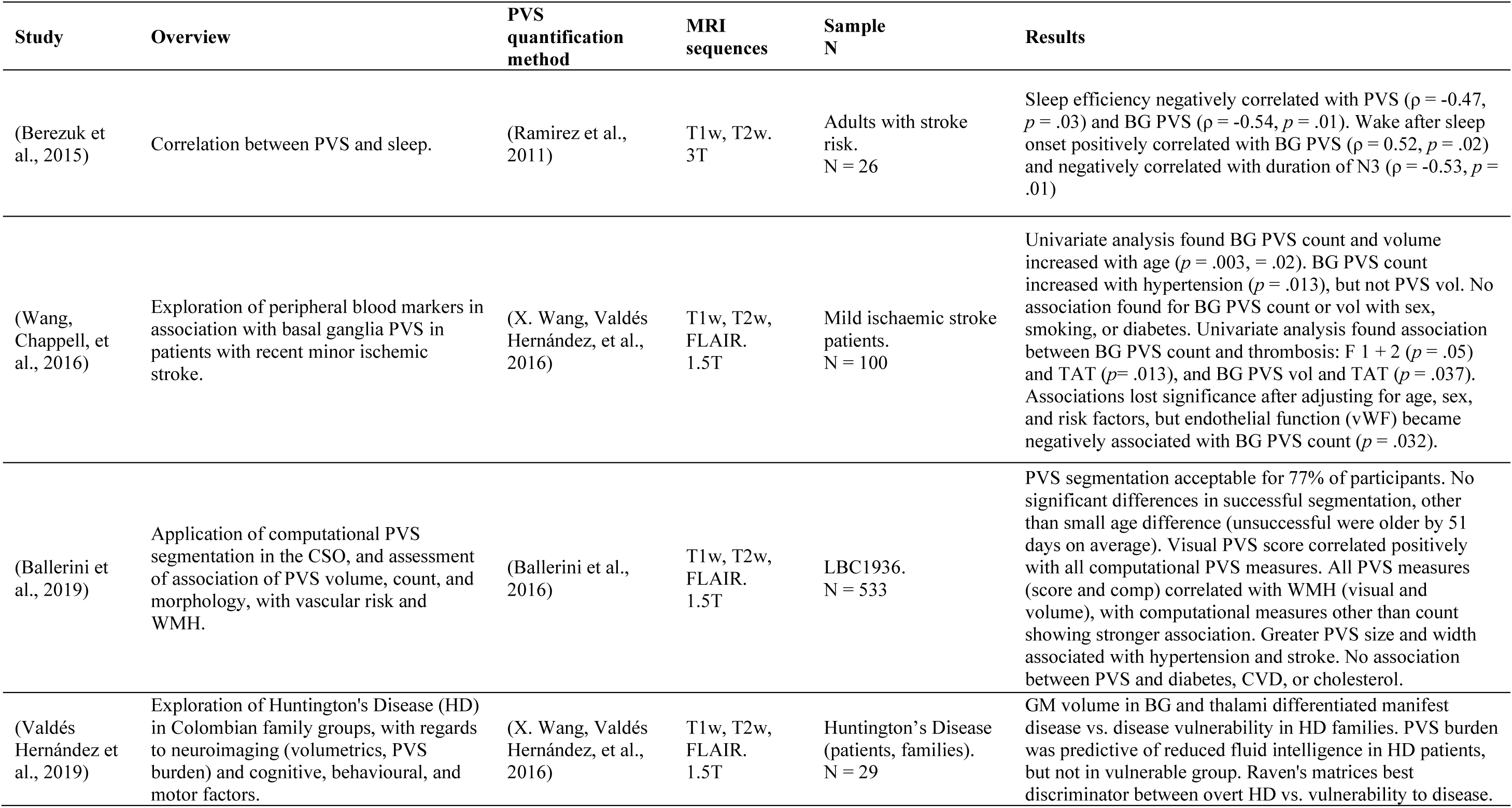
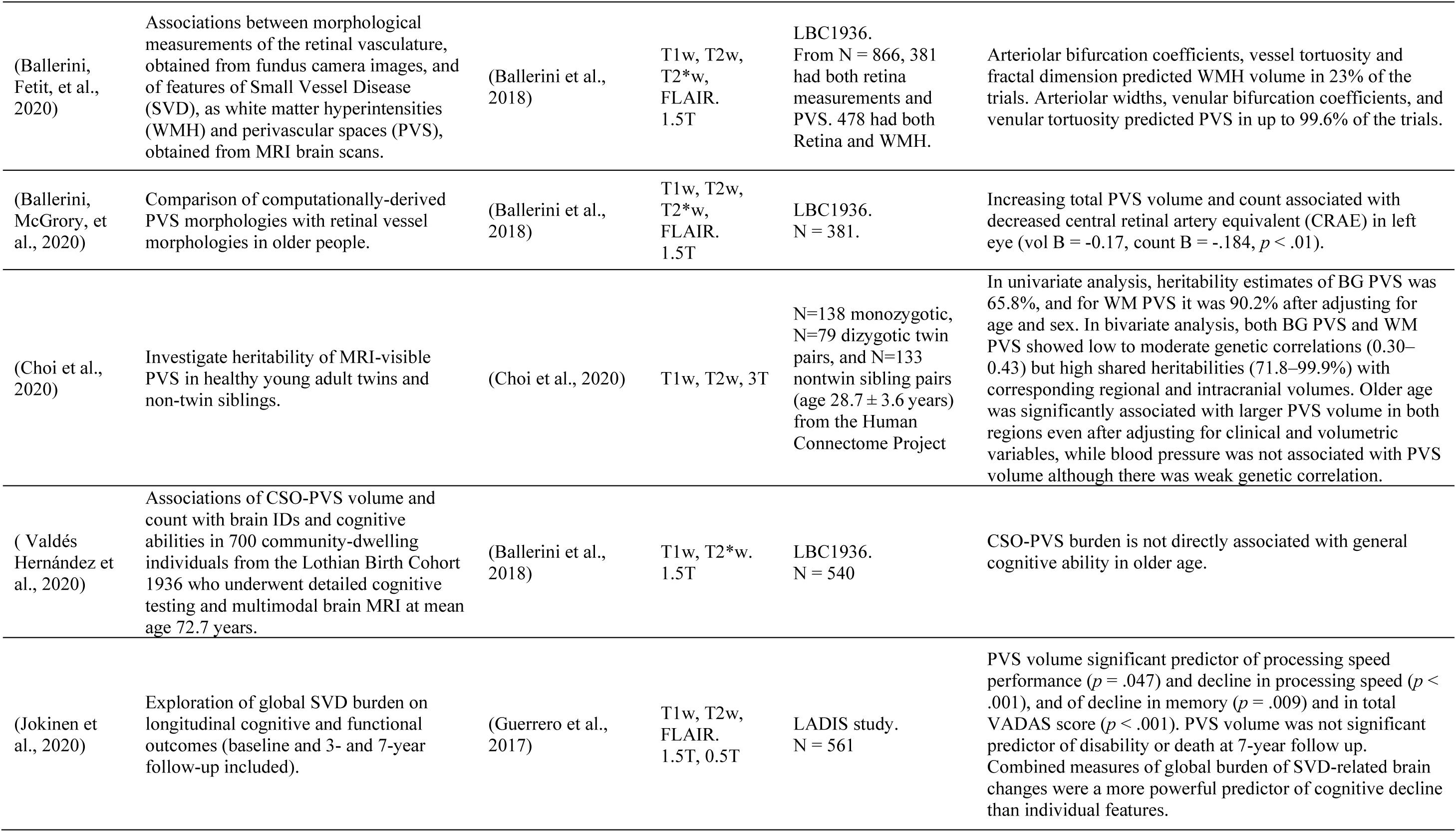
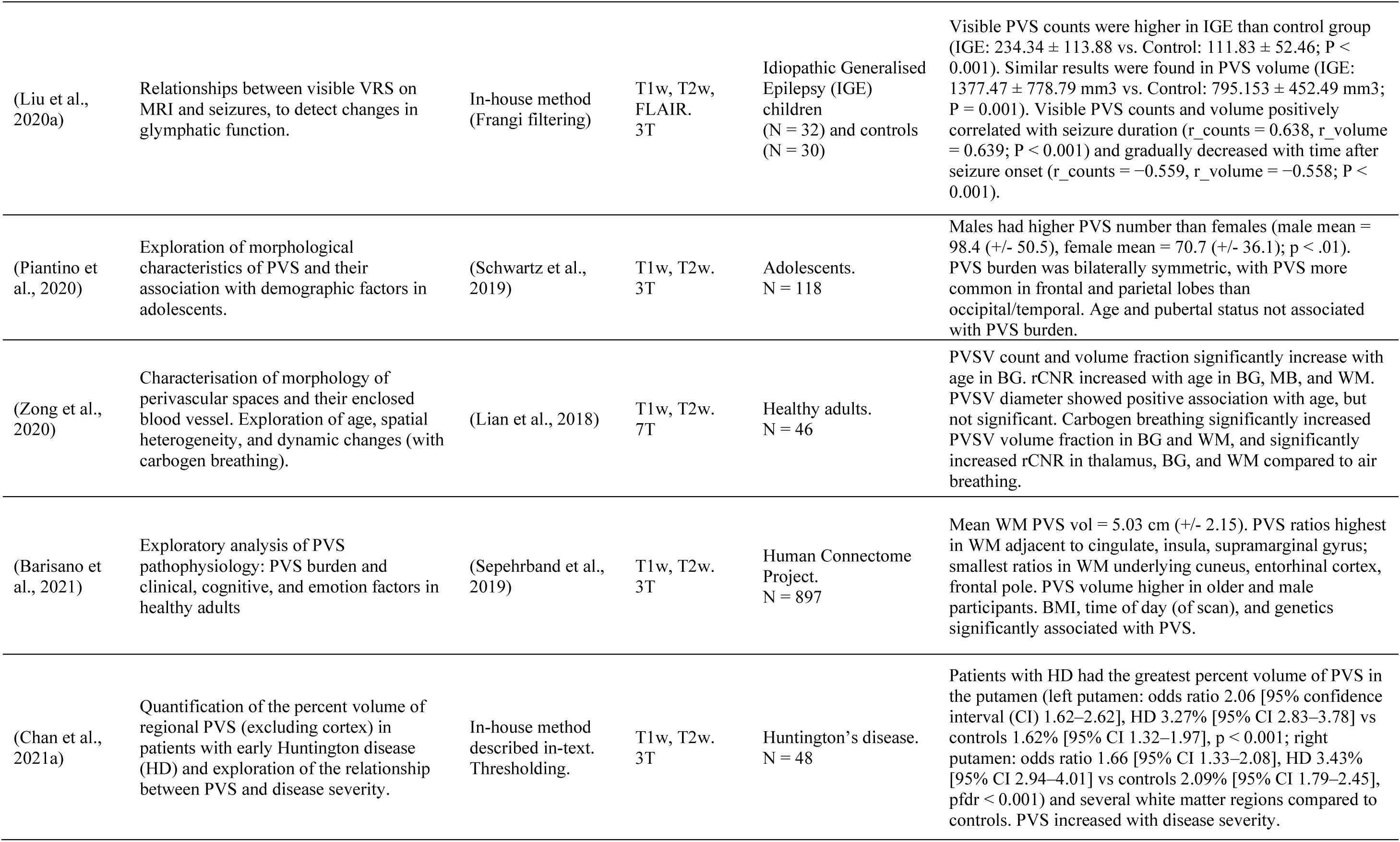
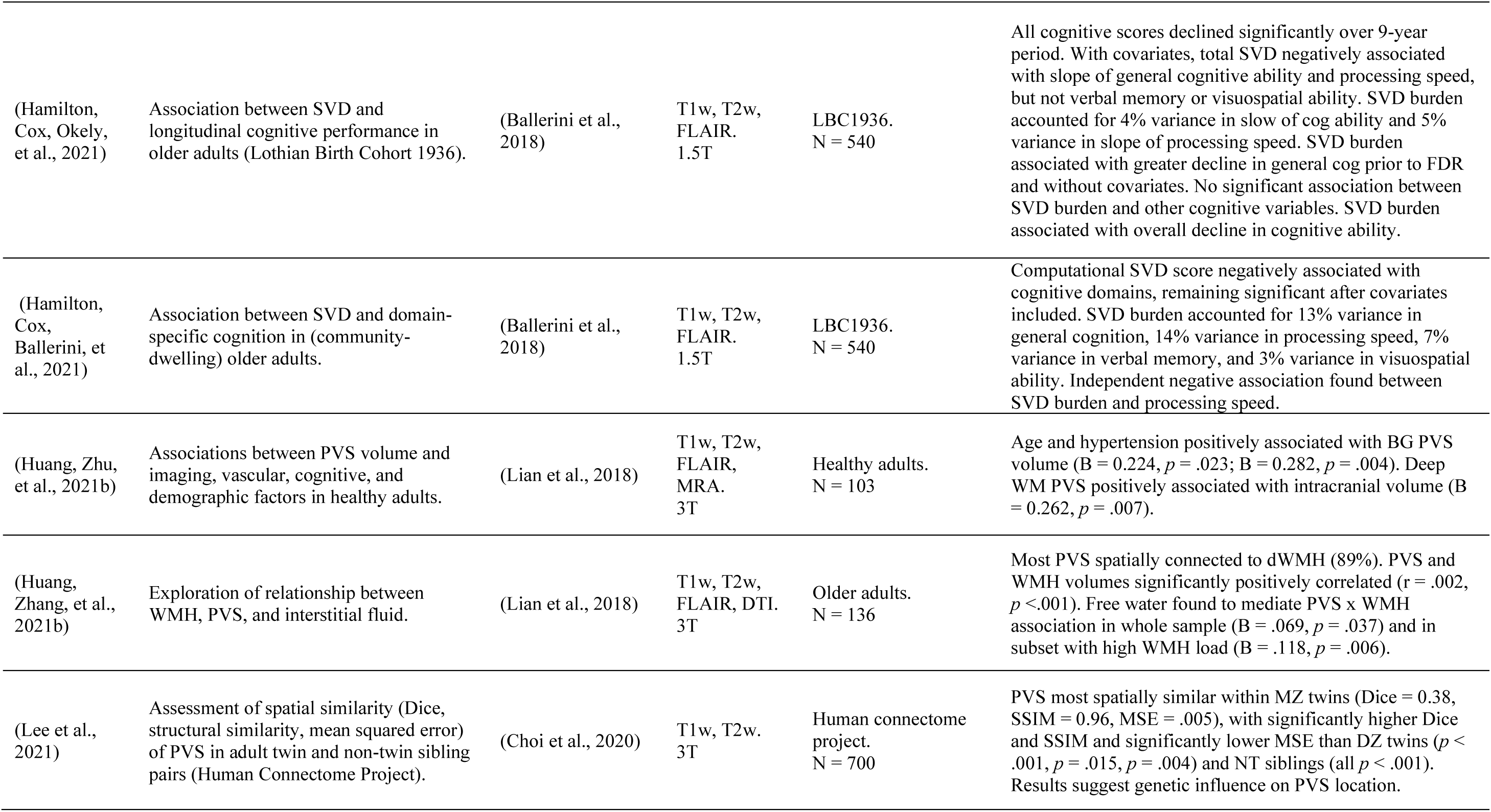
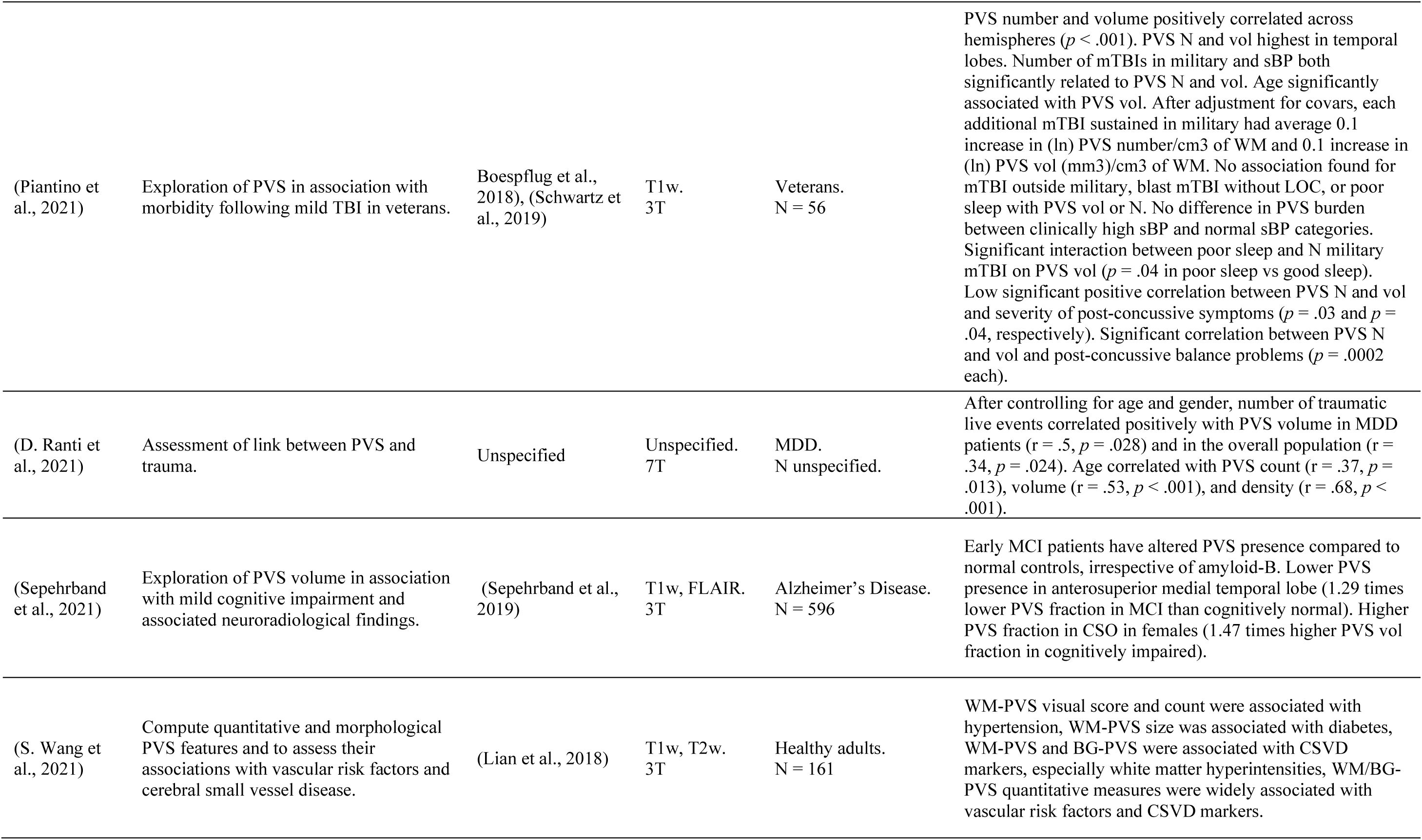
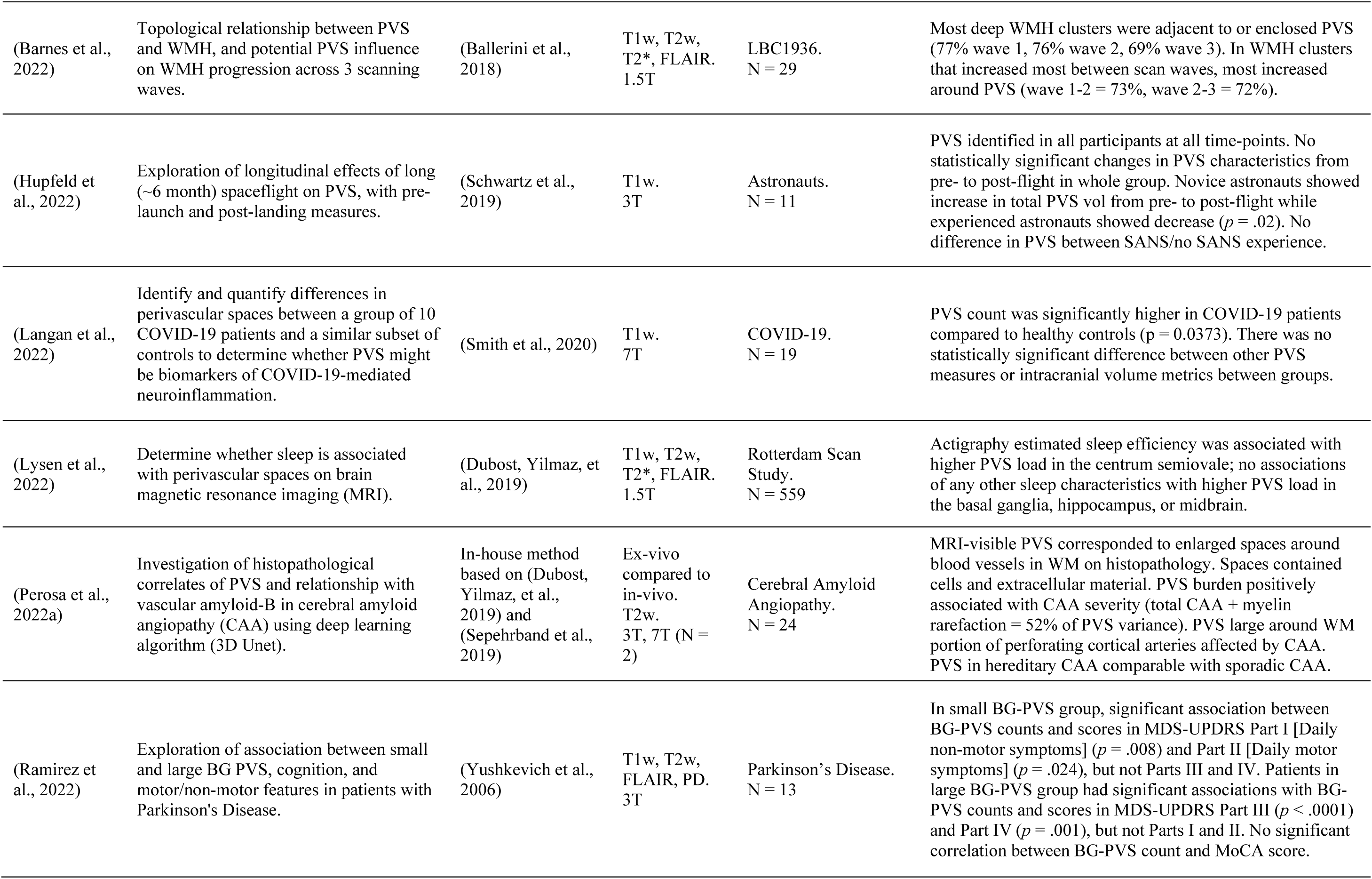
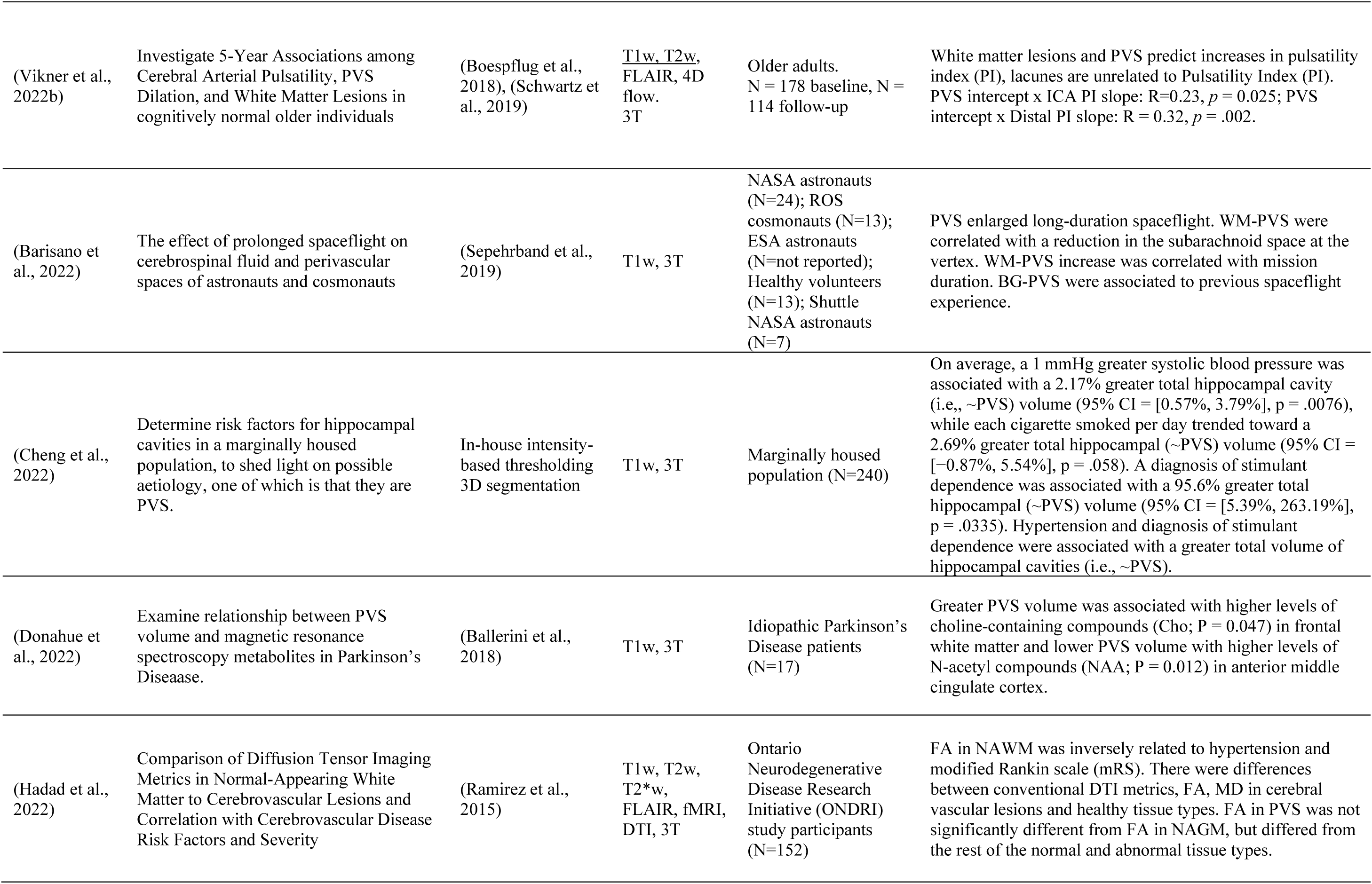
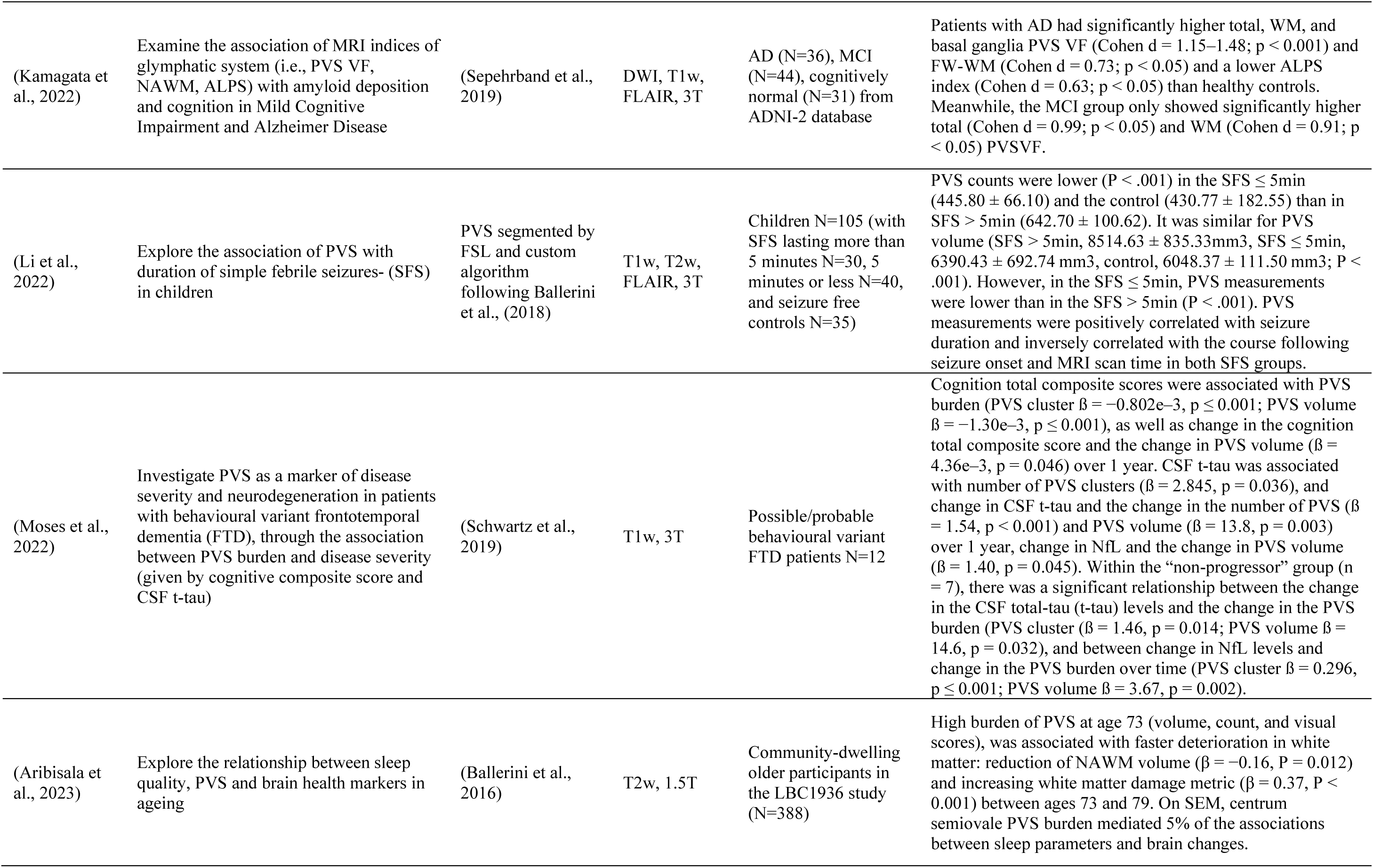
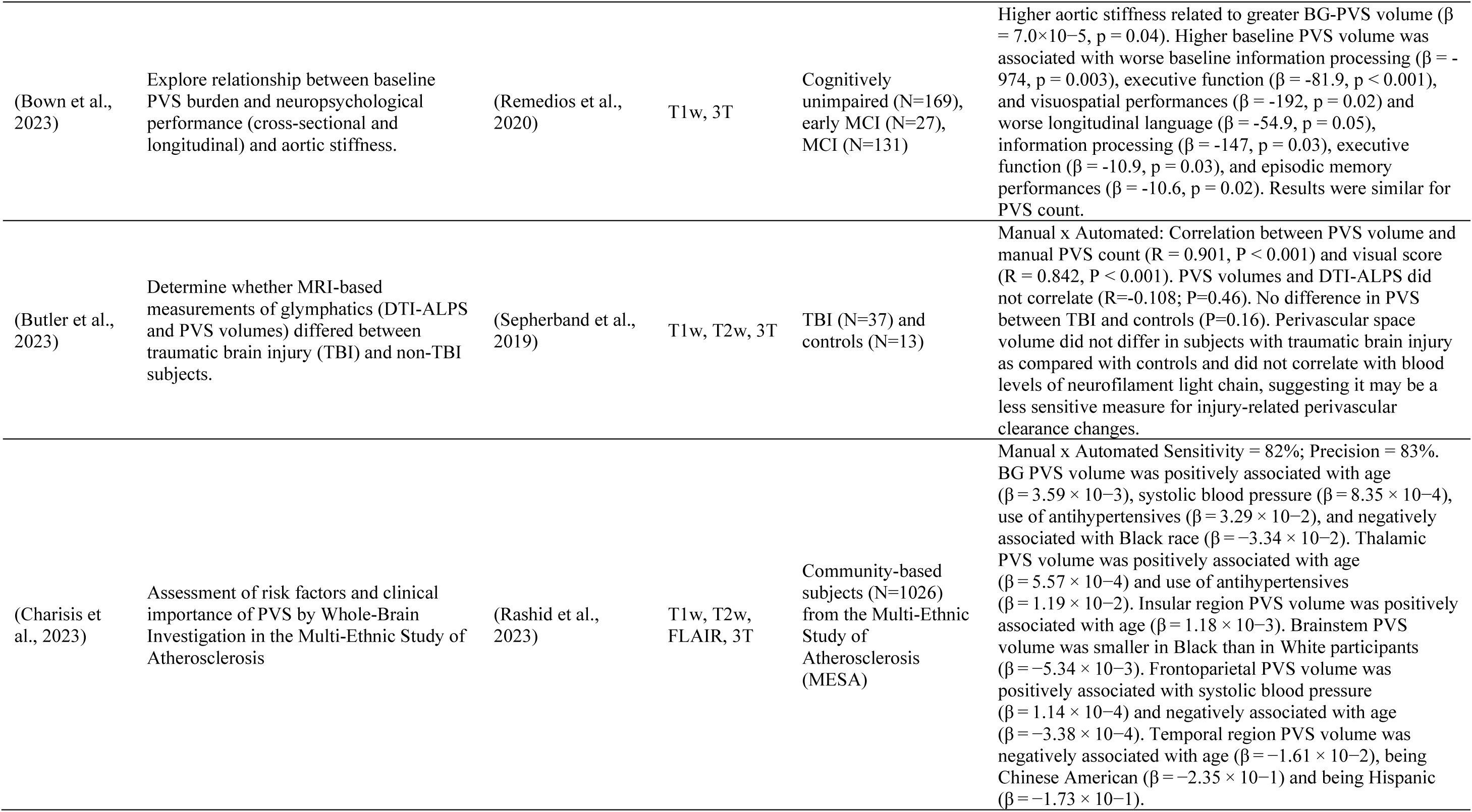
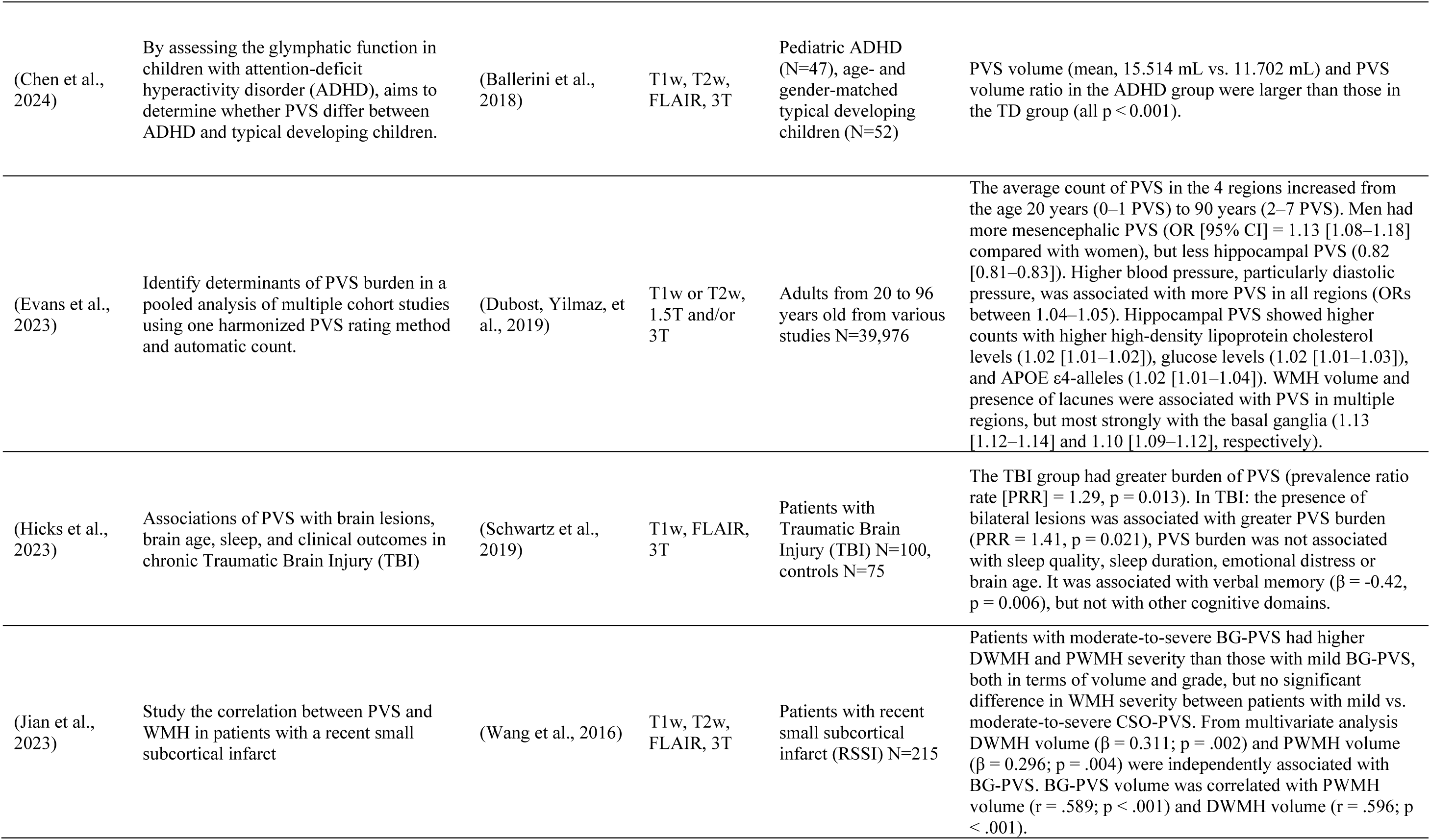
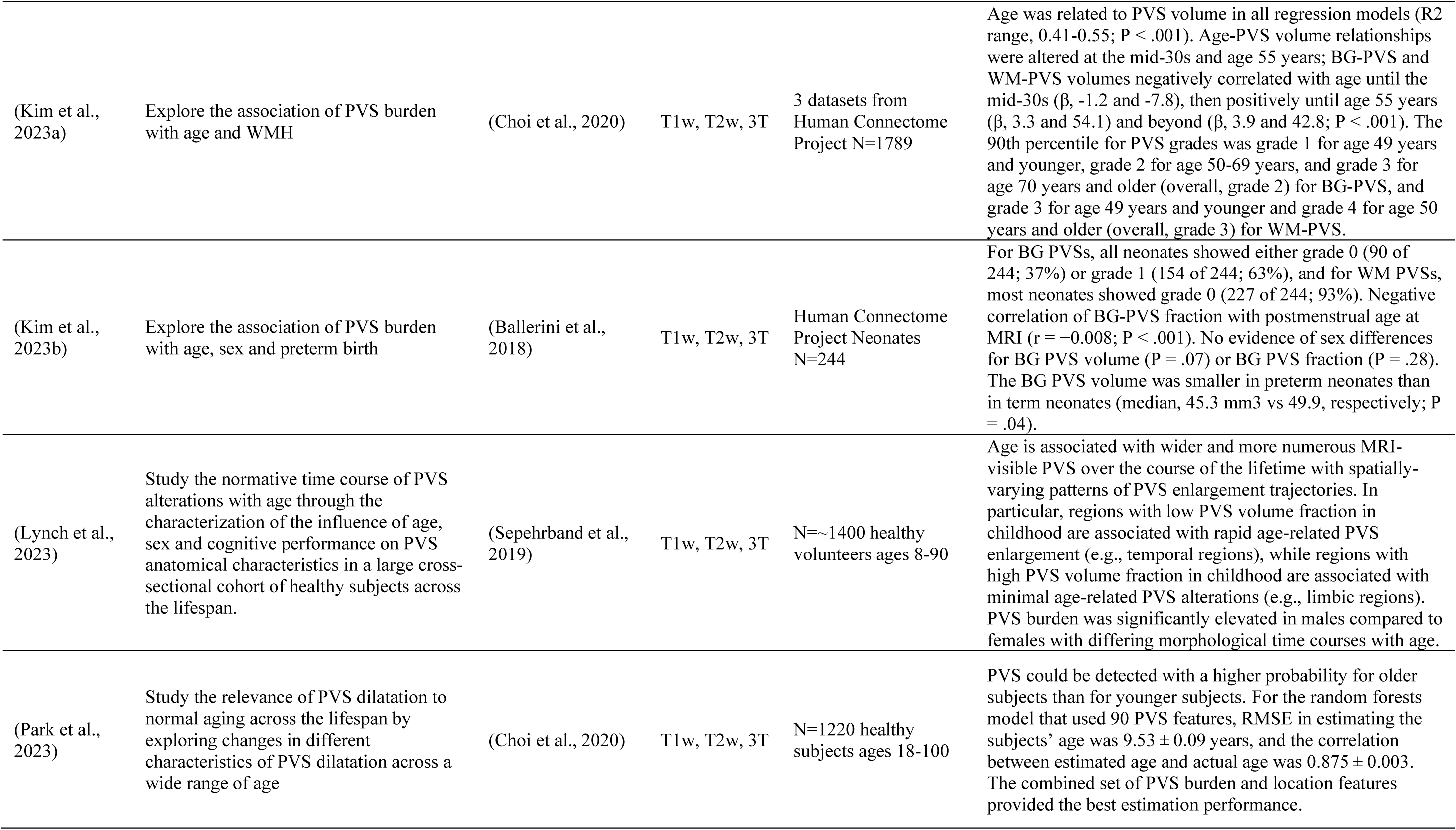
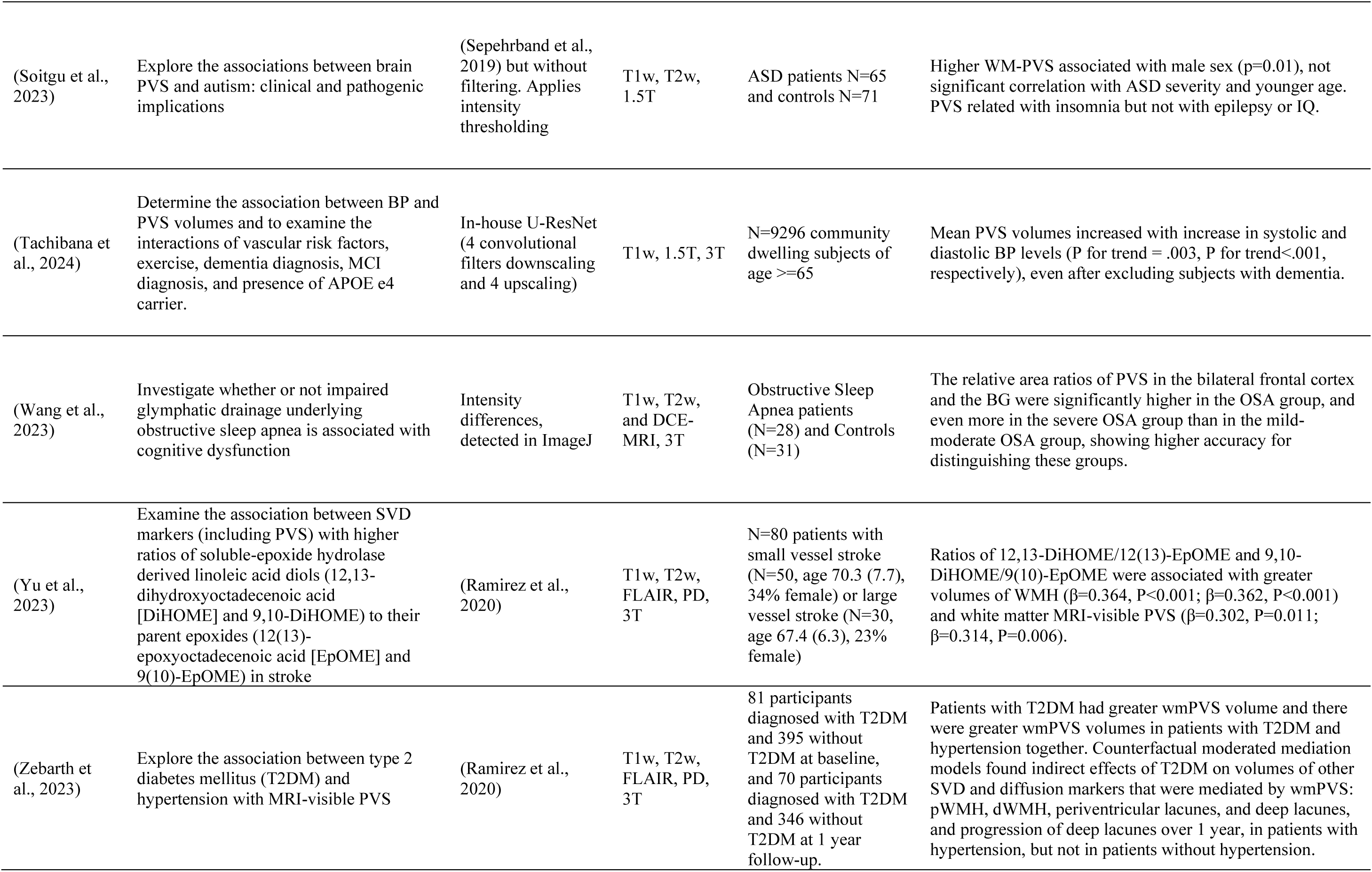
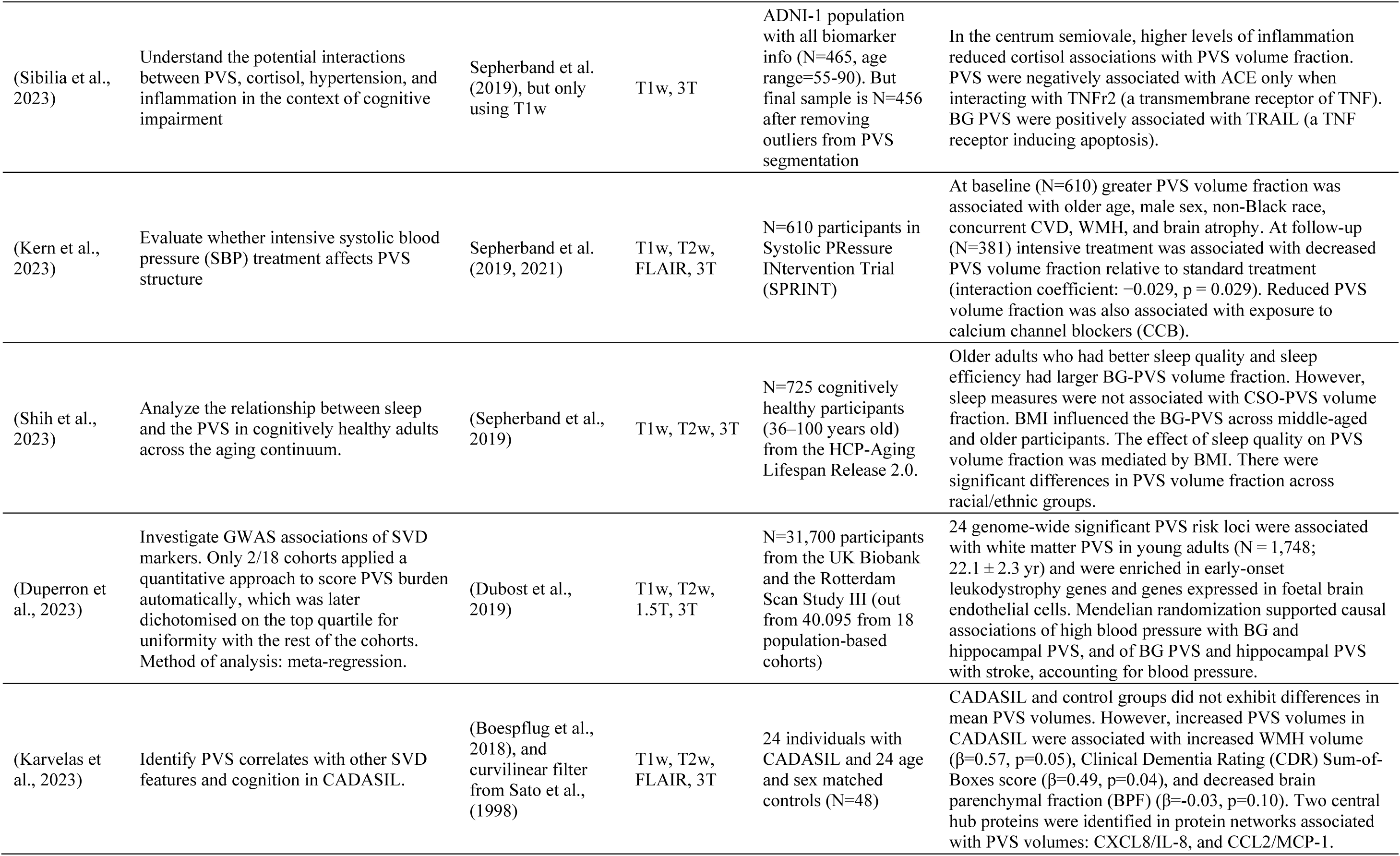
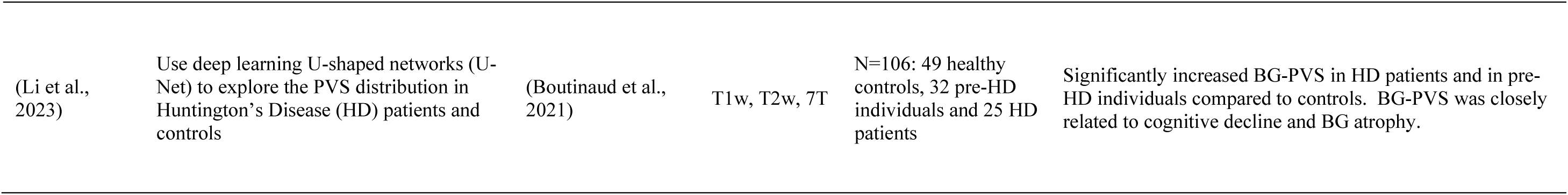
Application study summaries Legend: PVS: Perivascular Spaces, F1+2: Prothrombin Fragment 1 + 2, TAT: Thrombin-Antithrombin III complex, HD: Huntington’s Disease, vWF: von Willebrand’s Factor, GM: Grey Matter, WM: White Matter, CVD: Cerebrovascular Disease, CAA: Cerebral Amyloid Angiopathy, TBI: Traumatic Brain Injury, CSVD: Cerebral Small Vessel Disease, PVSVF: Perivascular spaces volume fraction, AD: Alzheimer’s Disease, MCI: Mild Cognitive Impairment, ADNI-2: Alzheimer’s Disease Neuroimaging Initiative second recruitment wave, ASD: Autism Spectrum Disorder, RMSE: Root Mean Square Error, T2DM: Type 2 Diabetes Mellitus, wmPVS (or WM PVS): Perivascular Spaces in the White Matter. Note: Choi et al., (2020) is a method development study as well as an application study. Therefore, it is included in both: Table 6 and Table 9, with the relevant information for each analysis.

**Table 10.**
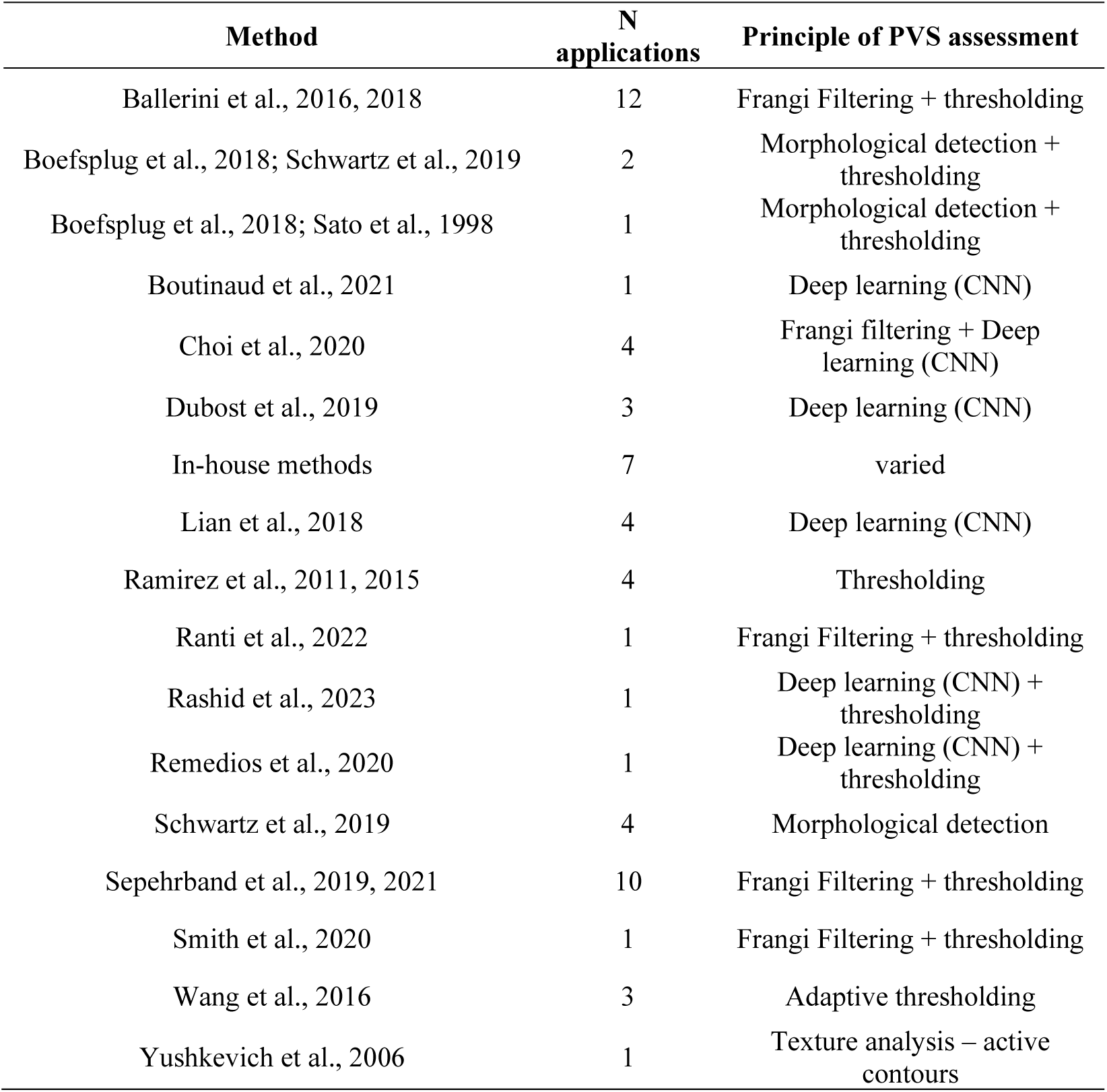
Frequency of computational PVS applications in included studies.

#### 3.5.3. MR sequences and field strengths

Frequencies of MR sequences and field strengths used in ’application’ studies in comparison with other study types are outlined in section 3.2.3, Tables 4 and 5. The most common combinations of MR sequences used in these studies were T1w and T2w (19 studies). This was also the combination of sequences used by Choi et al., (2020), which can be considered both: a ‘method development’ and an ‘application’ study. T1w alone, and the combination of T1w, T2w, T2*w and FLAIR (10 studies each) were also commonly used. The latter, however, was not used specifically for identifying PVS, but was used by full pipelines to extract regions of interest and obtain other imaging markers that were analysed by the studies in relation to PVS.

Application studies most frequently used 3T imaging (42 studies, 71.19%), although a large proportion used 1.5T imaging (20 studies, 33.9%) and only five studies used 7T (8.47%). This differs from method development studies, where distribution of field strengths was fairly evenly distributed, with a slight bias towards 1.5T imaging.

#### 3.5.4. Population types and sample sizes

Application studies were almost evenly split between those using a clinical population (50.85%) and those using only a non-clinical population (49.15%). Participants were most commonly independently living older adults (>60 years; 18 studies, 30.51%), followed by patients with cerebrovascular disease-related phenotype (e.g., stroke patients, hypertensive patients, patients with artherosclerosis) (9 studies, 15.25%). In terms of population characteristics, application and method development studies shared similar distribution (i.e., proportion). Median sample size of PVS application studies was N = 161 (IQR = 482.5), with two multicentre epidemiological studies with sample sizes greater than 30,000 (Evans et al., 2023; Duperron et al., 2023) and five with more than 1,000. Broad population type and age-groupings used in application studies compared to method development and improvement studies are presented in Tables 3 and 4, respectively.

#### 3.5.5. Application results

Six areas of interest emerged in common across a number of application studies: age, sex, hypertension, diabetes, sleep, white matter hyperintensities (WMH), and cognition.

##### 3.5.5.i. Age

Eleven studies reported a positive association between computational PVS measures and age (Barisano et al., 2021; Huang, Zhu, et al., 2021b; Piantino et al., 2021; D. Ranti et al., 2021; Wang, Chappell, et al., 2016; Zong et al., 2020; Choi et al., 2020; Aribisala et al., 2023; Evans et al., 2023; Park et al., 2023; Kern et al., 2023), one of which was a study in young adults (mean age = 28.8 years). But studies on life-time trajectories point at varying patterns of these associations throughout the lifespan (Kim et al., 2023a; Lynch et al., 2023). One study of adolescents (N=118) found no association between age and PVS burden (Piantino et al., 2020), and neither another study on children (Soitgu et al., 2023). A study on chronic traumatic brain injury did not find PVS burden being associated with brain age (Hicks et al., 2023). Studies that analysed the PVS association with age in different regions had conflicting results, with the majority reporting a positive association/correlation with age in all regions, and one study finding a negative association between age and PVS volume in the temporal region (Charisis et al., 2023; N=1026).

##### 3.5.5.ii. Sex

Nine studies provided information on PVS burden aggregated by sex. Two found no association between PVS burden and sex (Wang, Chappell, et al., 2016; Kim et al., 2023b), while the majority(i.e., six out of nine) found PVS burden to be greater in males than in females (Barisano et al., 2021; Piantino et al., 2020; Evans et al., 2023; Lynch et al., 2023; Soitgu et al., 2023; Kern et al., 2023). It is worth noting, however, that Evans et al., (2023) performed a regional analysis and also reported no sex differences in the number of PVS in the hippocampus. Only one study found higher CSO PVS fraction (burden) in females compared with males (Sepehrband et al., 2021).

##### 3.5.5.iii. Hypertension

Eleven out of thirteen studies identified a positive association between hypertension and PVS burden (Ballerini et al., 2019; Huang, Zhu, et al., 2021b; Wang et al., 2021; Wang, Chappell, et al., 2016; Cheng et al., 2022; Hadad et al., 2022; Charisis et al., 2023; Evans et al., 2023; Tachibana et al., 2024; Zebarth et al., 2023; Duperron et al., 2023). Duperron et al., (2023), the largest study amongst the ones included, found causal associations of high blood pressure with BG and hippocampal PVS, and of BG PVS and hippocampal PVS with stroke, accounting for blood pressure, even after Mendelian randomisation. In Wang et al. (2021), basal ganglia PVS count was associated with hypertension, but not PVS volume. Piantino et al. (2021) found no association between systolic or diastolic blood pressure and PVS burden, and Choi et al., (2020) did not find blood pressure to be associated with PVS volumes.

##### 3.5.5.iv. Diabetes

No association between PVS burden and diabetes was found in two studies (Ballerini et al., 2019; Wang, Chappell, et al., 2016), but a positive association was identified in a third study between white matter PVS burden and diabetes (S. Wang et al., 2021), and in another study exploring specifically the putative role of PVS burden in type 2 diabetes mellitus (T2DM) (Zebarth et al., 2023). This study also found that indirect effects of T2DM on volumes of other SVD markers (i.e., WMH and lacunes) were mediated by the PVS burden in the white matter.

##### 3.5.5.v. Sleep

The relationship between sleep and PVS burden was explored in eight studies (Berezuk et al., 2015; Lysen et al., 2022; Piantino et al., 2021; Aribisala et al., 2023; Hicks et al., 2023; Soitgu et al., 2023; Wang et al., 2023; Shih et al., 2023), with all except one (Hicks et al., 2023) identifying an association between at least one metric of sleep and PVS burden. Berezuk et al., (2015) and Lysen et al., (2022) reported a negative correlation between both sleep efficiency and PVS burden. Berezuk et al., (2015) further reported a negative association between the duration of N3 (i.e., deep sleep) and PVS burden, and a positive association between wakefulness after sleep onset (i.e., number of interruptions to sleep) and BG-PVS burden. But Lysen et al. (2022) found no associations between any other metric of sleep and PVS burden in any location. In a very heterogeneous sample Shih et al., (2023) found that older adults who had better sleep quality and sleep efficiency had larger BG-PVS volume fraction, while CSO-PVS volume fraction was not associated with sleep measures. But this study also found that the effect of sleep quality on PVS volume fraction was mediated by BMI, and that there were significant differences in PVS volume fraction across racial/ethnic groups. Piantino et al., (2021) reported a significant association between poor sleep and PVS volume, with PVS volume greater in those with poor sleep and mild traumatic brain injury (TBI) than those with good sleep and mild TBI. Aribisala et al., (2023) found that CSO-PVS volume mediated 5% of the associations found between sleep parameters and brain structural changes. Soitgu et al., (2023) reported that white matter PVS were related with insomnia in autistic children, and Wang et al., (2023) found that PVS volume ratios were associated with obstructive sleep apnoea (OSA), showing higher accuracy for distinguishing groups of patients with different degrees (i.e., severity) of OSA.

##### 3.5.5.vi. White matter hyperintensities

Nine studies reported positive associations between PVS burden and WMH volumes (Ballerini et al., 2019; Huang et al., 2021; Wang et al., 2021; Barnes et al., 2022; Aribisala et al., 2023; Evans et al., 2023; Kern et al., 2023; Karvelas et al., 2023; Jian et al., 2023), with the latter finding the association only for PVS in the BG and not in the CSO. Two of them found also topological relationships between the occurrence of PVS and the location of WMH deep clusters (Huang et al., 2021; Barnes et al., 2022). A study in chronic TBI (Hicks et al., 2023) reported a positive association between PVS burden, given as prevalent ratio rate, and bilateral lesions. However, it is unclear if the lesions referred are only as a consequence of the injuries or including also WMH.

##### 3.5.5.vii. Cognition

Fifteen studies explored the relationship between cognition and PVS with results varying depending on the characteristics of the cohorts and the cognitive domains assessed. Hicks et al., (2023), for example, reported PVS burden impacting verbal memory but not any other cognitive domain. Two studies found no direct or significant association between general cognition (general cognitive ability, Montreal Cognitive Assessment score) and PVS burden (Ramirez et al., 2022; Valdés Hernández et al., 2020). Valdés Hernandez et al. (2019) reported PVS burden as predictive of reduced fluid intelligence in Huntington’s disease patients but not in family members without overt disease, while Soitgu et al., (2023) did not find association between PVS burden and IQ in autistic children In mild cognitive impaired (MCI) and dementia, all studies reported different measures of PVS being more relevant in MCI (Seperhband et al., 2021; Bown et al., 2023) and frontotemporal dementia (Moses et al., 2022) groups than in controls. Jokinen et al (2020) reported PVS volume to be a significant predictor of processing speed, decline in processing speed, decline in memory, and of total vascular dementia assessment (VADAS-cog) score. PVS burden was related to poor cognition in CADASIL (Karvelas et al., 2023), and in Huntington’s disease (Valdés Hernández et al., 2019; Li et al., 2023). Kamagata et al., (2022) reported higher PVS volume fraction (total, in basal ganglia, and white matter) in Alzheimer’s disease (AD) patients compared to controls, and higher white matter PVS volume fraction in the mild cognitive impaired (MCI) group compared to controls. Hamilton et al. (2021a, 2021b) reported that worse total SVD burden (PVS-inclusive) was associated with cognitive decline, but did not report PVS-specific results.

### 3.6. Risk of Bias assessments

Risk of bias assessments were conducted based on the QUADAS-2 framework, with potential sources of bias assessed including: patient selection, index method bias (i.e., does the method used to quantify PVS introduce bias), reference accuracy (i.e., accuracy of ground truth), reference blind to index (i.e., was PVS quantification influenced by prior knowledge of ground truth), same reference for all, inclusion of all participants, method/application matches review question, applicability, and overall assessment.

#### 3.6.1. Method and improvement

Due to the small number of ‘improvement’ studies, these were grouped with ‘method development’ studies in the presentation of risk of bias assessments. In general, 42.31% of method and improvement studies were rated as low risk of bias, 17.31% were rated as instilling some bias concerns, and 40.38% had high risk of bias. One study did not provide enough detail for a bias assessment. The individual domain with the most frequent high risk of bias scoring was ‘applicability’ (how well the index measure can be applied to PVS quantification in the context of the review question). Reasons provided when rating applicability as having a high risk of bias included a lack of detail in the methodology, lack of validation testing, and no ability to discriminate between PVS and other lesion types. A summary risk of bias plot for method and improvement studies is shown in Figure 6.

**Figure 6.**
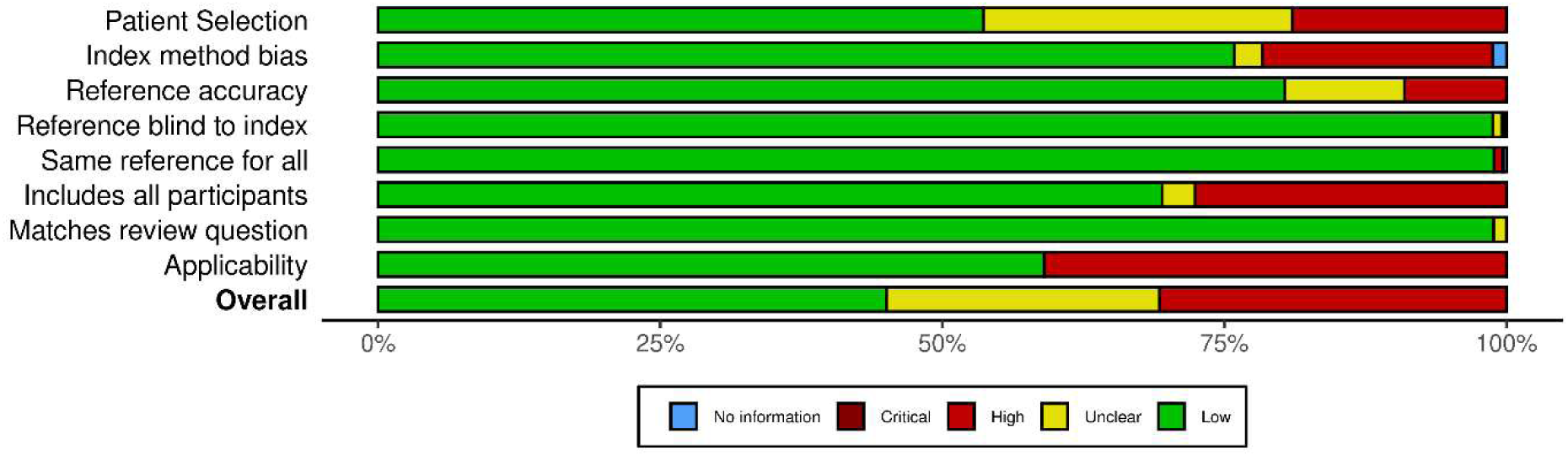
Risk-of-bias summary plot for method development and improvement studies.

#### 3.6.2. Application

There was low risk of bias in 59.32% of application studies, some risk in 25.42%, and high risk in 15,25%. The domain most frequently scoring as having a high risk of bias was ‘includes all participants’, rated high risk in 49.15% of application studies. Often, a small number of patients were excluded from application studies due to poor image quality (noise and motion artefacts) which compromised PVS quantification performance, without a statistical analysis of the relevance of missing values as a consequence. A summary risk of bias plot for application studies is provided in Figure 7.

**Figure 7.**
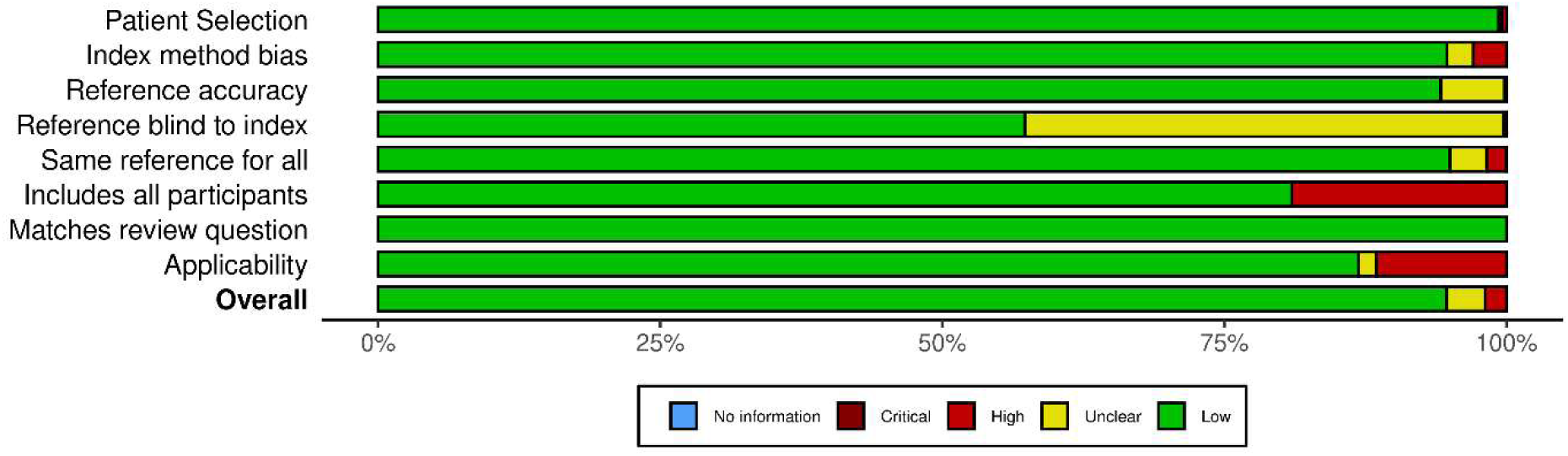
Risk-of-bias summary plot for application studies.

## 4. Discussion

### 4.1. Overview

We conducted a systematic review of the development, improvement, and application of computational PVS quantification approaches, including studies published up to September 2023. We explored 67 approaches involving 48 methods presented as part of 45 publications, eight frameworks or suggestions for improving some of these methods, nine in-house pipelines or methodologies using different elements or combinations from some of the computational approaches presented, and two re- purposed implementations from approaches developed for other purposes, these applied to clinical research. We collated information on the main aims and outcomes of each study, and extracted and analysed data relating to MR imaging sequences and field strengths used, population types and sample sizes, method characteristics including training and validation, and results.

The number of studies developing or using computational PVS quantification approaches has increased steadily since the first identified study of this type in 2002 (Kruggel et al., 2002), reaching a peak of 8 computational PVS assessment methods published in 2019, year from which the number of studies applying these methods has been experiencing an exponential growth. Currently, research into PVS using quantification approaches comprises a large field, favoured by the increase in the ability to analyse large datasets for which visual scores are lacking, and the increased interest in the function and relevance of PVS in brain health.

The use of a morphological filter to enhance PVS-like structures, with Frangi being the most commonly used, and the use of a U-Net configuration with or without residual connexions, were the two most widely applied and promising approaches, which have been independently validated. The commonest PVS feature (s) that are measured are volume and count, obtained from the PVS segmentations. From them, PVS volume, absolute, or as a percentage in the region of interest, appears to be the most consistent PVS parameter used by the clinical studies that applied a computational approach to assess PVS.

Imaging data from older adults, or a wide age range of adults, has most frequently been used in both the development and application of computational PVS quantification approaches, consistent with studies using non-computational approaches to PVS quantification (Francis et al., 2019). MR imaging was most often obtained from 1.5T and 3T imaging, with a recent increase in the use of 7T imaging. T2w imaging was most commonly used across all study types, often in combination with T1w and FLAIR imaging, these mainly being used as part of the frameworks developed for eliminating confounds and generating regions of interest, although a number of application studies used T1w only for assessing the PVS. We limited the scope of the present review to static, structural imaging of PVS, as opposed to including fluid dynamics, functional imaging, and/or cerebrovascular reactivity studies which may make greater use of alternative imaging modalities, including diffusion tensor imaging, 4D-flow, positron emission tomography (PET), single-photon emission computed tomography (SPECT).

### 4.2. Method development studies

The most common approaches to computational PVS quantification were thresholding, thresholding and filtering, and deep learning (CNN) methods. Deep learning approaches have become almost ubiquitous since 2017, with 19 of the 48 method development studies using CNN methodology. They have higher processing speed and capacity than the more conventional methods, and a single architecture can be designed and trained to separately assess different confounding features, e.g., lacunes, PVS, and WMH. But the large number of parameters that CNN architectures manage and need to adjust require a breath of size and variability in the training dataset impossible to achieve at present. Then, as the resultant model is overfitted to the imaging parameters and clinical presentations of the dataset used for training, these models would need to be re-trained for every different cohort at hand. This entitles a reasonable number of ground truth manual segmentations, not always feasible and highly observer-dependant. While some of the architectures reviewed here have been made publicly available, we could not find any pre-trained CNN model to independently test the accuracy levels reported in their original publications. The conventional approaches that threshold the output from a morphological filter that enhances PVS-like structures, on the other hand, do not require large amount of data to be tuned, but still the threshold and some parameters would need to be adjusted depending on the imaging protocol and image quality. Also, separate developments would be required to remove confounding pathology and deal with artefacts. In terms of reproducibility, sustainability, and applicability, both types of approaches, being fully automatic, could reach high levels, but these would depend, for now, on the skills of the computer programmers that generate and manipulate them, and data availability.

Computational segmentation pipelines used different pre-processing steps. General steps involved co-registration of the different MRI sequences to the one in which PVS are assessed, and intensity normalisation. No justification of the intensity normalisation approach has been given or evaluated in any of the studies. A study classed as improvement (Valdés Hernández et al., 2024) only evaluates the impact of the point, within the pipeline, in which a linear intensity normalisation is done. Evaluation of the impact of the type of intensity normalisation in the results is currently lacking. Few methods perform bias field correction. But the similarity indices and performance metric of all the methods presented is similar, raising questions about the impact bias field correction may have in the overall performance of the method. Few methods involve denoising too, which, contrary to the correction of inhomogeneities in the b1 magnetic field aimed at filtering out the low-frequencies, is designed to remove the high-frequencies in the image. But given the nature of the task, the application of a band- pass filter in the preprocessing stage may not be the best option to consider. Another preprocessing step worth mentioning is the generation of regions of interest (ROIs), mostly carried out using freesurfer. In some other cases, ROIs are the grey and white matter tissue classes. Given the importance of this step, its validation in different scenarios would have been required. For example, in the presence of narrow ventricles with disconnected horns resembling tubular structures, presence of calcified vessels resembling tubular structures that are also hypointense in T1w images, to mention just two of the most common scenarios (Valdés Hernández et al., 2024).

Computational PVS quantification approaches were validated against a range of reference standards; most often visual rating scores (27.08% of studies), although there was a relatively high proportion (22.92%) of them which used manual segmentations as a reference standard. While visual rating scores may be less time consuming and applicable to much larger sample sizes than producing manual segmentation data, high correlation between segmentation results and visual scores does not necessarily mean that the segmentations are good. Gonzalez Castro et al., (2017) showed that neuroradiological assessments can be subconsciously influenced by the degree of the overall disease present. Therefore, it is likely that, for example, visual rating scores of PVS in the basal ganglia in patients with SVD would be highly correlated with PVS segmentations, but perhaps not so in the basal ganglia of obstructive sleep apnoea patients that would not necessarily have any other confounding imaging feature in their brain scans. Further exploration of the correlation between different computational parameters with visual scores in cohorts with different characteristics is needed. Manual segmentations, on the other hand, enable comparison of more PVS metrics (e.g., volume, width, length) than visual-only assessments, but they have been only available for a limited number of scans or for one slice per region per scan, and most of the studies lack reports of inter-annotator reliability assessment of the manual annotations used.

Not all of the method development studies provided information on the performance or validation of their method, with three studies not using any reference standard of ground truth. Performance metrics used in those that did report results varied, although more common measures included correlations between computational and non-computationally derived PVS values, number of false positives/false negatives, and spatial similarity (Dice coefficient). It is important to note that method development studies reporting the strongest validation results between computational and manual/visual PVS quantification often had either small sample sizes or a significant user-intervention aspect to their methodology in their validation datasets. For example, Weurfel et al. (2008) reported an intraclass correlation coefficient of 1 (a perfect score), but data from only 4 participants were used, and Ranti et al. (2022) report a DSC score of 0.99 (almost perfect spatial overlap) but their method includes a final ‘manual correction’ stage.

A large proportion of method development studies lacked sufficient information with regards to parameters to be adjusted in methods using filtering and thresholding approaches, differentiation between PVS and other lesions (notably lacunes, which can have similar appearance), or applicability (and adjustments required) when using multi-site or multi-modality imaging. Some studies provided little to no information about MR equipment and sequences used including critical factors like voxel size and slice thickness, nor about the population from which imaging data were derived or the process of participant selection. This is reflected in the large number of studies rated as having a high risk of bias for applicability. Deep learning is able to deal with changes in voxel size and slice thickness as long as the model has been trained with such a data. The vanilla implementation of the Frangi filter, for example, is also not able to deal with anisotropic data while the implementation of the Jerman filter in Matlab is. This information, therefore, impacts in the subsequent number of applications using the methods developed.

Given the varied approaches to assessing and reporting performance of novel PVS quantification methods, it may be beneficial to work towards a standard for validation and reporting. For example, a dedicated open-source PVS quantification validation dataset/repository would be worthwhile, with representative PVS and SVD, image sequence, image quality, and multi-site imaging, and phantom data. This would enable validation of novel methods by developers themselves and reproducibility assessments from outside parties. It is worth noting that although most method developments have used data acquired at the commonly used field strengths and from relevant populations – e.g., older individuals who are therefore likely to have bigger or more visible PVS, some atrophy, and confounding pathologies like WMH –, this does not imply that they would yield successful results if applied without variations, to MRI data from more, less, or simply different disease and age populations, even if acquired in the same scanners. It may also be of benefit to identify minimum reporting standards for validation performance, such as agreement on a preferred reference standard, minimum required validation sample size, and relationship, accuracy, and spatial overlap metrics.

### 4.3. Improvement studies

We identified eight studies concerned with improving computational PVS quantification. Targets for improvement included maintaining PVS detectability with image compression (Paz et al., 2009), reducing noise and motion artefacts (Bernal et al., 2020, 2021, 2022; Zong et al., 2021; Valdés Hernández et al., 2024), comparing filtering approaches when applied to synthetic data (Bernal et al., 2022; Duarte Coello et al., 2024), works on physical and *in-silico* phantoms for establishing the limits of validity of PVS assessment methods (Duarte Coello et al., 2024), comparing PVS segmentations from T1w vs. T2w (Valdés Hernández et al., 2024), drawing recommendations for threshold selection (Valdés Hernández et al., 2024), tuning of filter parameters (Duarte Coello et al., 2024; Valdés Hernández et al., 2024), and improving the calculation of PVS morphometrics (i.e., width and length) (Duarte Coello et al., 2023). While compression, noise, and motion artefacts appeared to have viable proposed solutions, one study identified remaining issues with differentiation between PVS and other hyperintense (in T2w) features, finding none of the more commonly used filter methods (Frangi, Jerman, RORPO) were effective at differentiating lesion types. This review has identified two ways in which methods have approached this issue: 1) by excluding WMH from the PVS ROIs, and 2) by combining multiple sequences simultaneously (e.g., Boespflug et al. (2018), identify as PVS the voxels with CSF-like intensities in T1w, T2w, FLAIR and PD, while Seperhband et al., (2019) segment on the combination of T1w and T2w and look for the intensity profile in FLAIR excluding the voxels on the top 10 percentile of the FLAIR signal distribution). But both approaches has drawbacks. The first one dismisses the existence of PVS inside WMH, consequently leading to misleading results, for example reporting few PVS in patients with wide and confluent WMH, just because the “clean” ROIs were small. The second one could also lead to erroneous results depending on the sensitivities of the MRI sequences involved, especially in the presence of thick slices and inter-slice gaps. Distinguishing between PVS and other hyperintense lesions remains an important target for improvement in computational PVS quantification.

### 4.4. Application studies

The three most frequently applied computational PVS quantification approaches identified in this review were the one firstly proposed by Ballerini et al. (2016) and implemented as part of different frameworks (24 applications), those derived from the work of Lian et al. (2018) (10 applications), and morphological detection as proposed by Schwartz et al. (2019) (6 applications). Ballerini et al.’s approach involved a Frangi filter approach with thresholding optimised based on visual PVS scores. Lian et al.’s approach was a deep learning (CNN) method, involving image enhancement and denoising stages prior to deploying a fully-automated CNN, trained on manually segmented PVS data. Schwartz et al. segmented PVS using a stepwise local homogeneity search of a white matter-masked T1w image, with FLAIR hyperintensity constraints and further linearity, width, and volume constraints. While these three approaches are the most frequently implemented, this is driven by the research groups in which the methods were developed, rather than an indication of a unification of PVS quantification approaches.

Factors identified in common among application studies included age, sex, hypertension, diabetes, WMH, sleep, and cognition, although comparable results were only available for a modest number of studies per factor. Other factors, like ethnicity, are starting to emerge, with two studies (Charisis et al., 2023; Shih et al., 2023) reporting differences in the spatial distribution (Charisis et al., 2023) and volume fraction (Shih et al., 2023) of PVS across racial/ethnic groups. Computational PVS measures used varied, but commonly included PVS count and volume, often segregated by region (e.g., basal ganglia PVS, centrum semiovale/white matter PVS). Increased age, hypertension, WMH, and sleep parameters were broadly associated with computational PVS metrics, although the only one that consistently (i.e., in all the nine studies that explored it) showed an association (in this case positive) with PVS burden was WMH either total volume or visual scores. This was followed by hypertension, with eleven out of twelve studies identifying a positive association between hypertension and PVS burden. The associations between age and PVS burden and hypertension and PVS burden have been previously documented in the literature, and been identified in non-computational studies (Francis et al., 2019). However, this review, different from previous reviews, as includes recent and large population studies involving adults from the whole lifespan or children and adolescents, points at the existence of various patterns of associations and correlations between age and PVS burden, with no association (Piantino et al., 2020; Soitgu et al., 2023) or even negative associations in some brain regions (Lynch et al., 2023) in early life, a negative correlation until the mid-30s (Kim et al., 2023a), and a positive association/correlation afterwards with different strength degrees (Kim et al., 2023).

Other equally large population studies covering a large lifespan point to some factors that may relate to these findings. For example, Shih et al., (2023) report that body mass index (BMI) influenced the basal ganglia PVS burden across middle-aged and older participants. A large GWAS study (Duperron et al., (2023) investigating the associations of small vessel disease markers found that 24 genome-wide significant PVS risk loci were associated with white matter PVS only in young adults, suggesting a genetically linked effect of age in PVS burden. In relation to sleep, the eight studies that explored its association with PVS evaluated different parameters: sleep efficiency, interruptions, quality, duration, falling asleep at different times during the day, for just mentioning some. But all results suggest a positive association between PVS burden and poor sleep health. The dimension of the relationship between sleep and PVS (e.g., in clinical populations with different characteristics) has been less extensively explored, and may be a good candidate for further research.

Associations between sex and PVS burden were mixed, with two studies out of nine finding no association (X. Wang, Chappell, et al., 2016; Kim et al., 2023b), one reporting that PVS burden was higher in females (Sepehrband et al., 2021), and six reporting that PVS burden was higher in males. These findings reflect the broader literature, which often has not reported on sex differences, or reports that potential sex differences in PVS burden remain unclear. Similarly, mixed results were identified for associations between PVS burden and diabetes. Of the four studies that referred to diabetes, two found no association (Ballerini et al., 2019; X. Wang, Chappell, et al., 2016) and two found a positive association between diabetes and white matter PVS (S. Wang et al., 2021; Zebarth et al., 2023). A previous systematic review and meta-analysis reported no associations identified between diabetes and PVS burden (Francis et al., 2019), but studies reporting on diabetes in relation to PVS burden are too few to draw any substantial conclusions as of yet.

PVS burden has been explored in association with cognition in a number of studies. In the present review, we found studies reported mixed evidence for poorer performance on cognitive testing in those with greater PVS burden. Two of the fifteen studies identified that examined cognition relied on a measure of total SVD burden, rather than PVS-specific associations. Both reported negative associations between SVD burden and cognitive decline (particularly decline in processing speed; Hamilton, Cox, Ballerini, et al., 2021; Hamilton, Cox, Okely, et al., 2021). From the rest, three studies reported no association between PVS burden and measures of cognition (Ramirez et al., 2022; Valdés Hernández et al., 2020; Soitgu et al., 2023), and nine indicated a positive association between poor cognition, at least in one domain, and PVS burden. But those indicating a positive association studied specific clinical populations for which cognitive impairment was one of their phenotypes: Huntington’s disease (Valdés Hernández et al., 2019; Li et al., 2023), CADASIL (Karvelas et al., 2023), TBI (Hicks et al., 2023), MCI (Seperhband et al., 2021; Bown et al., 2023; Kamagata et al., 2023), frontotemporal dementia (Moses et al., 2022), and AD (Kamagata et al., 2023). One study involving elderly patients aged 65-84 enrolled in the LADIS study (Pantoni et al., 2005), also reported a positive association between PVS burden and cognition. But all participants in this study had mild cognitive complaints, motor disturbances, minor cerebrovascular events, mood disturbances, and/or other neurological problems (Jokinen et al., 2020). Other systematic reviews and meta-analyses in recent years have also yielded mixed results as to the association between PVS and cognition (Francis et al., 2019; Hilal et al., 2018; Jie et al., 2020), with further research required to shed a light on the role of PVS by stage in cognitive decline, since PVS could increase or decrease depending on compensatory mechanisms, or as part of possibly a neuroprotective or coping mechanism, prior to or amidst continual decline in cognitive functions.

### 4.5. Strengths and Limitations

The systematic nature of this review, the broad search terminology and inclusion criteria, and the large number of years covered (i.e., up until September 2023), are perhaps the main strengths of this work. Although there would have been sources missed due to database indexing issues, articles not being published in English language, or access restrictions, the systematic and inclusive nature of our work have allowed us to cover most of the literature on the theme published up to this date, distil, and make available data to inform future research in the field of PVS research. We have summarised and meta-analysed the characteristics, advantages, scope, applicability, and shortcomings of the computational methods developed up to date to assess PVS burden. This work has also covered the works to support the improvement of these methods, establish their limits of validity, and facilitate their use by explaining the effect of tuning their different parameters and drafting recommendations based on the evaluation of the most widely used methods in different settings. But this review covers only findings from computationally-derived measurements. Owed by the availability of databases with imaging and associated neuroradiological assessments and clinical data, still a considerable number of studies use visual scores in their analyses. For illustration, from January to September 2023 alone, our search in Web of Science identified 47 publications of clinical studies with PVS either as outcome measure or as predictor (at least one of them in some cases) that used PVS visual scores, which were excluded from our analyses, against only 23 studies that used computationally-derived metrics and were, therefore, included. Consequently, conclusions cannot be drawn from our findings on factors related to PVS burden as they only reflect results from a percentage of all the studies in this area of research. A wider literature analysis including also data obtained from neuroradiological (i.e., visual) assessments would be necessary to better inform on the associations of PVS with clinical, demographic, genetic, cognitive, and lifestyle factors.

### 4.6. Conclusions and future directions

Computational approaches to PVS quantification have increased in prevalence in recent years. Novel automated PVS quantification approaches often use thresholding and filtering approaches, or deep learning (CNN) approaches, each with different advantages as well as shortcomings. The method to use will, ultimately, depend on resources availability (i.e., ground truth, disposal or not of a pre-trained model, computational capacity, and skills of the researchers/image analysts). Barriers to successful computational PVS quantification, including image compression, image quality, and nature and quality of the reference standard assessments have been identified, and potential solutions have been proposed. In particular, further work is needed to address the issue of lesion type discrimination (e.g., PVS differentiation from lacunes and small WMH in T2w contrast images, and calcified vessels or mineralisation around small arterioles in T1w contrast images). Although some methods were developed using images from different scanners and magnetic field strengths, methods’ performance in the same population imaged at different magnetic field strengths are not known. Best practices and adjustments for assessing longitudinal changes in PVS are also not documented. Also, most methods were developed using representative samples with wide range of PVS and confounding disease burden, but results were not analysed taking these into account. Further studies that develop methods for quantifying disease markers should analyse accuracy results by disease load in order to inform better the scope of their applicability. Computational PVS approaches are increasingly being applied in epidemiological and clinical studies, with age, WMH, hypertension, different diseases involving cognitive deficits as part of their phenotypes, and sleep, emerging as factors associated with PVS burden. More research is needed to better understand the mechanisms and mediating factors in the associations that have been consistently found, and the reasons for discrepancies in the factors that have yielded conflicting results across the studies.

## Data Availability

All data extracted is given in the tables of the manuscript.

