## Supplementary Tables 1 and 3 for "A Systematic Review and Meta-Analysis of Automated Methods for Quantifying Enlarged Perivascular Spaces in the Brain"

Supplementary Table 1. Number and % of publications by year and type

| | All | | Method | | Improvement | | Application | |
|---|---|---|---|---|---|---|---|---|
| | N | % | N | % | N | % | N | % |
| 2002 | 1 | 0.87 | 1 | 2.08 | 0 | 0 | 0 | 0 |
| 2004 | 1 | 0.87 | 1 | 2.08 | 0 | 0 | 0 | 0 |
| 2008 | 2 | 1.74 | 2 | 4.17 | 0 | 0 | 0 | 0 |
| 2009 | 2 | 1.74 | 1 | 2.08 | 1 | 12.5 | 0 | 0 |
| 2011 | 1 | 0.87 | 1 | 2.08 | 0 | 0 | 0 | 0 |
| 2015 | 4 | 3.48 | 3 | 6.25 | 0 | 0 | 1 | 1.69 |
| 2016 | 8 | 6.96 | 7 | 14.58 | 0 | 0 | 1 | 1.69 |
| 2017 | 4 | 3.48 | 4 | 8.33 | 0 | 0 | 0 | 0 |
| 2018 | 6 | 5.22 | 6 | 12.5 | 0 | 0 | 0 | 0 |
| 2019 | 10 | 8.70 | 8 | 16.67 | 0 | 0 | 2 | 3.39 |
| 2020 | 11 | 9.56 | 3 | 6.25 | 1 | 12.5 | 7 | 11.86 |
| 2021 | 15 | 13.04 | 2 | 4.17 | 2 | 25.00 | 11 | 18.64 |
| 2022 | 22 | 19.13 | 7 | 14.58 | 1 | 12.5 | 14 | 23.73 |
| 2023 | 28 | 24.35 | 2 | 4.17 | 3 | 37.5 | 23 | 38.98 |
| Total | 115 | 100 | 48 | 100 | 8 | 100 | 59 | 100 |

Supplementary Table 3. PVS quantification assessment: accuracy metrics

| Study | Values of the accuracy metrics used |
|---|---|
| (Descombes et al., 2004) | Type I error (false positives) 13% and type II error (false negatives) 2.7% |
| (Uchiyama et al., 2008) | AUC=0.945 to classify lacunar infarcts from PVS. |
| (Uchiyama et al., 2009b) | AUC=0.945 to classify lacunar infarcts from PVS, sensitivity=93.3% and specificity=75% for detection of lacunar infarcts. |
| (González-Castro, Valdés Hernández, et al., 2016a) | Accuracy: SIFT: 82.34%, textons: 79.61% |
| (González-Castro, Valdés Hernández, et al., 2016b) | Accuracy = 80.03; True Negative Rate = 79.36; True Positive Rate = 80.67 |
| (Park et al., 2016) | sensitivity = 0.69 (SD 0.09), PPV = 0.80 (SD 0.07) |
| (Dubost et al., 2017) | TPR = 62.0, number of false positives = 1.5 and FDR = 31.4 |
| (Hou et al., 2017) | PPV 0.73 and SN 0.77 |
| (Zhang et al., 2017) | sensitivity = 0.65 (SD 0.04), PPV = 0.68 (SD 0.04) |
| (Lian et al., 2018) | PPV 0.83 and SN 0.74 |

| | |
|---|---|
| (Niazi et al., 2018) | Automated PVS vs. visual counting = 0.77% false positive pixels<br>Automated PVS vs. visual counting = 19.39% false negative pixels |
| (Dubost, Adams, et al., 2019) | MSE=4.85; MSE=4.65 |
| (Dubost, Dünnwald, et al., 2019) | MAE: CNN: 6.39 (CSO), 5.49 (BG), 3.0 (hippocampi); GP-U-Net: 5.58 (CSO), 5.67 (BG), 2.58 (hippocampi). |
| (Sudre et al., 2019) | Sensitivity of 72.7%, median overlap positive agreement of 59% for boxes/voxels agreed by all raters, and 30% when at least one rater disagreed. |
| (van Wijnen et al., 2019) | On test set: EDM: FAUC=45.761, Sensitivity= 53.63; DGM: FAUC=50.757, Sensitivity= 55.26; IDM: FAUC=53.078, Sensitivity=55.35 |
| (Dubost et al., 2020) | Manual x Automated: FAUCs = 72.0 +- 13.3; Sensitivity = 62.1+-8.7; Average number of false positives = 2.33+-1.71, Average number of false negatives = 2.44+-2.01 |
| (Smith et al., 2020) | 83% of PVS match with the ground truth and 94% of the ground truth match with the segmentation. |
| (Ranti et al., 2022) | Manual x Automated: Sensitivity = 82.9, Specificity = 91.9; Semi-automated x Semi-automated (two raters): inter-rater reliability = 97.8. |
| (Spijkerman et al., 2022) | Bland-Altman: smaller PVS count identified by automated method compared to human. |
| (Sudre et al., 2022) 'BigrBrain' | F1=35.81, AED=14.50 ,AVD=45.30 |
| (Sudre et al., 2022) 'Neurophet' | F1=0, AED=29, AVD=390.15 |
| (Sudre et al., 2022) 'TeamTea' | F1=17.12, AED=41, AVD=106.05 |
| (Sudre et al., 2022) 'TheGPU' | F1=38.92, AED=16, AVD=45.20 |
| (Williamson et al., 2022) | Accuracy/AUC of 0.802/0.834 on the training set, 0.768/0.847 on the validation set, and 0.897(95% CI = [0.758, 0.971])/0.879 on the test set. On the held-out test set, specificity=0.96, sensitivity=0.80, and F1=0.86 |
| (Lan et al., 2023) | FP = 0.0093 ± 0.0069. When trained using QC data, FP = 0.0020 ± 0.0026 |
| (Rashid et al., 2023) | sensitivity = 0.82, precision = 0.83 |

*Legend: PVS: Perivascular Spaces, AUC: Area Under the (ROC) Curve, FAUC: F stat (ratio of two variances) of the AUC from two measurements (e.g. manual vs. automatic) CI: Confidence Interval, AED: Absolute Error Difference, AVD: Absolute Volume Difference, DSC: Dice Similarity Coefficient, FP: False Positives, PPV: Positive Predicted Value, MSE: Mean Squared Error, MAE: Mean Absolute Error, F1: accuracy metric that combines the precision and recall scores of a model by computing how many times a model made a correct prediction across the entire dataset, QC: Quality Control, TPR: True Positive Rate, FDR: False Discovery Rate, CNN: Convolutional Neural Network, SIFT: Scale Invariant Feature Transform*
