## Supplementary Table 2 for "A Systematic Review and Meta-Analysis of Automated Methods for Quantifying Enlarged Perivascular Spaces in the Brain"

Supplementary Table 2. Code availability

| Publication | Pre-processing | Method |
|---|---|---|
| Kruggel et al. (2002)[14] | Not provided | Not provided |
| Descombes et al. (2004)[15] | Not provided | Not provided |
| Uchiyama et al. (2008)[16] | Not provided | Not provided |
| Wuerfel et al. (2008)[17] | Not applicable | MedX discontinued |
| Uchiyama et al. (2009)[18] | Not provided | Not provided |
| Ramirez et al. (2011)[19] | Not applicable (part of the Lesion Explorer software) | https://github.com/sbips/sips |
| Cai et al. (2015)[20] | Not applicable | Matlab 2012b: https://www.mathworks.com |
| Pereira et al. (2015)[21] | Not provided | Not provided |
| Ramirez et al. (2015)[8] | Not applicable (part of the Lesion Explorer software) | https://github.com/sbips/sips |
| Ballerini et al. (2016)[22] | Brain extraction uses optiBET: https://montilab.psych.ucla.edu/fmri-wiki/optibet/ White matter segmentation (FSL-FAST): https://fsl.fmrib.ox.ac.uk/fsl/fslwiki/FAST | Uses: https://www.mathworks.com/matlabcentral/fileexchange/24409-hessian-based-frangi-vesselness-filter |
| González-Castro et al. (2016a)[23] | Brain extraction uses optiBET: https://montilab.psych.ucla.edu/fmri-wiki/optibet/ and FSL-FAST https://fsl.fmrib.ox.ac.uk/fsl/fslwiki/FAST ROI segmentation uses FSL-SUSAN and FSL-FIRST: https://fsl.fmrib.ox.ac.uk/fsl/fslwiki/SUSAN and https://fsl.fmrib.ox.ac.uk/fsl/fslwiki/FIRST | Not provided |
| González-Castro et al. (2016b)[24] | Brain extraction uses optiBET: https://montilab.psych.ucla.edu/fmri-wiki/optibet/ and FSL-FAST https://fsl.fmrib.ox.ac.uk/fsl/fslwiki/FAST | Not provided |

| | | |
|---|---|---|
| | ROI segmentation uses FSL-SUSAN and FSL-FIRST:<br>https://fsl.fmrib.ox.ac.uk/fsl/fslwiki/SUSAN and<br>https://fsl.fmrib.ox.ac.uk/fsl/fslwiki/FIRST | |
| Park et al. (2016)[25] | Not provided | Not provided |
| Wang et al. (2016a).[9] | Analyze software: https://analyzedirect.com/ | Analyze software: https://analyzedirect.com/ |
| Zhang et al. (2016)[26] | Skull-stripping uses FSL-BET:<br>https://fsl.fmrib.ox.ac.uk/fsl/fslwiki/BET<br>ROI segmentation uses FSL-FAST:<br>https://fsl.fmrib.ox.ac.uk/fsl/fslwiki/FAST<br>Registration uses FSL-FLIRT:<br>https://fsl.fmrib.ox.ac.uk/fsl/fslwiki/FLIRT | Not provided |
| Zong et al. (2016)[27] | Bias field correction uses N3:<br>http://www.bic.mni.mcgill.ca/software/N3/<br>Skull-stripping uses FSL-BET:<br>https://fsl.fmrib.ox.ac.uk/fsl/fslwiki/BET<br>ROI segmentation uses FSL-FAST:<br>https://fsl.fmrib.ox.ac.uk/fsl/fslwiki/FAST<br>Registration uses FSL-FLIRT:<br>https://fsl.fmrib.ox.ac.uk/fsl/fslwiki/FLIRT | Frangi filter : https://uk.mathworks.com/matlabcentral/fileexchange/24409-hessian-based-frangi-vesselness-filter<br>OOF: https://uk.mathworks.com/matlabcentral/fileexchange/41612-optimally-oriented-flux-oof-for-3d-curvilinear-structure-detection<br>Random forest implementation not provided |
| Dubost et al. (2017)[28] | ROI Segmentation uses Freesurfer:<br>https://surfer.nmr.mgh.harvard.edu/ | Not provided |
| González-Castro et al. (2017)[29] | Brain extraction uses optiBET: https://montilab.psych.ucla.edu/fmri-wiki/optibet/ and FSL-FAST<br>https://fsl.fmrib.ox.ac.uk/fsl/fslwiki/FAST<br>ROI segmentation uses FSL-SUSAN and FSL-FIRST:<br>https://fsl.fmrib.ox.ac.uk/fsl/fslwiki/SUSAN and<br>https://fsl.fmrib.ox.ac.uk/fsl/fslwiki/FIRST | Not provided |
| Hou et al. (2017)[30] | Non-local Haar filter implementation not provided<br>Noise reduction uses BM4D: https://webpages.tuni.fi/foi/GCF-BM3D/ | Frangi filter : https://uk.mathworks.com/matlabcentral/fileexchange/24409-hessian-based-frangi-vesselness-filter<br>OOF: https://uk.mathworks.com/matlabcentral/fileexchange/41612-optimally-oriented-flux-oof-for-3d-curvilinear-structure-detection<br>Random forest implementation not provided |
| Zhang et al. (2017)[31] | Skull-stripping uses FSL-BET:<br>https://fsl.fmrib.ox.ac.uk/fsl/fslwiki/BET | Frangi filter : https://uk.mathworks.com/matlabcentral/fileexchange/24409-hessian-based-frangi-vesselness-filter |

| | | |
|---|---|---|
| | ROI segmentation uses FSL-FAST: https://fsl.fmrib.ox.ac.uk/fsl/fslwiki/FAST<br>Registration uses FSL-FLIRT: https://fsl.fmrib.ox.ac.uk/fsl/fslwiki/FLIRT | OOF: https://uk.mathworks.com/matlabcentral/fileexchange/41612-optimally-oriented-flux-oof-for-3d-curvilinear-structure-detection<br>Random forest implementation not provided |
| Ballerini et al. (2018)[32] | https://imaging.brainlab.ca/lesion-explorer.html | Uses: https://www.mathworks.com/matlabcentral/fileexchange/24409-hessian-based-frangi-vesselness-filter |
| Boespflug et al. (2018)[33] | DICOM to nifti (MRIConvert): https://surfer.nmr.mgh.harvard.edu/fswiki/mri_convert<br>Segmentation (Freesurfer): https://surfer.nmr.mgh.harvard.edu/<br>Bias field correction (Slicer): https://www.slicer.org/<br>Co-registration (to T1w, 3DAllineate from AFNI): https://afni.nimh.nih.gov/pub/dist/doc/program_help/3dAllineate.html | Not provided |
| Jung et al. (2018)[34] | Not provided | Not provided |
| Lian et al. (2018)[35] | Not provided | Not provided |
| Martinez-Ramirez et al. (2018)[36] | Not provided | Not provided |
| Niazi et al. (2018)[37] | Not provided | Not provided |
| Dubost et al. (2019a)[38] | Not provided | Not provided |
| Dubost et al. (2019b)[39] | Bias field correction: http://www.bic.mni.mcgill.ca/software/N3/<br>ROI Segmentation (Freesurfer): https://surfer.nmr.mgh.harvard.edu/ | Not provided |
| Dubost et al. (2019c)[40] | ROI extraction: FSL FIRST and FAST: https://fsl.fmrib.ox.ac.uk/fsl/fslwiki | Not provided |
| Jung et al. (2019)[41] | Not provided | Not provided |
| Schwartz et al. (2019)[42] | DICOM to nifti (MRIConvert): https://surfer.nmr.mgh.harvard.edu/fswiki/mri_convert<br>ROI Segmentation (Freesurfer): https://surfer.nmr.mgh.harvard.edu/<br>Bias field correction (Slicer3): https://www.slicer.org/ | Not provided |

| | | |
|---|---|---|
| Sepehrband et al. (2019)[43] | ROI Segmentation (Freesurfer): https://surfer.nmr.mgh.harvard.edu/ Co-registration: FSL FNIRT: https://fsl.fmrib.ox.ac.uk/fsl/fslwiki/FNIRT/ | Can be requested from: https://sepehrband.github.io/mvp_doc/about/. User guide: https://sepehrband.github.io/mvp_doc/pvs/ |
| Sudre et al. (2019)[44] | Not provided | Not provided |
| Van Wijnen et al. (2019)[45] | CSO Segmentation (Freesurfer): https://surfer.nmr.mgh.harvard.edu/ | https://github.com/kimvwijnen/geodesic.distance.transform |
| Choi et al. (2020)[46] | ROI Segmentation (Freesurfer): https://surfer.nmr.mgh.harvard.edu/ | https://github.com/hufsaim/pvsseg |
| Dubost et al. (2020)[47] | ROI Segmentation (Freesurfer): https://surfer.nmr.mgh.harvard.edu/ | Not provided |
| Smith et al. (2020)[48] | Not applicable | Uses: https://www.mathworks.com/matlabcentral/fileexchange/24409-hessian-based-frangi-vesselness-filter GUI: https://github.com/smithd37/pvssas |
| Boutinaud et al. (2021)[49] | ROI Segmentation (Freesurfer): https://surfer.nmr.mgh.harvard.edu/ | https://github.com/pboutinaud/SHIVA_PVS |
| Yang et al. (2021)[50] | Not provided | Not provided |
| Ranti et al. (2022)[51] | ROI Segmentation (Freesurfer): https://surfer.nmr.mgh.harvard.edu/ | GUI: https://github.com/smithd37/pvssas |
| Spijkerman et al. (2022)[52] | Co-registration: https://elastix.lumc.nl/ | Not provided |
| Sudre et al. (2022a)[53] 'BigrBrain' | Co-registration (rigid to T1w): https://sourceforge.net/p/niftyreg/git/ci/master/tree/ Bias field correction: http://www.bic.mni.mcgill.ca/software/N3/ | https://hub.docker.com/r/whereisvaldo/challenge2021/tags Tag: 1_PVS.bigrbrain, Digest: e5a67e374a66 |
| Sudre et al. (2022b)[53] 'Neurophet' | Co-registration (rigid to T1w): https://sourceforge.net/p/niftyreg/git/ci/master/tree/ Bias field correction: http://www.bic.mni.mcgill.ca/software/N3/ | https://hub.docker.com/r/whereisvaldo/challenge2021/tags Tag: 1_PVS.neurophet, Digest: 0d1276bf5572 |
| Sudre et al. (2022c)[53] 'TeamTea' | Co-registration (rigid to T1w): https://sourceforge.net/p/niftyreg/git/ci/master/tree/ Bias field correction: http://www.bic.mni.mcgill.ca/software/N3/ | https://hub.docker.com/r/whereisvaldo/challenge2021/tags Tag: 1_PVS.teamtea, Digest: fd8f063b6fa9 |
| Sudre et al. (2022d)[53] 'TheGPU' | Co-registration (rigid to T1w): https://sourceforge.net/p/niftyreg/git/ci/master/tree/ Bias field correction: http://www.bic.mni.mcgill.ca/software/N3/ | https://hub.docker.com/r/whereisvaldo/challenge2021/tags Tag: 1_PVS.thegpu, Digest: 7c3c622885c7 |

| | | |
|---|---|---|
| Williamson et al. (2022)[54] | Data storage and de-identification use AMBRA: http://ambrahealth.com<br>Aligning and skull-stripping use AFNI: https://afni.nimh.nih.gov/ | https://github.com/willi3by/PVSNet |
| Lan et al. (2023) | Not provided. However, mentions publicly available tools like Freesurfer (https://surfer.nmr.mgh.harvard.edu/), ITK snap for generating ground truth segmentations (http://www.itksnap.org/pmwiki/pmwiki.php), and co-registration to MNI space using FSL-FNIRT (https://fsl.fmrib.ox.ac.uk/fsl/fslwiki/FNIRT) | Not provided |
| Rashid et al. (2023) | Not provided | Not provided |
